# Visual field evaluation using Zippy Adaptive Threshold Algorithm (ZATA) Standard and ZATA Fast in patients with glaucoma and healthy individuals

**DOI:** 10.1101/2023.01.05.23284206

**Authors:** Pinaz Nasim, S Ve Ramesh, Neetha IR Kuzhuppilly, Preethi Naik, Paul H Artes, Shonraj Ballae Ganeshrao

## Abstract

**Purpose:** To evaluate visual fields using Zippy Adaptive Thresholding Algorithm (ZATA) Standard and ZATA Fast among patients with glaucoma and healthy individuals.

**Methods:** We recruited 22 healthy participants and 55 patients with glaucoma from the ophthalmology clinics at Kasturba Hospital, Manipal, India. Inclusion criteria were age 35 to 85 years, best corrected visual acuity (BCVA) 0.3 logMAR or better. Glaucoma patients had characteristic glaucomatous optic disc damage. All participants were free from any other ocular pathology except from mild cataract. Participants performed ZATA Standard and ZATA Fast tests on a Henson 9000 perimeter and Swedish Interactive Thresholding Algorithm (SITA) Standard and SITA Fast tests on a Humphrey Field Analyser. Tests were repeated within 90 days to evaluate the test-retest variability.

**Results:** The mean difference in the mean deviation (MD) values of ZATA Standard and SITA Standard tests was 1.7 dB, and that in ZATA Fast and SITA Fast tests was 0.9 dB. The sensitivity values of ZATA and SITA tests differed by 3 dB. ZATA Standard and ZATA Fast were 30% and 6% faster than the corresponding SITA tests. Grayscale and probability maps varied slightly between the four tests but represented a similar visual field for most patients in the study.

**Conclusions:** ZATA Standard and ZATA Fast are suitable for clinical practice. However, differences between ZATA and SITA tests suggest that they should not be used interchangeably when monitoring over time.

## Introduction

Glaucoma is an insidious disease that damages the retinal ganglion cells, leads to progressive and irreversible visual field loss, and is a major cause of blindness in India and worldwide.^1–4^ Static Automated Perimetry is a key measure of visual function in glaucoma,^5^ which measures differential luminance sensitivity by testing the ability to detect a low-intensity white light stimulus against a uniformly illuminated white background.^6^

Swedish Interactive Thresholding Algorithm (SITA) Standard and SITA Fast incorporated in the Humphrey Field Analyzer (Carl Zeiss Meditec, Jena, Germany) have been used for over two decades and have been thoroughly evaluated clinically.^7–9^ SITA has become the de-facto standard for visual field evaluation in glaucoma and has been used in trials such as the Ocular Hypertension Study and the United Kingdom Glaucoma Treatment Study.^10,11^

Zippy Adaptive Threshold Algorithms (ZATA) Standard and ZATA Fast, incorporated in the Henson 9000 perimeter (Topcon Healthcare, UK), are more recent innovations, and to date no published evidence of their performance is available. In this study, we focused on the test-retest variability of ZATA Standard and ZATA Fast compared to SITA Standard and SITA Fast tests. Test-retest variability is the random difference observed when tests are repeated.^12^ It is a characteristic of visual field test strategies and is one of the principal limitations of perimetry.^13^

## Methods

### Participants

Patients with glaucoma were consecutively recruited from the ophthalmology clinics of Kasturba Hospital, Manipal Academy of Higher Education (MAHE), Manipal, Karnataka, India. Healthy participants were recruited from people attending their annual health check for employment or self-care. All participants received a complete eye examination. Inclusion criteria were age between 35 to 85 years and a best corrected visual acuity (BCVA) of 0.3 logMAR or better. Glaucoma patients had characteristic optic disc changes such as focal or diffuse neuroretinal rim thinning, localized notching, or nerve fiber layer defects. Only patients with primary glaucoma were included (primary open-angle, primary angle closure, and normal tension), and patients with secondary glaucoma were excluded. We did not include glaucoma patients with a recent history of uncontrolled intraocular pressure, those who were non-compliant with prescribed medications, or those who had previously failed to attend scheduled follow-up visits. All glaucoma patients had previously undergone at least one visual field test.

Healthy participants had no optic disc abnormality or ocular pathology other than mild cataract or a previous history of uncomplicated cataract surgery. All glaucoma patients and healthy participants volunteered to complete four visual field tests within approximately 90 minutes. On the baseline study visit, all participants underwent a demonstration visual field test before the study tests.

### Ethics

The study was approved by the Institutional Ethics Committee, Kasturba Medical College, and Kasturba Hospital (approval number: 370/2020). Written informed consent was obtained from all participants following a detailed explanation of the study in the participants’ preferred language (Kannada or English).

### Visual Fields

The four visual field tests performed were SITA Standard, SITA Fast, ZATA Standard, and ZATA Fast. Humphrey Field Analyser (HFA) 720i and 840 (Carl Zeiss Meditec, Jena, Germany) were used for SITA Standard and SITA Fast tests. The Henson 9000 perimeter (Topcon Healthcare, UK) was used for ZATA Standard and ZATA Fast tests. In both perimeters, the background intensity was 10 cd/m^2^. The maximum stimulus intensity in the HFA and the Henson 9000 perimeter is 3183 cd/m^2^. The four strategies used in the study are briefly explained below.

SITA strategies are a combination of staircase and Bayesian algorithms that use prior knowledge of age-corrected normal sensitivities and inter-subject variability of glaucomatous and normal visual fields.^14^ Based on this prior information, SITA builds two models (so-called “likelihood functions”) during the test, one for patients with glaucoma and another for normal observers.^14^ Following a failure to detect a stimulus, the likelihood decreases for the “normal” model and increases for the “glaucoma” model, and the opposite occurs following successful detection.^14^ The sequence of stimulus presentation is similar to a full-threshold staircase.^14^ The SITA strategies follow a growth pattern where thresholds are first measured at four “seed” locations, one in each quadrant at 12.7 degrees from fixation, and these thresholds are subsequently used to set starting stimulus intensities for neighboring untested locations.^14^ The difference between SITA Standard and SITA Fast is that the latter provides for a slightly greater tolerance to error and therefore requires fewer stimulus presentations than SITA Standard.^15^

ZATA Standard and ZATA Fast are also Bayesian algorithms. ZATA tests have only one prior model for each location, which moves toward higher sensitivities after successful detection and lower sensitivities after failure to detect. The first stimulus presented at each location is 4 dB brighter than the age-matched data. Locations at which the first stimulus was detected are not tested further (David B Henson, personal communication, 2022). For subsequent visits, the patient’s previous response becomes prior information for ZATA strategies. In contrast with SITA, ZATA does not follow a defined growth pattern. ZATA has two versions, Standard and Fast, which differ in stopping criteria.

### Testing Protocol

All visual field tests were conducted on a 24-2 test pattern with Goldmann size III stimuli. The order of tests between ZATA and SITA strategies was chosen randomly by a coin flip. Participants were allowed a break of 5 to 15 minutes between tests to avoid fatigue. Glaucoma patients also underwent visual field tests on the eye not included in the study if clinically indicated. All tests were repeated within ninety days to evaluate the test-retest variability of the visual field tests.

## Data Analysis

The visual field data were converted to right eye format, and blind-spot locations (15, -3 degrees and 15, 3 degrees) were excluded from further analysis. The data analysis was conducted using R version 4.0.3.^16^ Mean deviation (MD) values, sensitivity, and test duration of ZATA Standard and ZATA Fast were compared to SITA Standard and SITA Fast.

## Results

### Participants

Visual field data were obtained from 55 patients with glaucoma and 22 healthy participants. All tests were repeated within a median (25^th^, 75^th^ percentile) of 16 (8, 27) and 14 (7, 25) days of the first visit by healthy participants and glaucoma patients, respectively. Demographic and ocular characteristics of the two groups are shown in *Table 1*.

**Table 1:** Details of the study participants. Data shown are median (25^th^, 75^th^ percentiles) for continuous data and counts (percentages) for nominal data. NA=not available; MD = mean deviation BCVA = best corrected visual acuity

|  | **Glaucoma patients**<br>**(n=55)** | **Healthy participants**<br>**(n=22)** |
| --- | --- | --- |
| **Age (years)** | 64 (55, 71) | 56 (45, 63) |
| **MD (dB)** | -7.6 (-15.3, -2.7) | -0.6 (-1.7, 0.2) |
| **BCVA (logMAR)** | 0.10 (0.00, 0.30) | 0.00 (0.00, 0.10) |
| **Inter-visit interval (days)** | 14 (7, 25) | 16 (8, 27) |
| **Female** | 27 (49%) | 13 (59%) |
| **Open Angle Glaucoma (OAG)** | 35 (63%) | NA |
| **Angle Closure Glaucoma (ACG)** | 20 (36%) | NA |
| **Pseudophakic** | 11 (20%) | 0 |

**Table 2:** Median (25^th^, *75*^th^ percentile) of mean deviation (MD) values and their differences across the two visits obtained from the four tests

| Test | Baseline MD<br>(dB) | Retest MD<br>(dB) | Difference<br>(dB) | Wilcoxon<br>p-value |
| --- | --- | --- | --- | --- |
| **SITA Standard** | -3.6 (-11.3, -0.8) | -2.8 (-9.4, -0.7) | -0.4 (-1.8, 0.6) | 0.48 |
| **ZATA Standard** | -0.9 (-8.2, 0.2) | -1.2 (-10.6, 0.3) | 0.0 (-0.8, 1.2) | 0.91 |
| **SITA Fast** | -3.5 (-10.5, -1.1) | -3.1 (-9.5, -1.0) | -0.2 (-0.9, 0.8) | 0.75 |
| **ZATA Fast** | -1.5 (-11.2, 0.2) | -1.4 (-10.6, 0.4) | -0.3 (-1.1, 0.3) | 0.54 |

The distribution of the MD and the Pattern Standard Deviation (PSD) values of all participants are illustrated in *Figure 1*.

**Figure 1:**
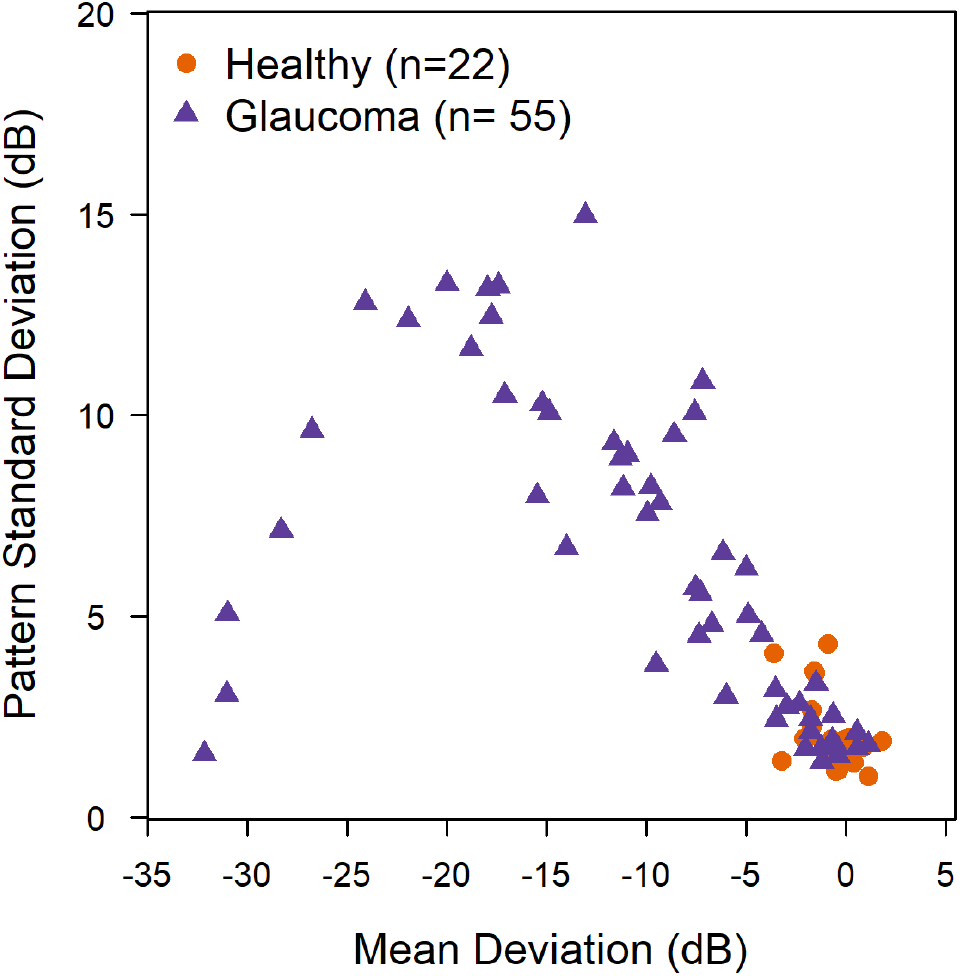
Distribution of mean deviation and pattern standard deviation of the SITA Standard test performed on the baseline study visit.

### MD values of ZATA Standard and ZATA Fast compared to SITA Standard and SITA Fast

The MD value provides an index of the overall visual field damage,^17^ with more negative MD values indicating more severe visual field damage.

The Bland-Altman “limits of agreement” plot in *figure 2* shows the difference between MD values of ZATA and SITA tests as a function of the average MD. The shaded region encompasses 95% of the data (mean ±1.96 SD of the differences).

**Figure 2:**
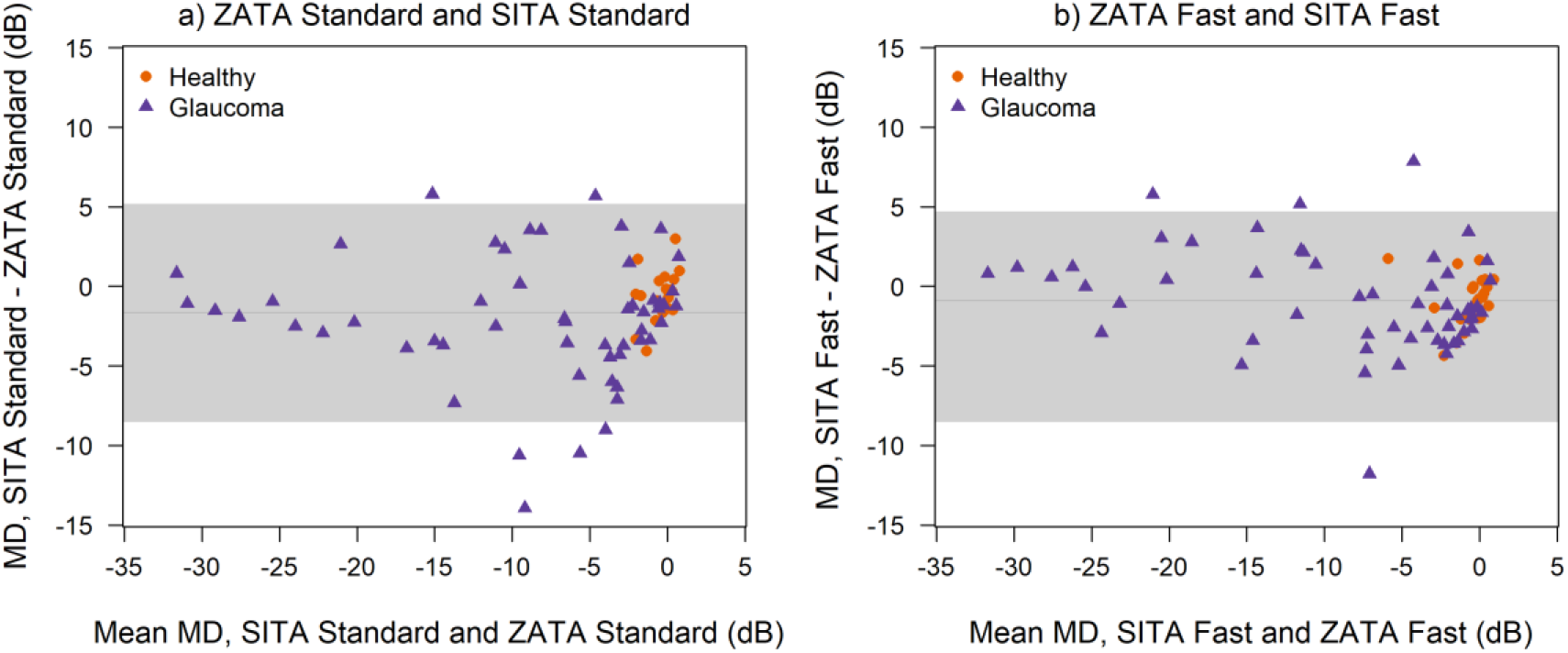
Bland-Altman plots showing mean deviation (MD) values a) ZATA Standard and SITA Standard, b) ZATA Fast and SITA Fast.

The mean (95% CI) difference in the MD values of the ZATA Standard and SITA Standard was 1.7 (−8.5, 5.2) dB (*figure 2a*), and that in the MD values of ZATA Fast and SITA Fast was 0.9 (−6.4, 4.7) dB (*figure 2b*).

### Test-retest variability of MD values

Results of Bland-Altman analysis of test-retest variability in MD (the difference in the MD values across the two visits) in the four tests are shown in *figure 3*. The shaded region in the plot is the ±1.96 SD of the differences between the test and retest.

**Figure 3:**
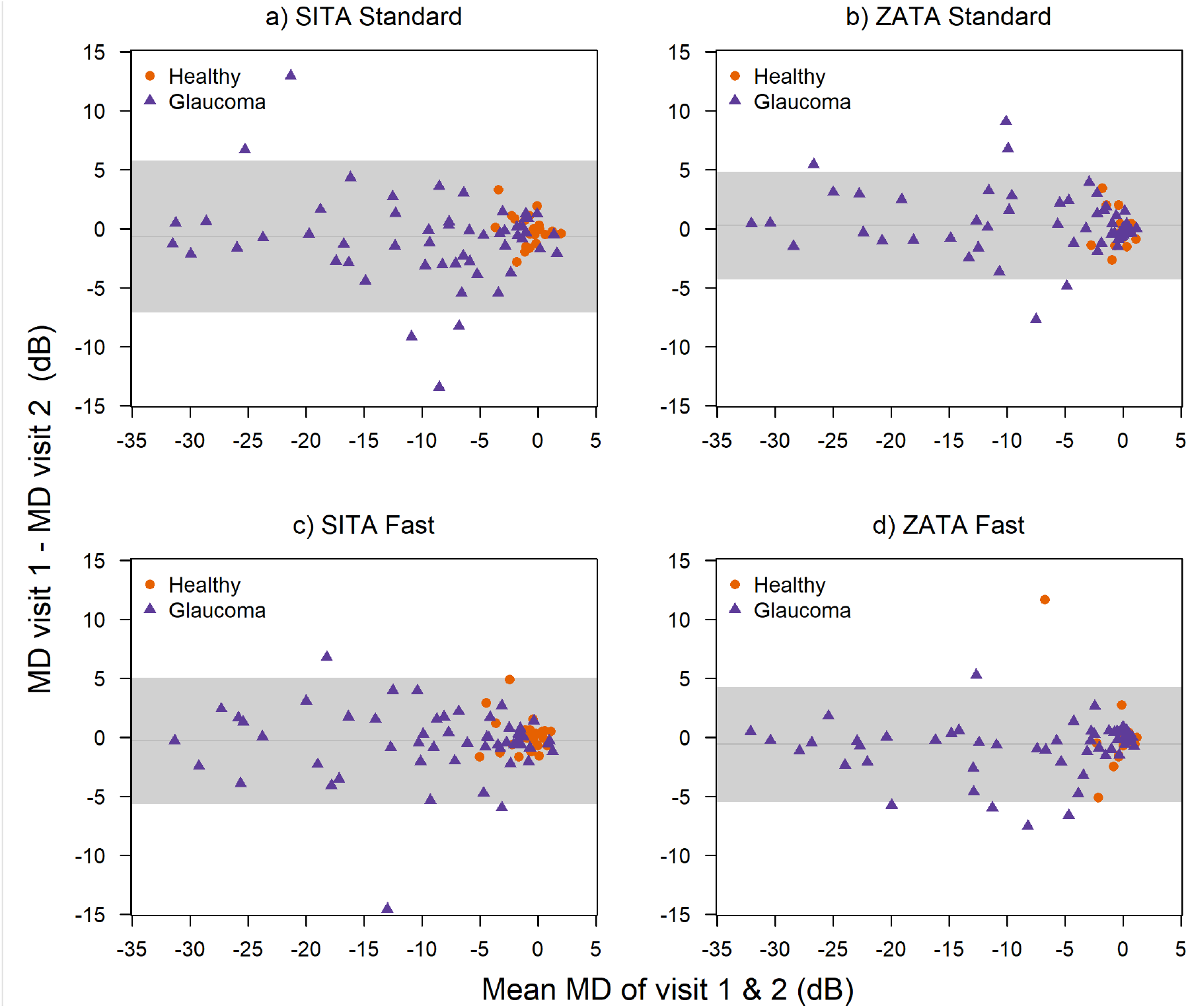
Bland-Altman plots showing mean deviation (MD) values across the two visits for a) SITA Standard, b) ZATA Standard, c) SITA Fast, and d) ZATA Fast.

It represents the overall test-retest variability of the MD values, which appeared similar in the four tests. Data from ZATA tests appeared less dispersed than those from SITA tests. With all four tests, data also appeared less dispersed at more positive MD values (e.g., better than -5 dB), suggesting a relatively lower variability in the MD with less damaged visual fields.

### Frequency distribution of pointwise sensitivity estimates of ZATA Standard, ZATA Fast, SITA Standard, and SITA Fast

*Figure 4a* shows the frequency distribution, and *figure 4b* shows the cumulative distribution of pointwise sensitivity estimates of the ZATA Standard, ZATA Fast, SITA Standard, and SITA Fast strategies. Frequency and cumulative distributions of sensitivity estimates of the SITA Standard test were close to those of the SITA Fast test; similarly, distributions of sensitivity estimates of the ZATA Standard test were close to those of the ZATA Fast test. The first peak of the frequency distribution (plotted as solid circles) was at 0 dB in all four tests (*figure 4a*). With the SITA strategies, 9% to 12% of the total sensitivity estimates were at 0 dB, and 8% to 9% were at 29 dB. With the ZATA strategies, 13% to 15% of the total sensitivity estimates were at 0 dB, and approximately 25% of the sensitivity estimates were at 30 dB (*figure 4a*). The median sensitivity was 30 dB in ZATA Standard and ZATA Fast, compared to 27 dB in SITA Standard and SITA Fast (*figure 4b*).

**Figure 4:**
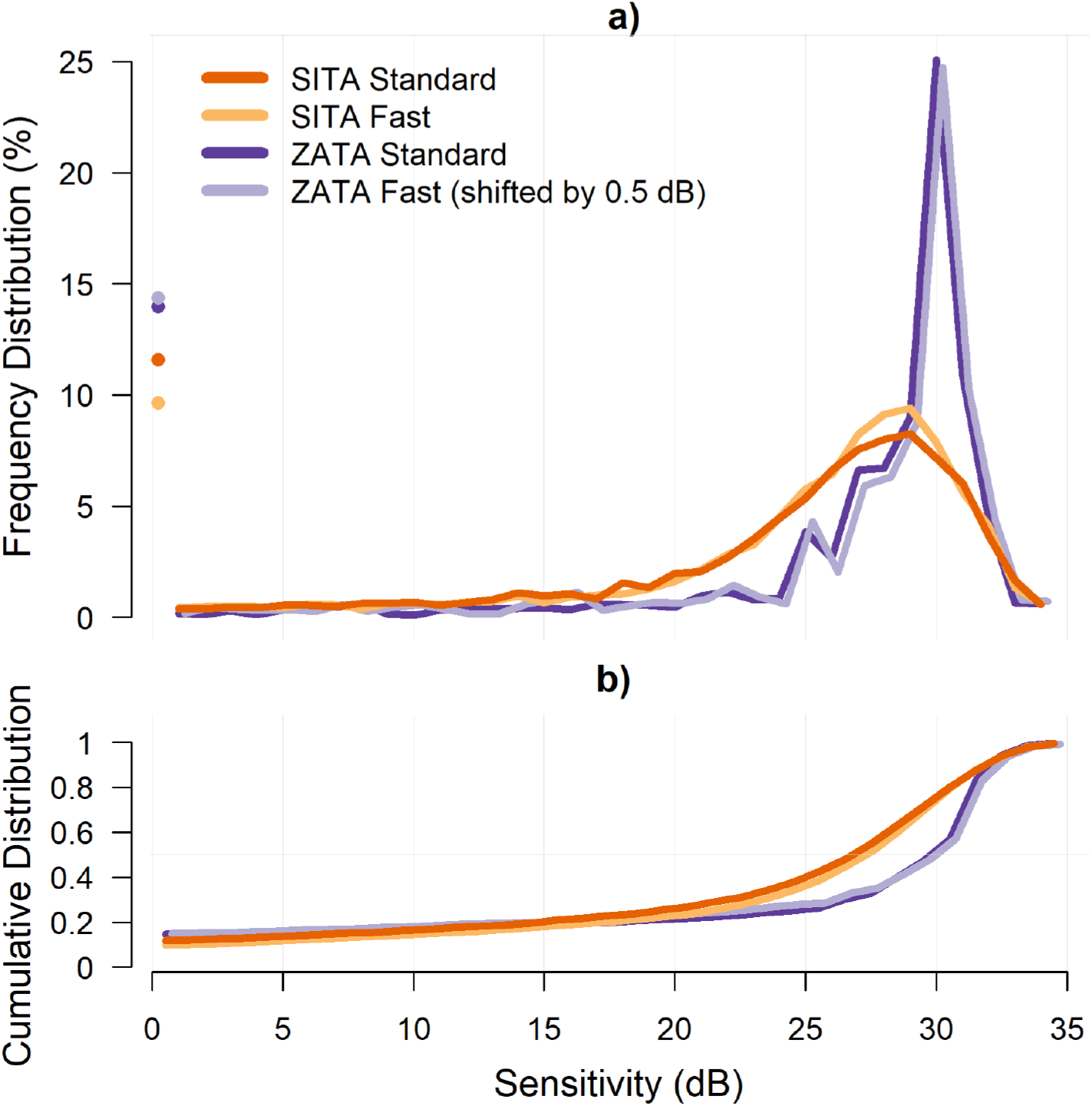
Frequency distribution (figure 4a) and the cumulative distribution (figure 4b) of the sensitivities ranging from 0 to 35 dB, obtained from the four tests across the two visits. The solid circles (figure 4a) represent the percentage of sensitivity estimates equal to 0 dB in the four tests (shown separately for better visibility). The ZATA Fast data have been shifted by 0.25 dB for better visibility.

### Test-retest variability of pointwise sensitivity estimates

*Figure 5* shows the distributions of the pointwise sensitivities on visit 2 (retest) as a function of those at visit 1 (baseline), stratified into broad categories. In the absence of variability, the test and retest sensitivities should fall into the same interval. Clearly, the retest distributions spread out over a much wider range. Tighter distributions (lower variability) are seen at higher baseline sensitivities, and wider distributions (higher variability) at lower baseline sensitivities, such as 0-15 dB and 15-22 dB (*figure 5*).

**Figure 5:**
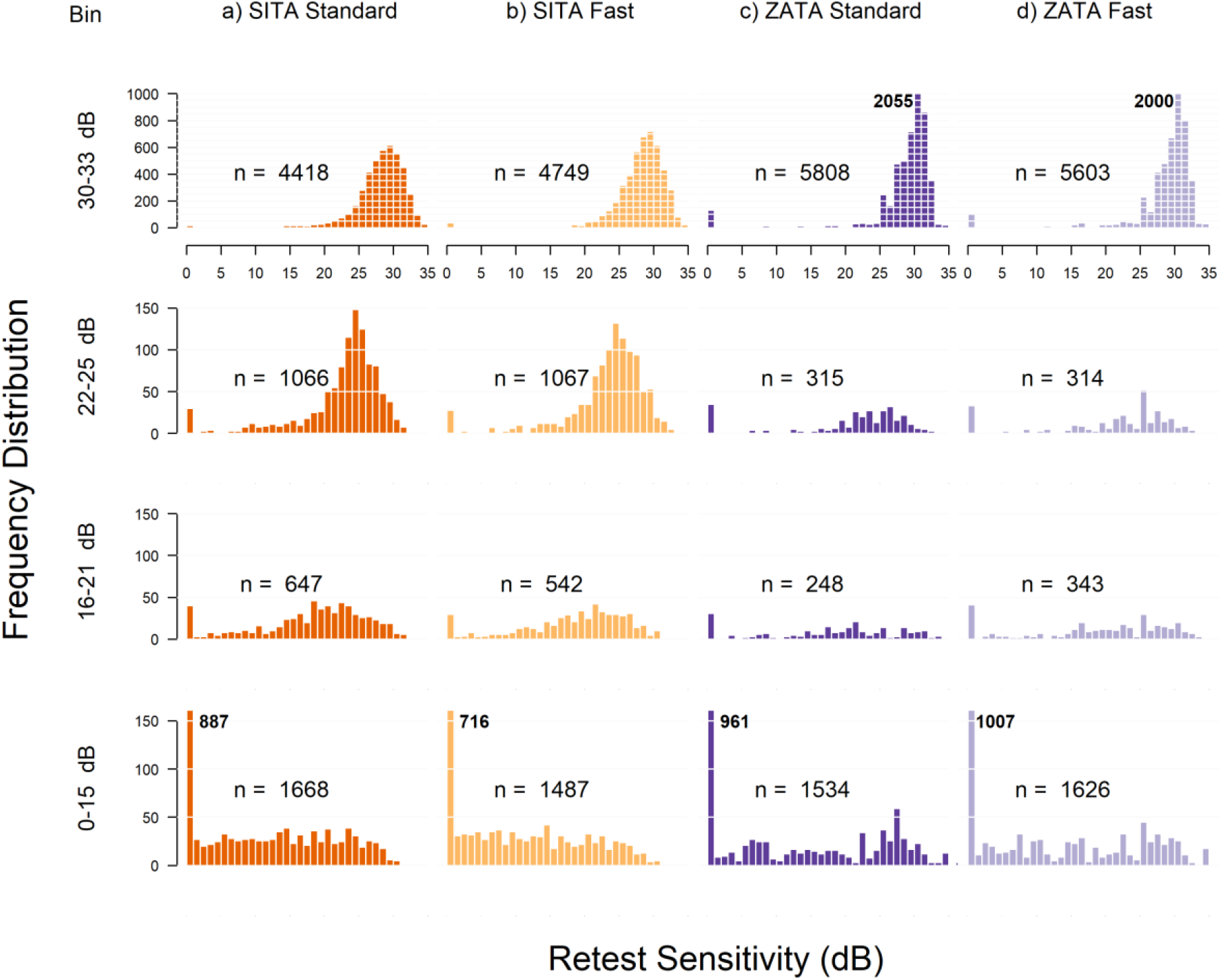
Histograms showing frequency distributions of retest sensitivities for baseline sensitivities stratified as 0-15 dB, 16-21 dB, 22-25 dB, and 30-33 dB. Retest distributions show very high peaks at 0 dB in the first row (0-15 dB baseline sensitivity) and 30 dB in the last row (30-33 dB baseline sensitivity). These peaks have been truncated to facilitate comparison. The bold number beside the peak is the true size of the peak; n = total number of retest points.

*Figure 6* shows the test-retest sensitivity range for the baseline sensitivity on the x-axis. For baseline sensitivity between 1 to 5 dB, the test-retest (10% to 90% percentile) ranged from 0 to 26 dB for SITA tests and from 0 to 28 dB for ZATA tests. At high baseline sensitivity (e.g., 25 to 35 dB), ZATA Standard and ZATA Fast test-retest ranges appeared slightly tighter than those of SITA Standard and SITA Fast tests. For baseline sensitivity ranging from 15 to 25 dB, SITA tests showed a tighter test-retest range than ZATA tests, while ZATA and SITA tests showed similarly wide test-retest ranges at low baseline sensitivity (e.g., 0 to 15 dB).

**Figure 6:**
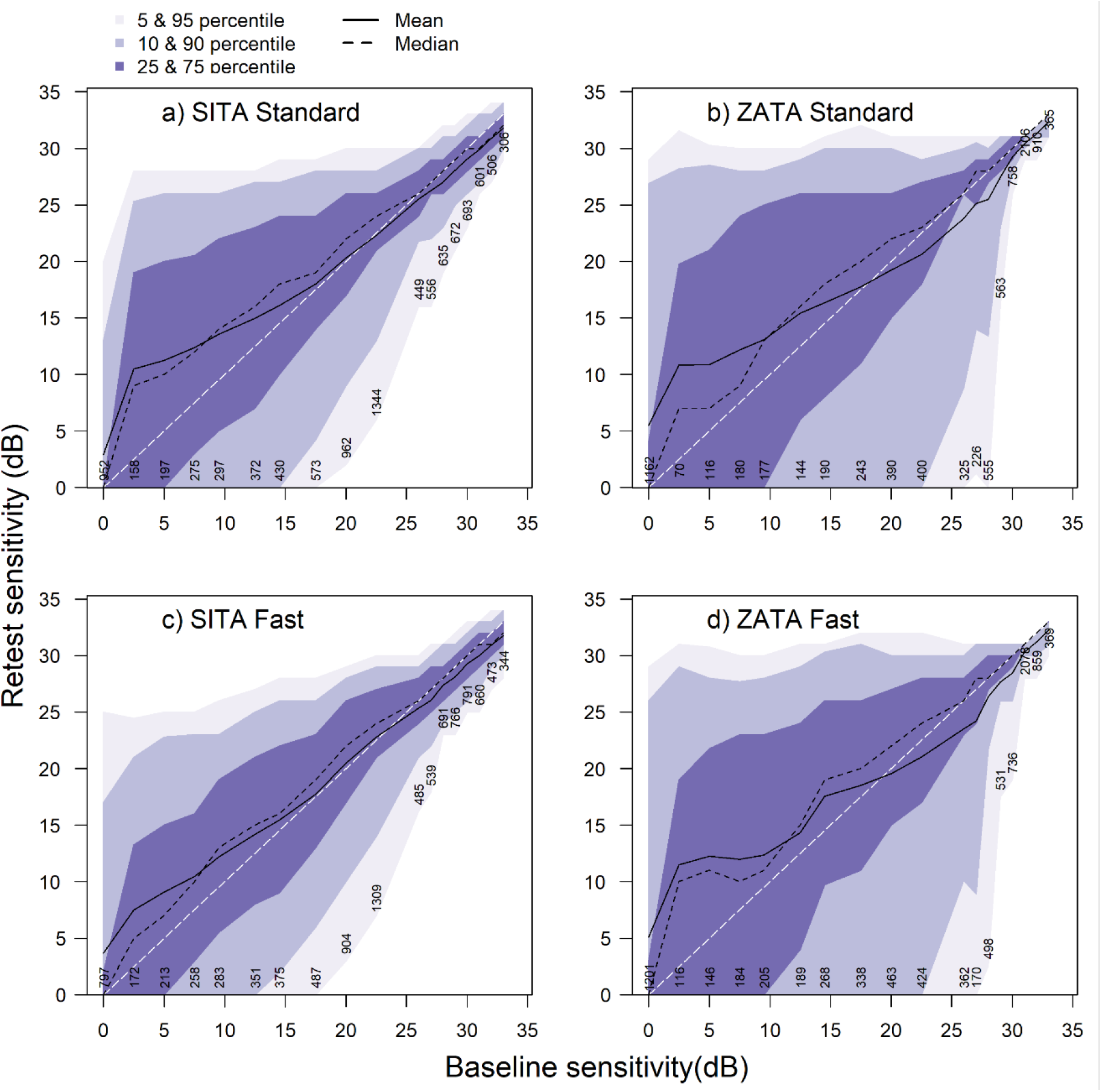
5th and 95th, 10th and 90th, and 25th and 75th percentiles of retest sensitivity at stratified baseline sensitivities. Percentiles of the retest sensitivity range were marked with different shade intensities; light (5th-95th), medium (10th-90th), and dark (25th-75th). The median and mean are indicated by dotted and continuous black lines, respectively.

### Test-duration

All four tests took slightly longer with damaged compared to normal visual fields *(figure 7)*. With ZATA Standard and ZATA Fast, test duration in highly damaged visual fields declined.

**Figure 7:**
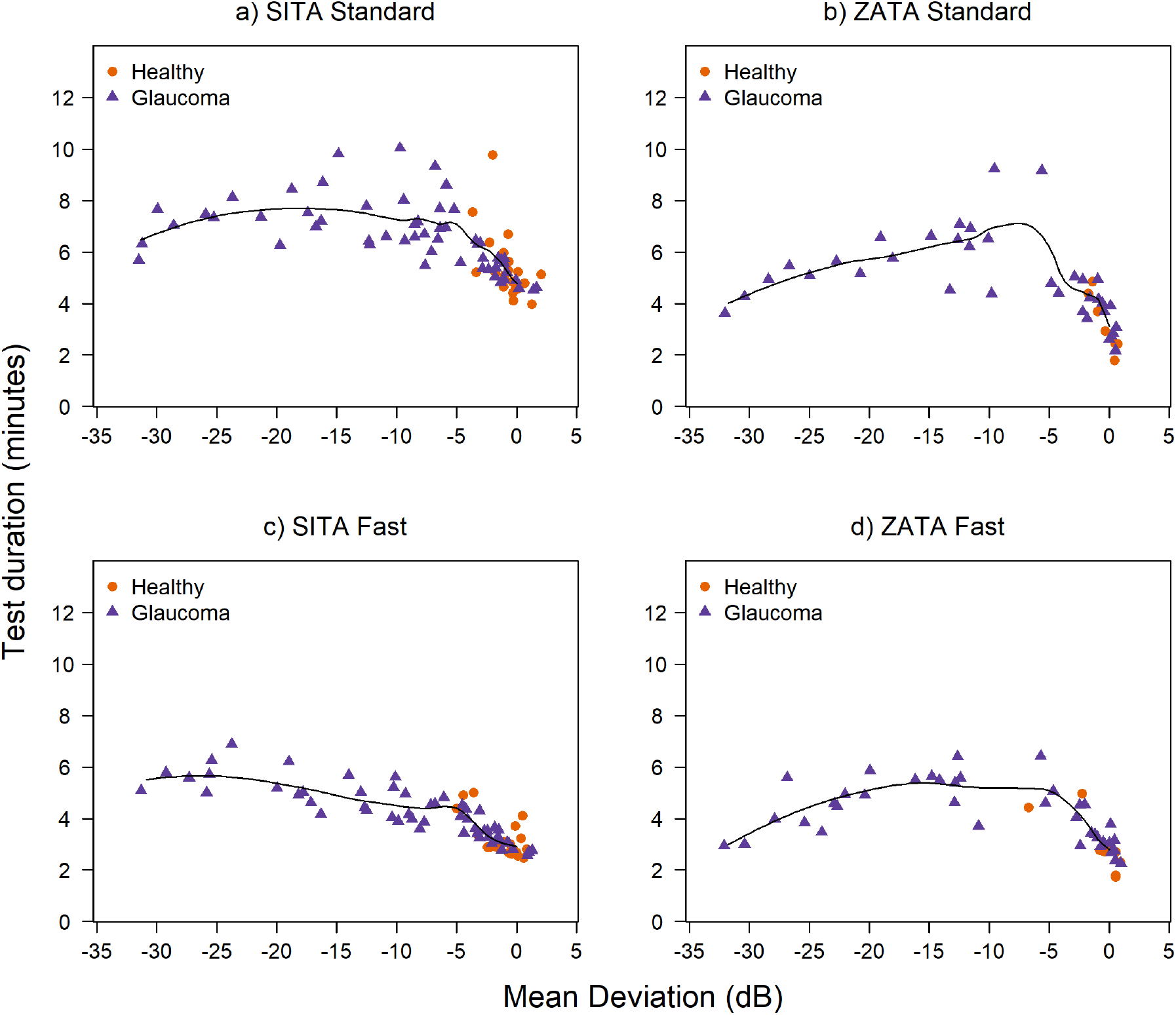
Duration (minutes) taken to perform the tests as a function of mean deviation (MD) values obtained from the tests. A Loess curve^18^ was fitted to the data (solid line). For simplicity, the values of test duration and the MD were averaged across the two visits.

### Visual field representations of ZATA tests compared to SITA tests

*Figures 8a-d* show examples of the visual field defects identified by the four tests. Due to space limitations, only data from selected glaucoma patients are shown here; data from all other patients are given in the supplementary file. The examples contain grayscale and total deviation (TD) and pattern deviation (PD) probability plots from the four tests. The four tests vary slightly in the presentation of the defect in grayscale and probability plots but represent similar visual field damage in almost all cases.

**Figure 8a:**
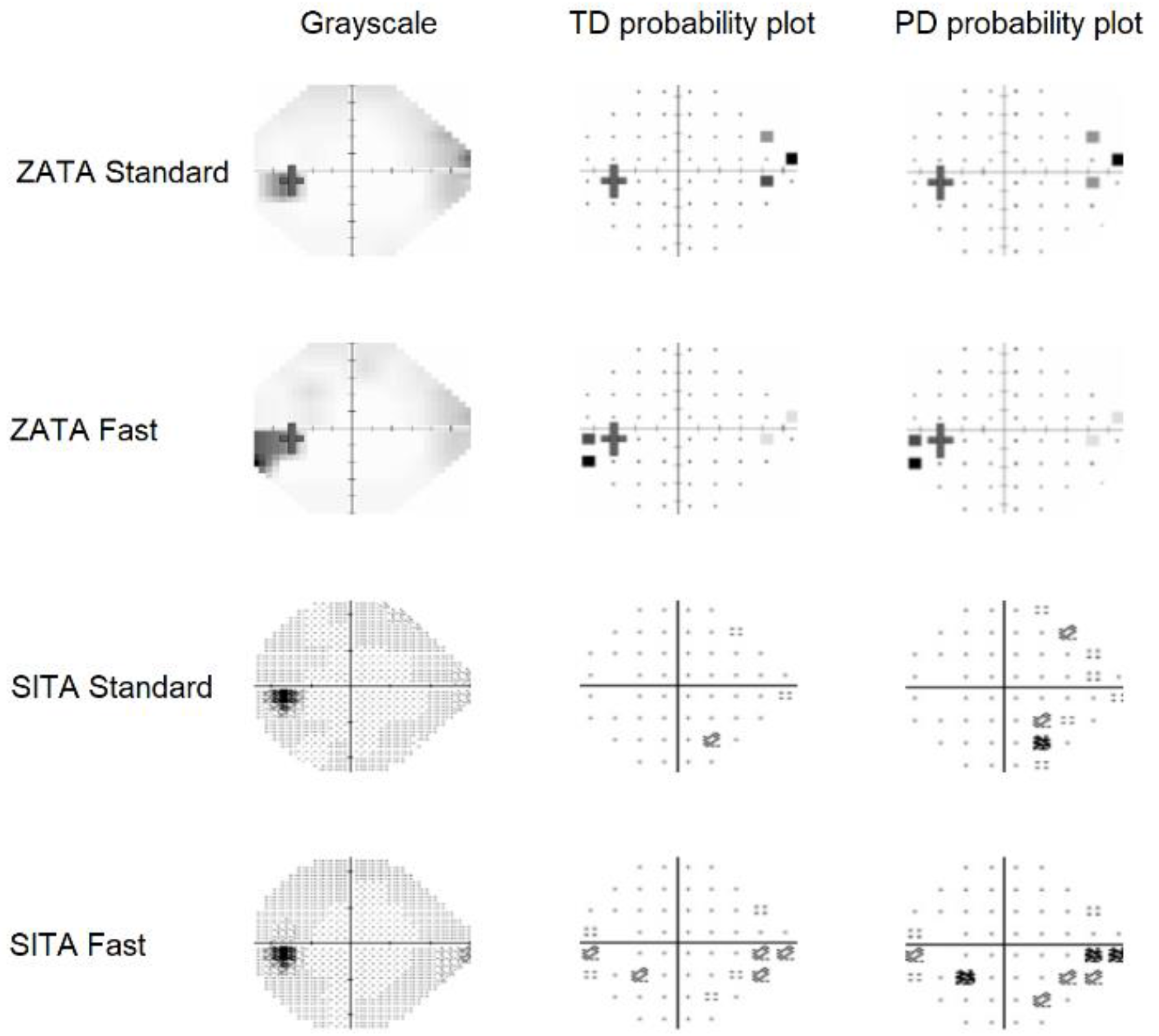
Grayscale, total deviation (TD) probability, and pattern deviation (PD) probability maps from ZATA Standard, ZATA Fast, SITA Standard, and SITA Fast with nasal step defect. The defect depth and the number of defective locations varied in the four tests’ probability maps. ZATA Standard and SITA Fast appeared similar, as do ZATA Fast and SITA Standard.

**Figure 8b:**
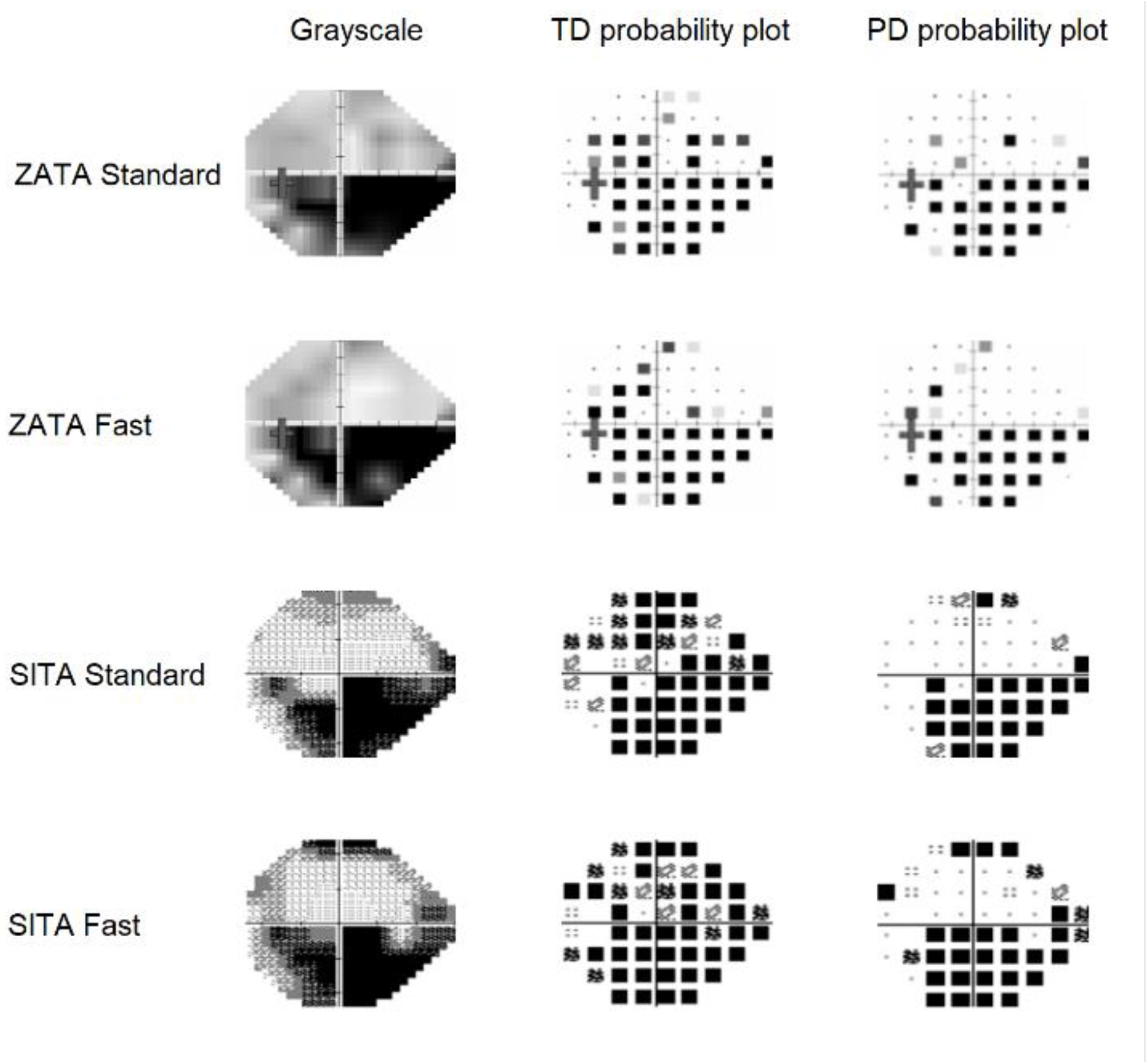
Grayscale, total deviation (TD) probability, and pattern deviation (PD) probability maps from ZATA Standard, ZATA Fast, SITA Standard, and SITA Fast with mainly inferior field defects. The grayscale and the pattern deviation probability maps show a similar pattern in the four tests.

**Figure 8c:**
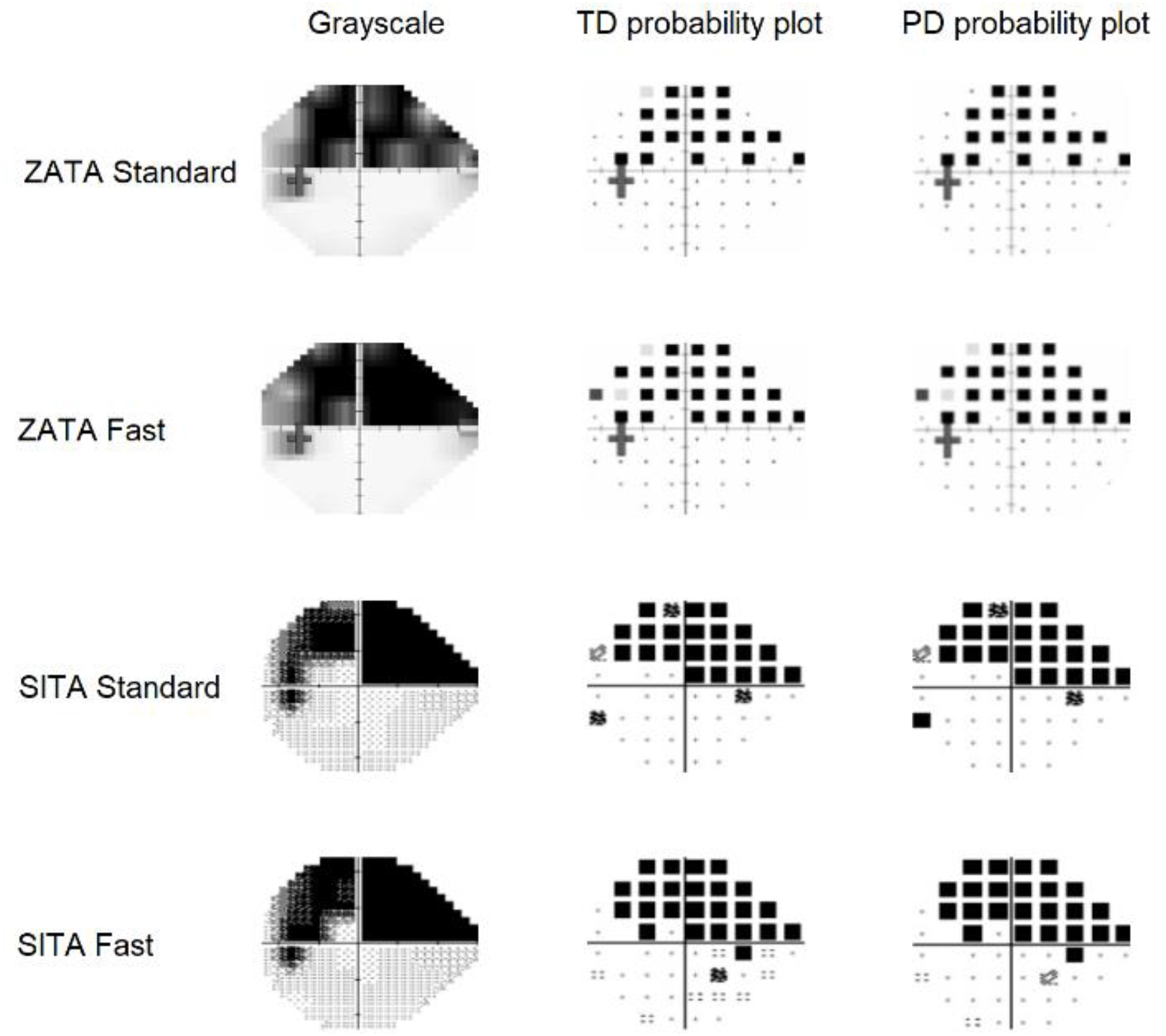
Grayscale, total deviation (TD) probability, and pattern deviation (PD) probability maps from ZATA Standard, ZATA Fast, SITA Standard, and SITA Fast with superior field defect. The grayscale, total deviation, and pattern deviation probability maps show similar defects in SITA and ZATA tests.

**Figure 8d:**
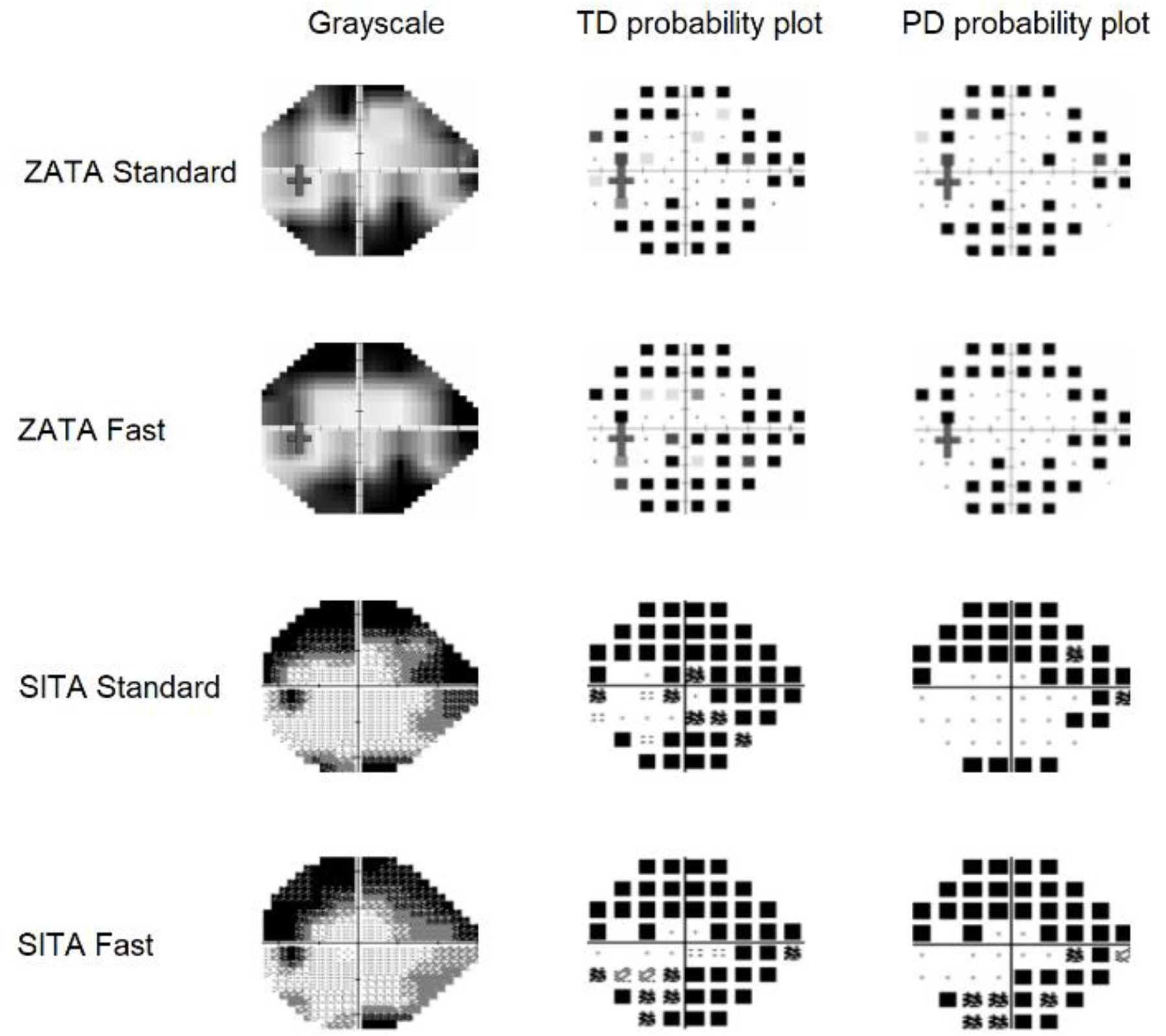
Grayscale, total deviation (TD) probability, and pattern deviation (PD) probability maps from ZATA Standard, ZATA Fast, SITA Standard, and SITA Fast with bi-arcuate scotoma. The defect pattern in the grayscale, total deviation probability map, and pattern deviation probability map slightly varied between ZATA and SITA tests. However, the four tests show a similar type of defect.

## Discussion

The ZATA Standard and ZATA Fast tests of the Henson 9000 perimeter have not previously been studied in a clinical population. We compared these visual field tests to the widely used SITA Standard and SITA Fast tests of the Humphrey Field Analyser. Our results suggest that ZATA and SITA tests characterized patients’ visual fields similarly and that the two were comparable in most cases.

The MD values obtained from the ZATA and SITA tests agreed to within approximately ±5 dB in almost all patients (see limits of agreement in Fig 2). While this level of agreement can hardly be described as “close”, it is similar to the test-retest intervals of all four tests (Fig. 3). In other words, the random differences between the tests appeared similar to the random differences seen when the tests were repeated. This would suggest that there is no compelling reason to prefer one test over another when patients are investigated for visual field damage. But, when patients are followed over time, the systematic differences between the tests suggest that they should not be interchanged.

In the pointwise analysis, the median sensitivity of the ZATA tests was 3 dB higher than that of the SITA tests. Importantly, the shape of the distributions also differed conspicuously between the ZATA and SITA tests. We believe that these differences between the ZATA and SITA distributions reflect the “prior distributions” as well as how they are used by the two algorithms.^14^ However, these data are not in the public domain and therefore our explanation remains speculative.

The distributions of the retest sensitivities *(figure 5, 6)* are useful for examining in greater detail the properties of the four tests. With both SITA and ZATA tests, the test-retest intervals increased with visual field damage *(figure 6)*, as has been reported by others.^12,19^ These data suggest that ZATA tests have somewhat lower variability than SITA tests in the high sensitivity region, while variability of ZATA is relatively higher in areas of low sensitivity.

Both ZATA and SITA are Bayesian algorithms that have an explicit trade-off between bias and variance.^20^ By permitting somewhat larger bias, the efficiency of threshold estimation can be increased. While it is difficult to investigate the bias-variance characteristics of perimetric test strategies in the real patient data of our study, it is obvious that the sensitivity distributions of the ZATA tests have more pronounced peaks (*figure 4, 5*) than those of SITA. This suggest that the prior distributions provide relatively greater influence on the sensitivity estimates of ZATA as compared to SITA.

The main objectives of visual field tests are to quantify visual field damage and to describe the topographical pattern of damage. Our results demonstrate that grayscale plots and probability maps of the ZATA and SITA tests represent similar visual field damage in almost all patients (f*igure 8*, and supplementary material).

In our study, SITA Standard showed 45% larger test-retest variability than reported previously,^21^ and SITA Fast showed approximately 60% larger test-retest variability compared to a study by Heijl et al.^22^ Our participants are typical for a tertiary care facility in India. They may differ from participants recruited for clinical trials and studies who are typically more highly selected. For example, many participants of our study were inexperienced with visual field tests and had some level of cataract (only 20% were pseudophakes). This may contribute to the larger test-retest variability seen in our study compared to others^22,23^ but it should not affect the comparison between the 4 tests. We also did not exclude test results based on reliability indices since neither false positive or false negative responses or fixation losses are dependable indicators of test reliability.^24,25^

In conclusion, visual fields obtained with the ZATA tests are similar to those obtained with SITA, and therefore these tests can be used in clinical practice. However, ZATA and SITA tests should not be used interchangeably when patients are followed over time.

## Supporting information

Gray scale and probability maps of the participants in the study

## Data Availability

All data produced in the present study are available upon reasonable request to the authors.

https://github.com/PinazNasim/Visual-field-evaluation-using-ZATA.git

## Acknowledgments

We acknowledge Electron Eye Technology, UK, for donating Henson perimeters to the Department of Optometry, Manipal College of Health Professions, Manipal Academy of Higher Education, India.

