## Supplementary material for "Visual field evaluation using Zippy Adaptive Threshold Algorithm (ZATA) Standard and ZATA Fast in patients with glaucoma and healthy individuals": Gray scale and probability maps of the participants in the study

Participant:1

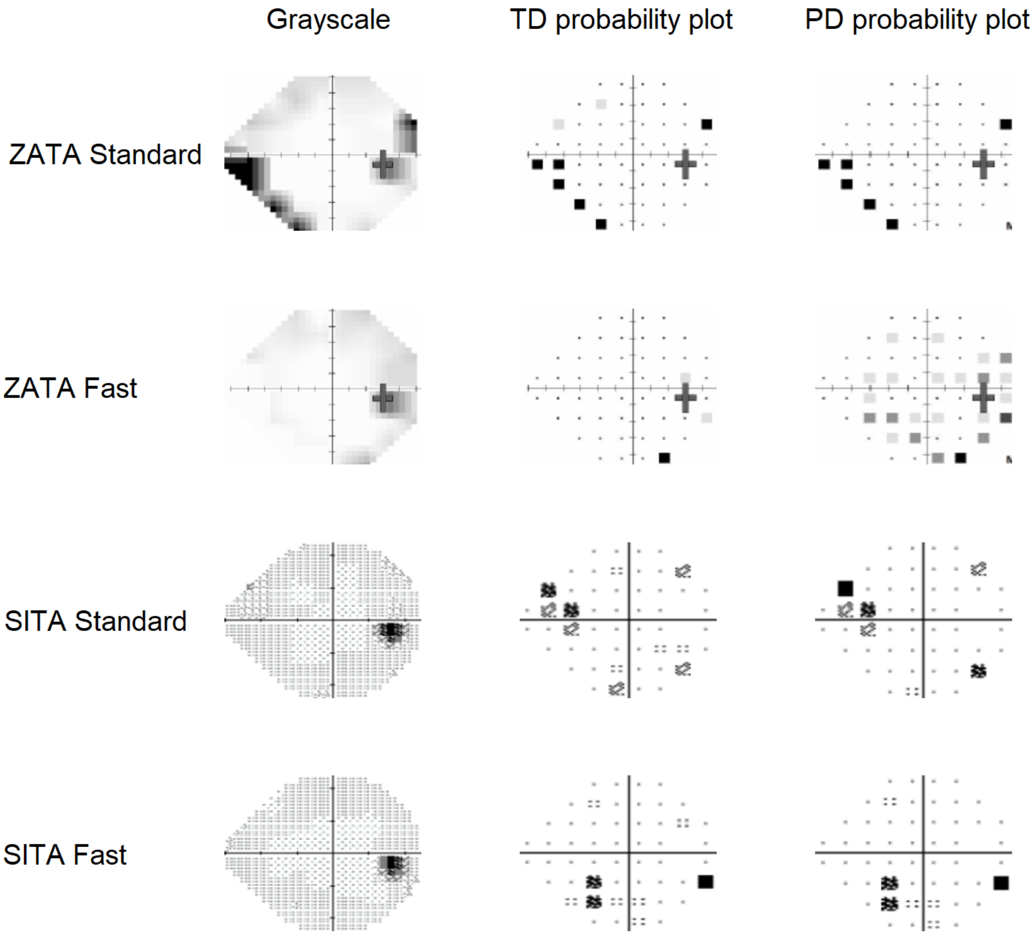

Participant:2

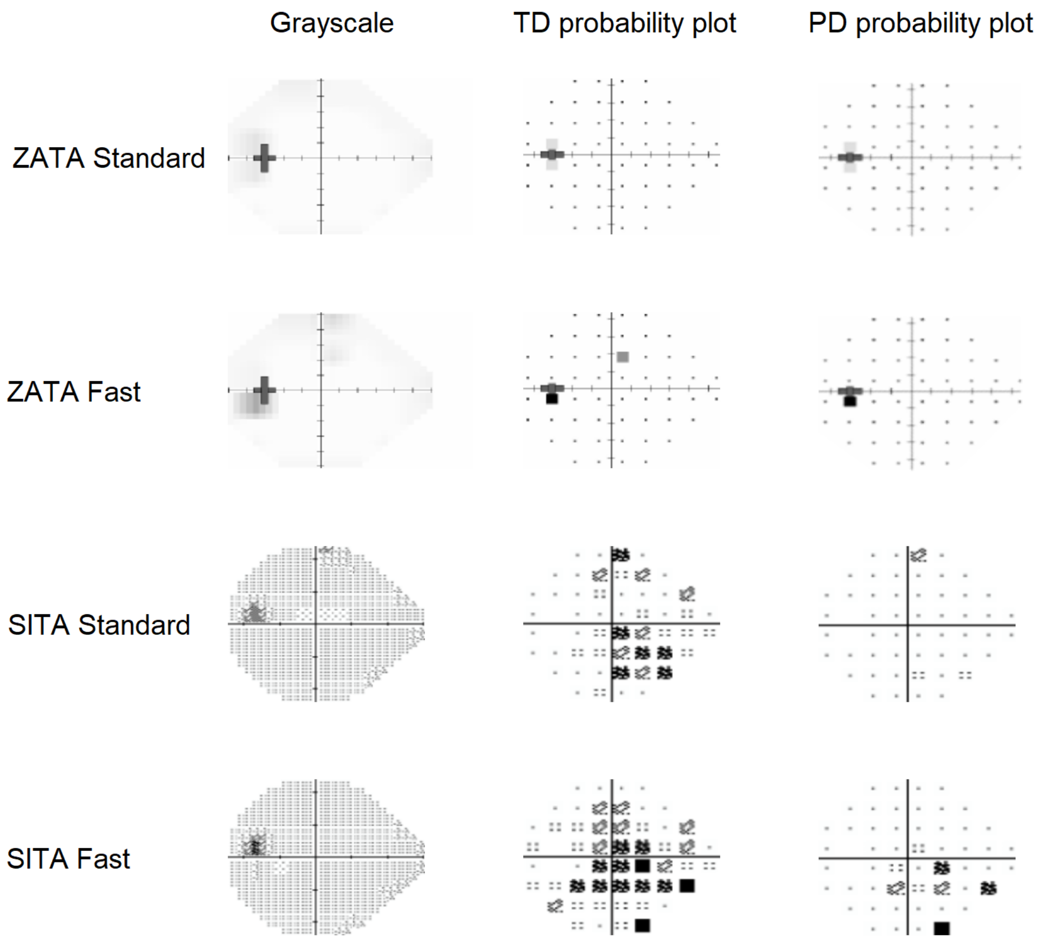

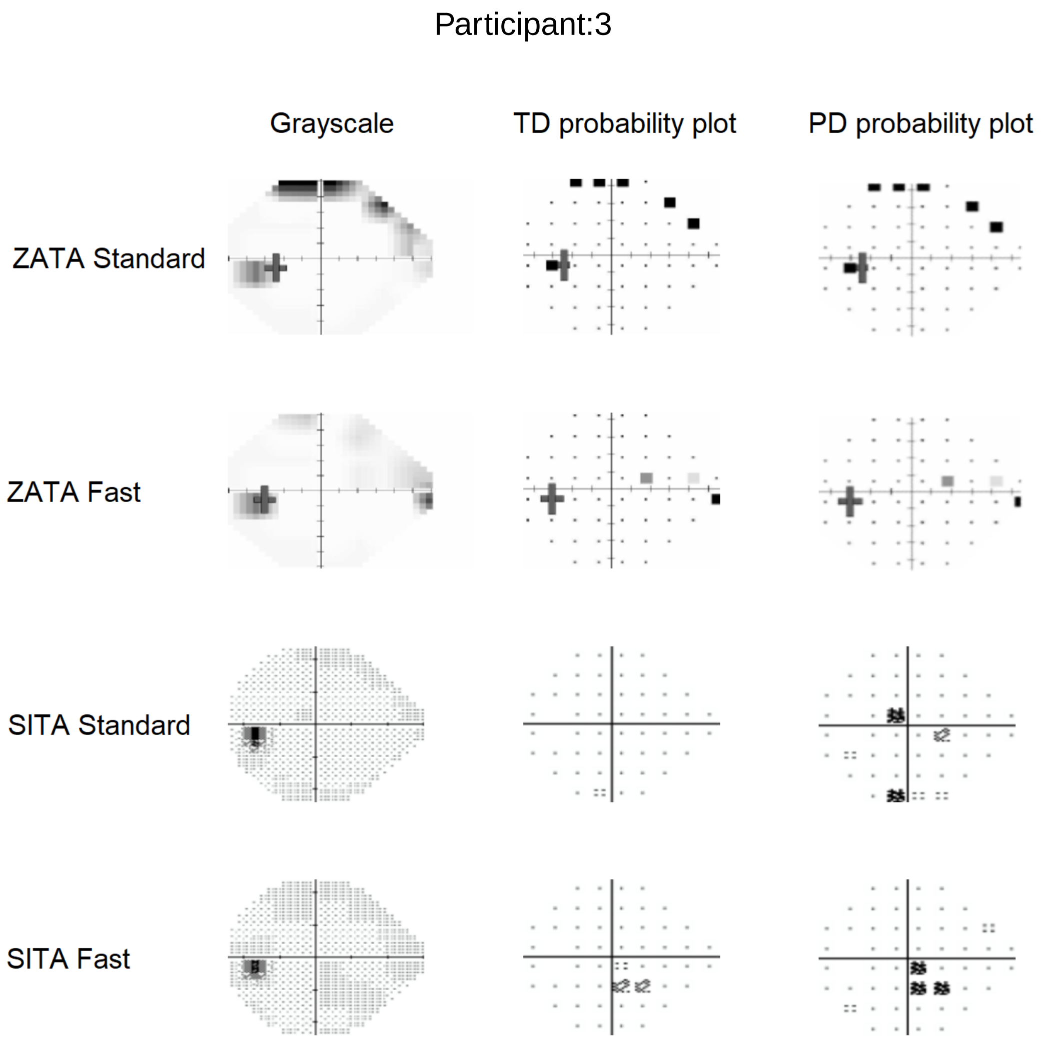
Participant:3
Grayscale
TD probability plot
PD probability plot
ZATA Standard
ZATA Fast
SITA Standard
SITA Fast

### Participant:4

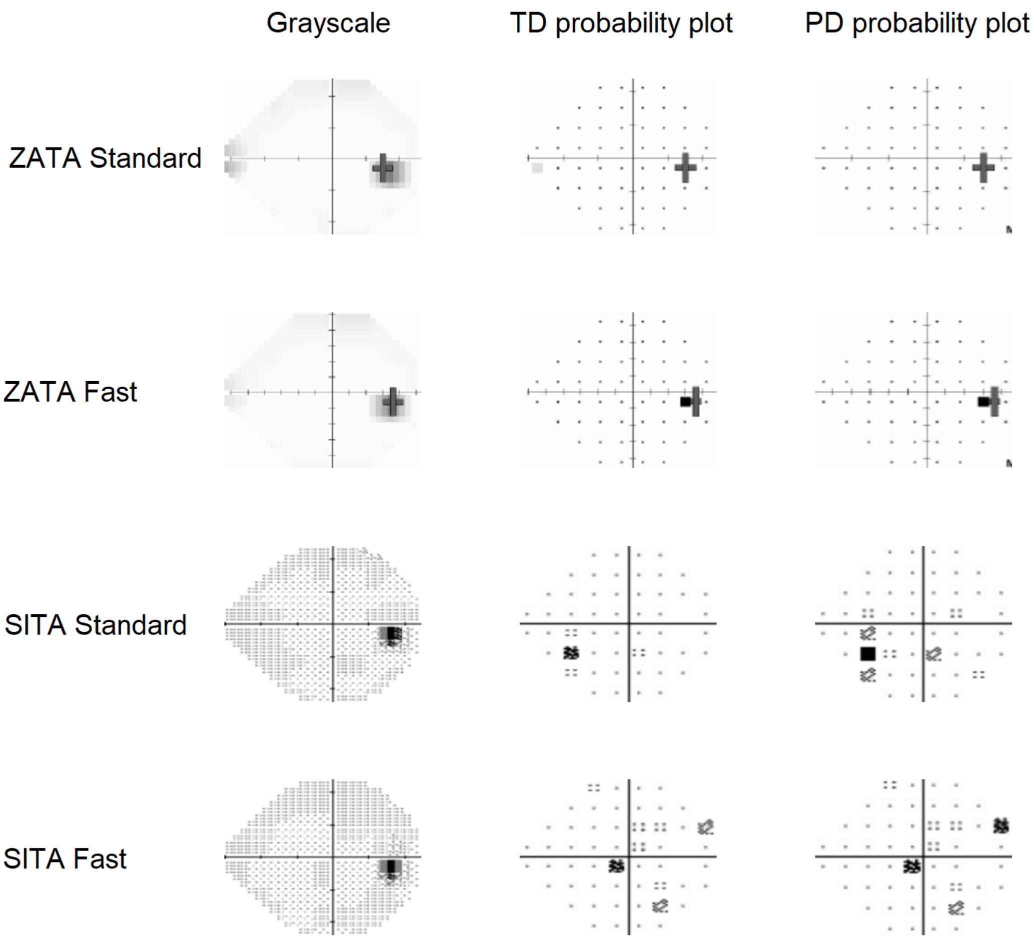

Participant:5

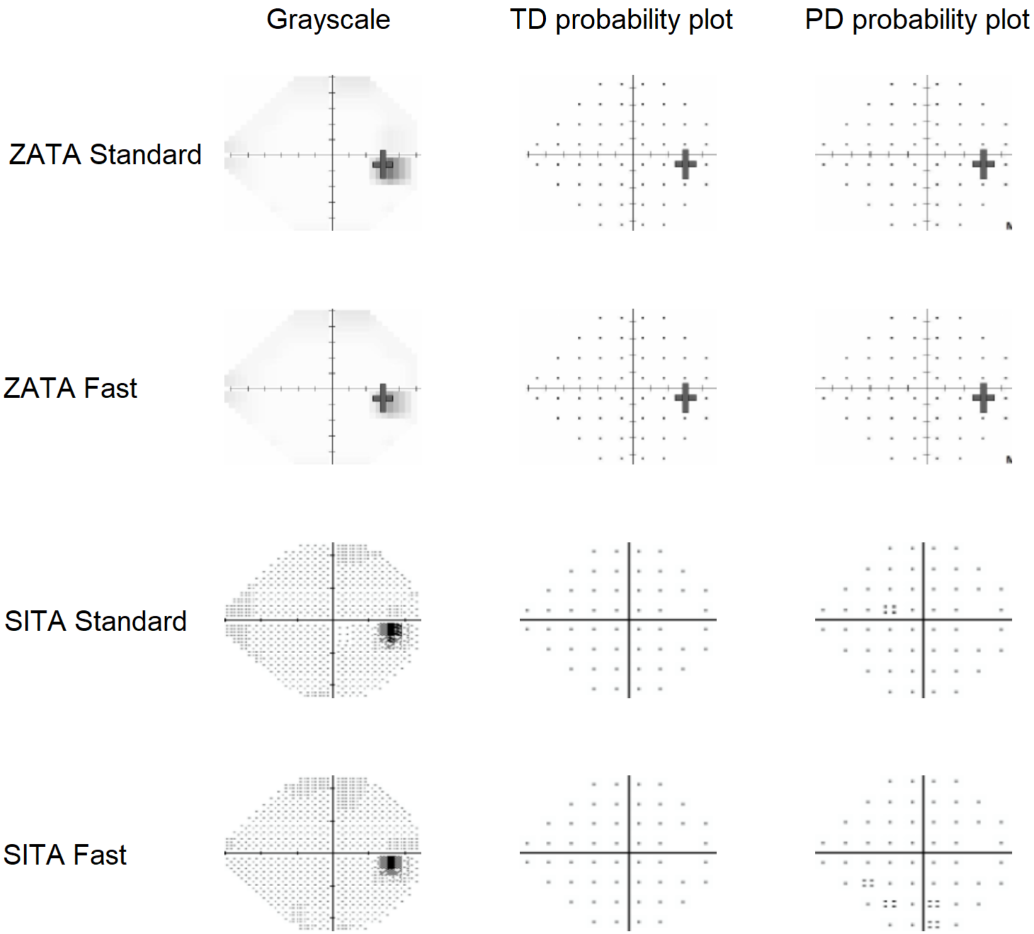

Participant:6

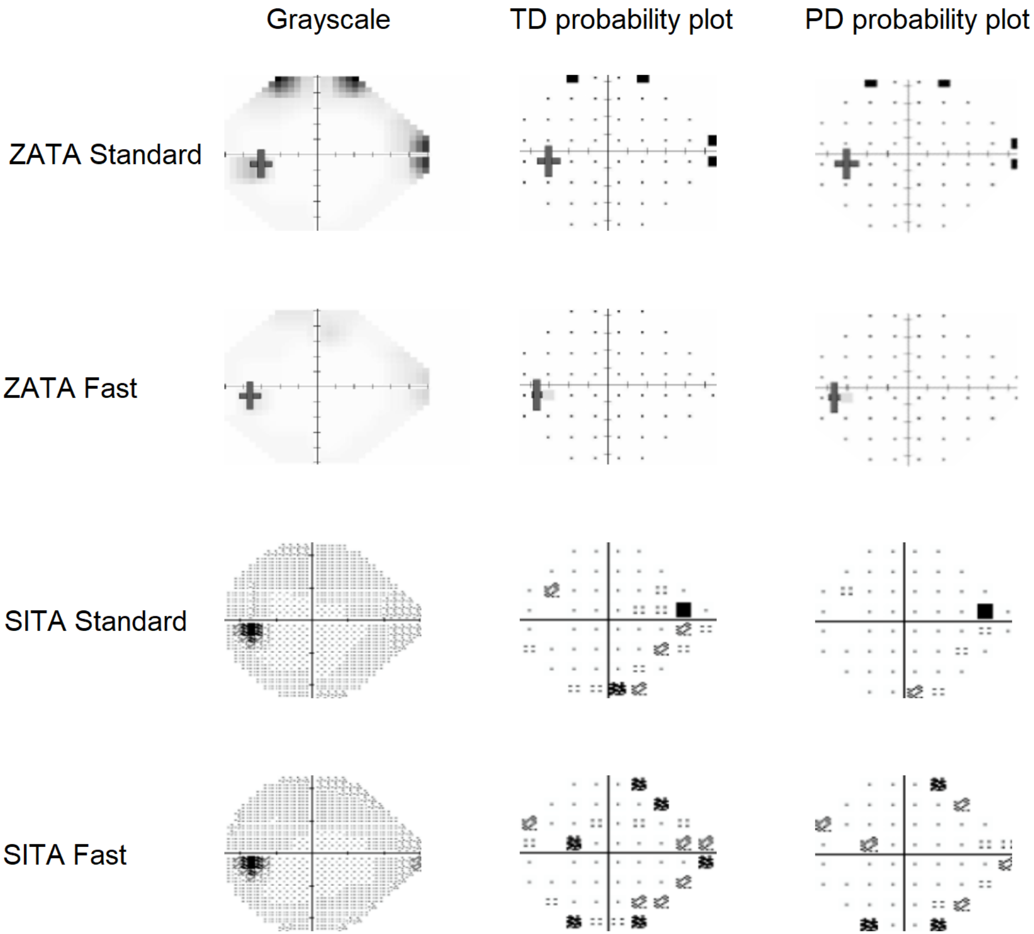

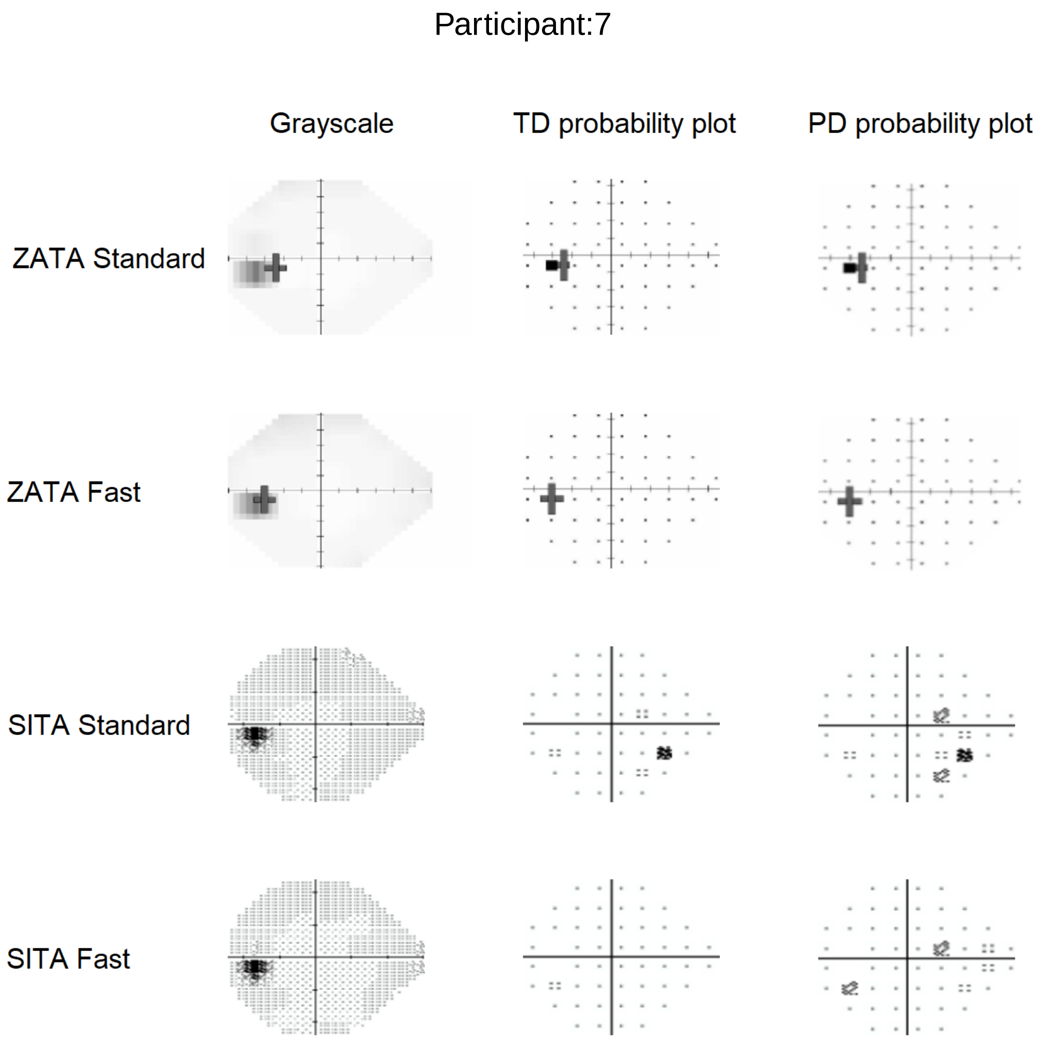
Participant:7
Grayscale
TD probability plot
PD probability plot
ZATA Standard
ZATA Fast
SITA Standard
SITA Fast

Participant:8

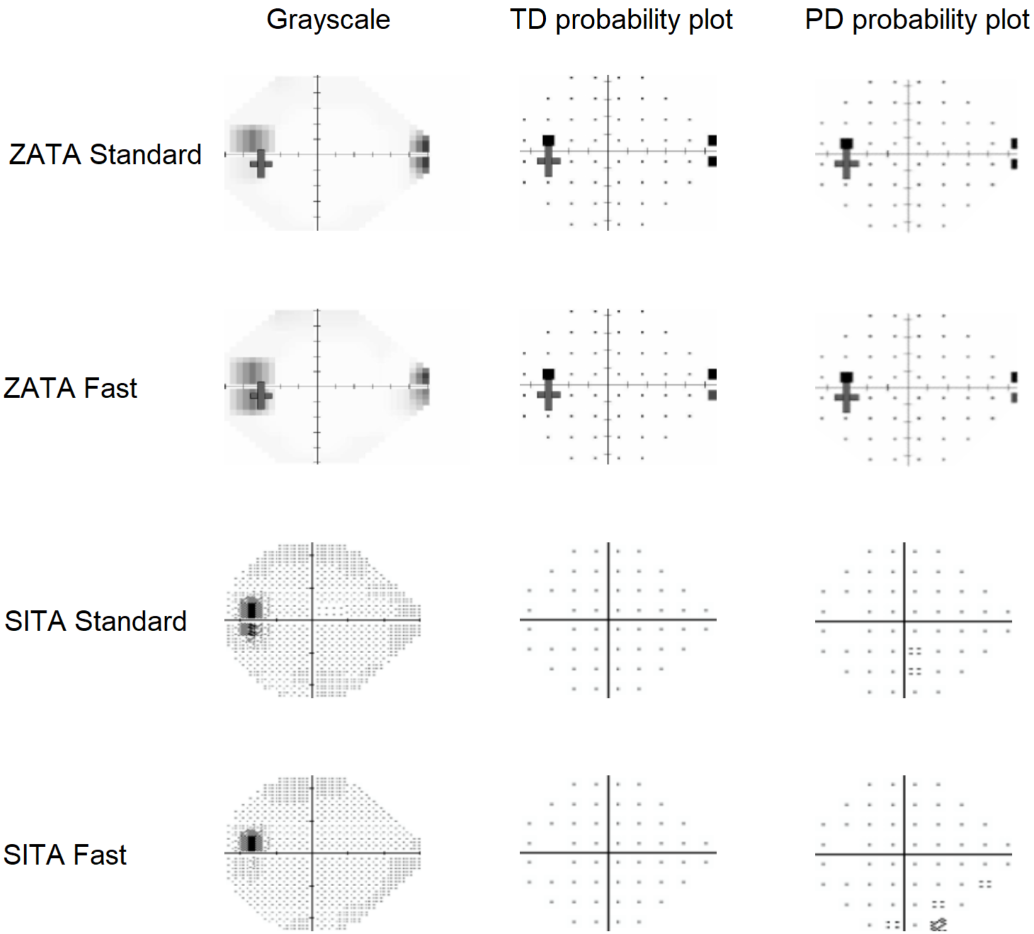

Participant:9

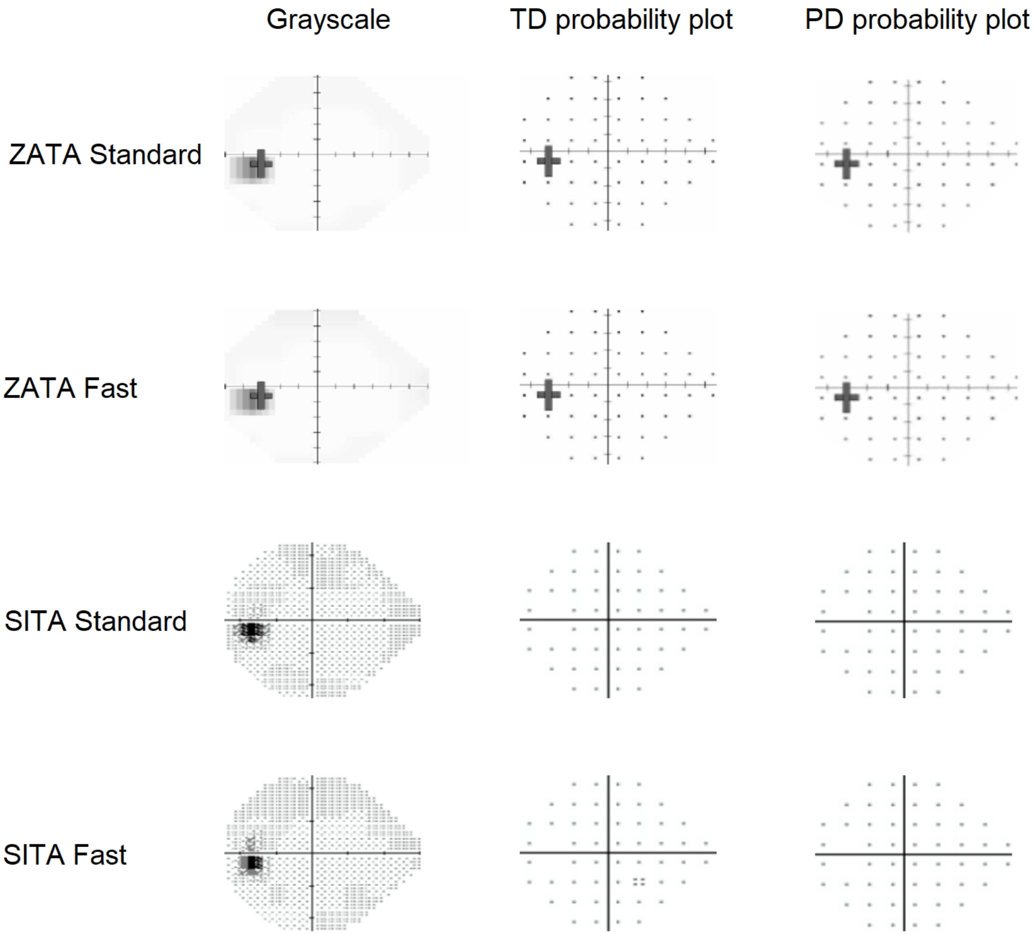

### Participant:10

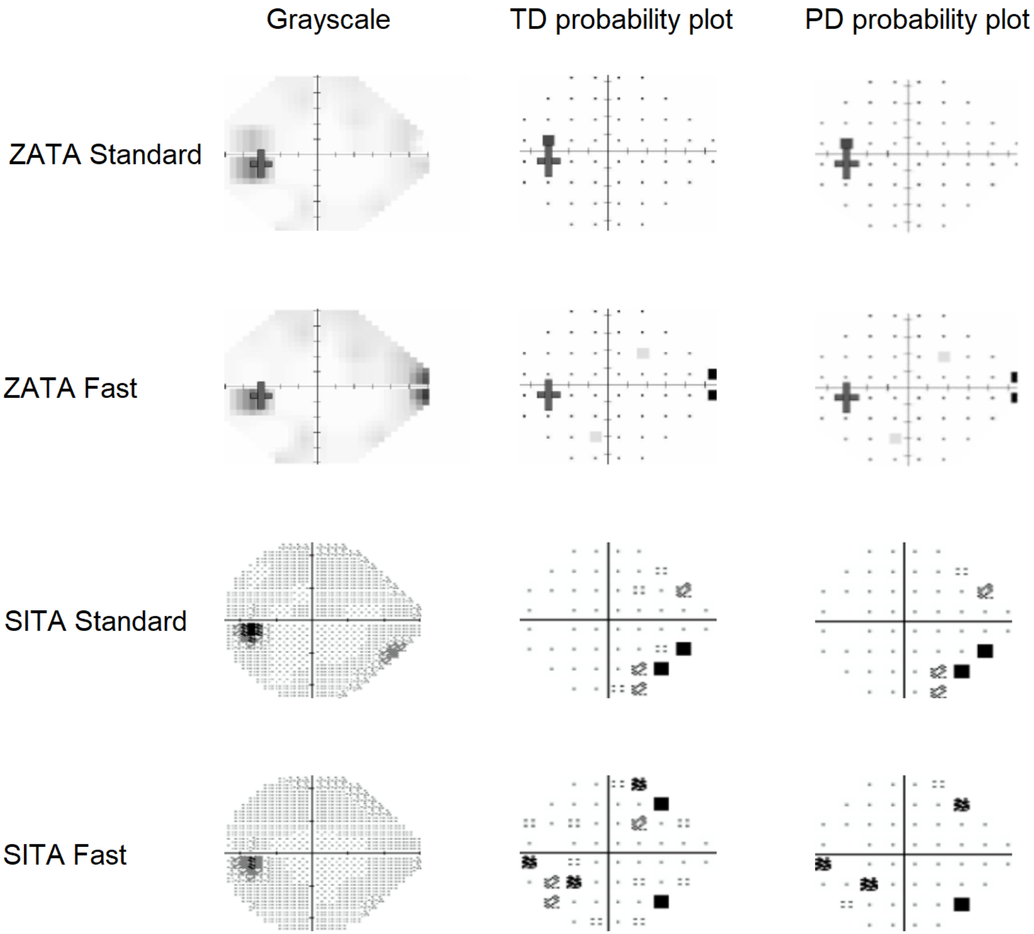

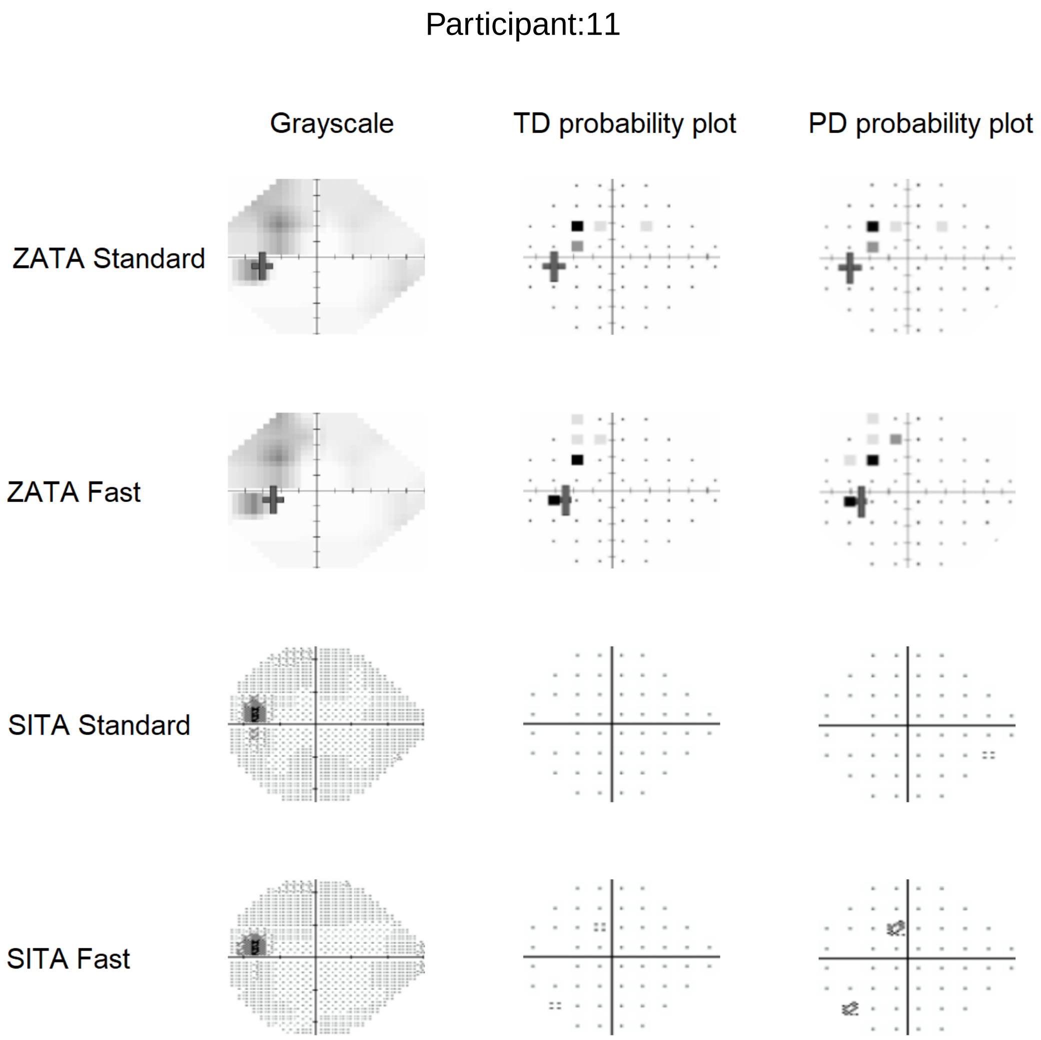
Participant:11
Grayscale
TD probability plot
PD probability plot
ZATA Standard
ZATA Fast
SITA Standard
SITA Fast

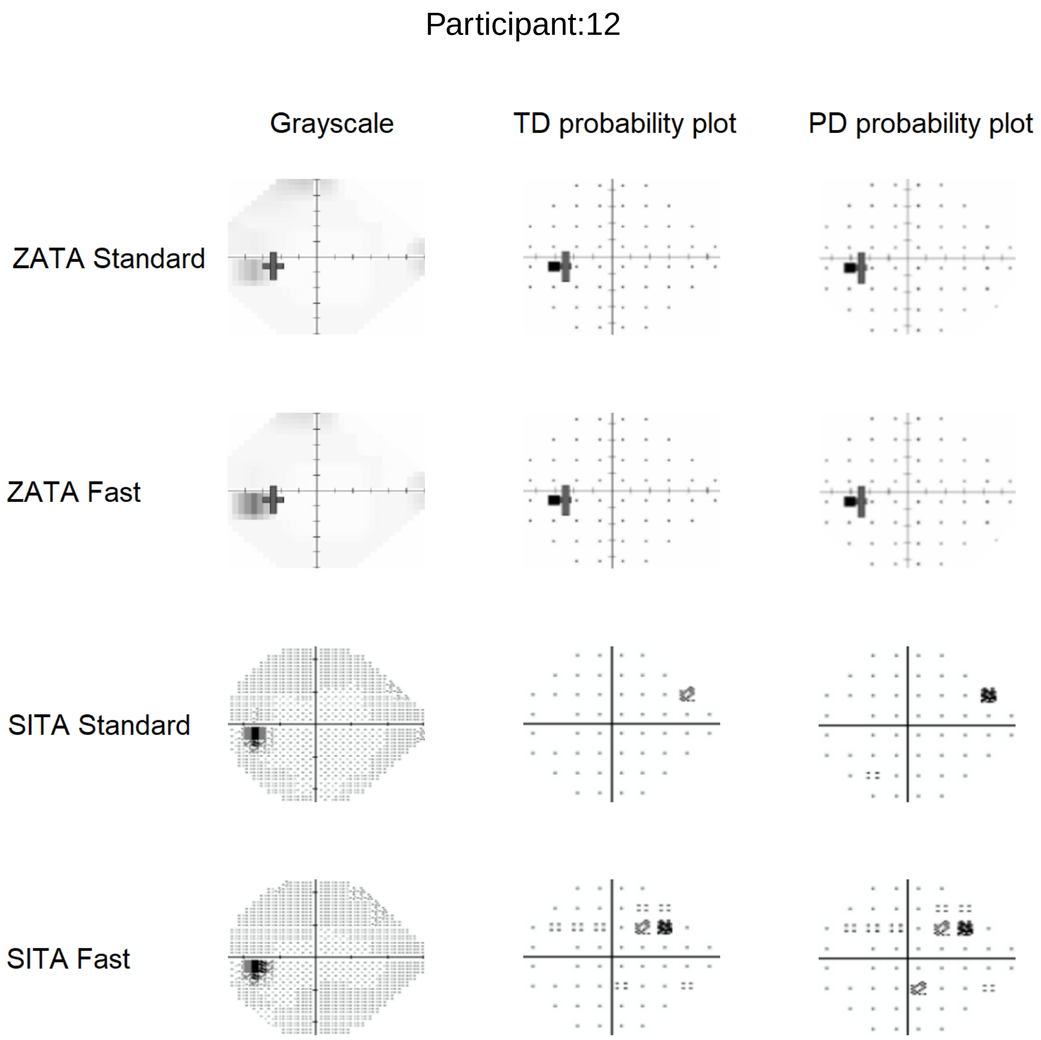
Participant:12
Grayscale
TD probability plot
PD probability plot
ZATA Standard
ZATA Fast
SITA Standard
SITA Fast

Participant:13

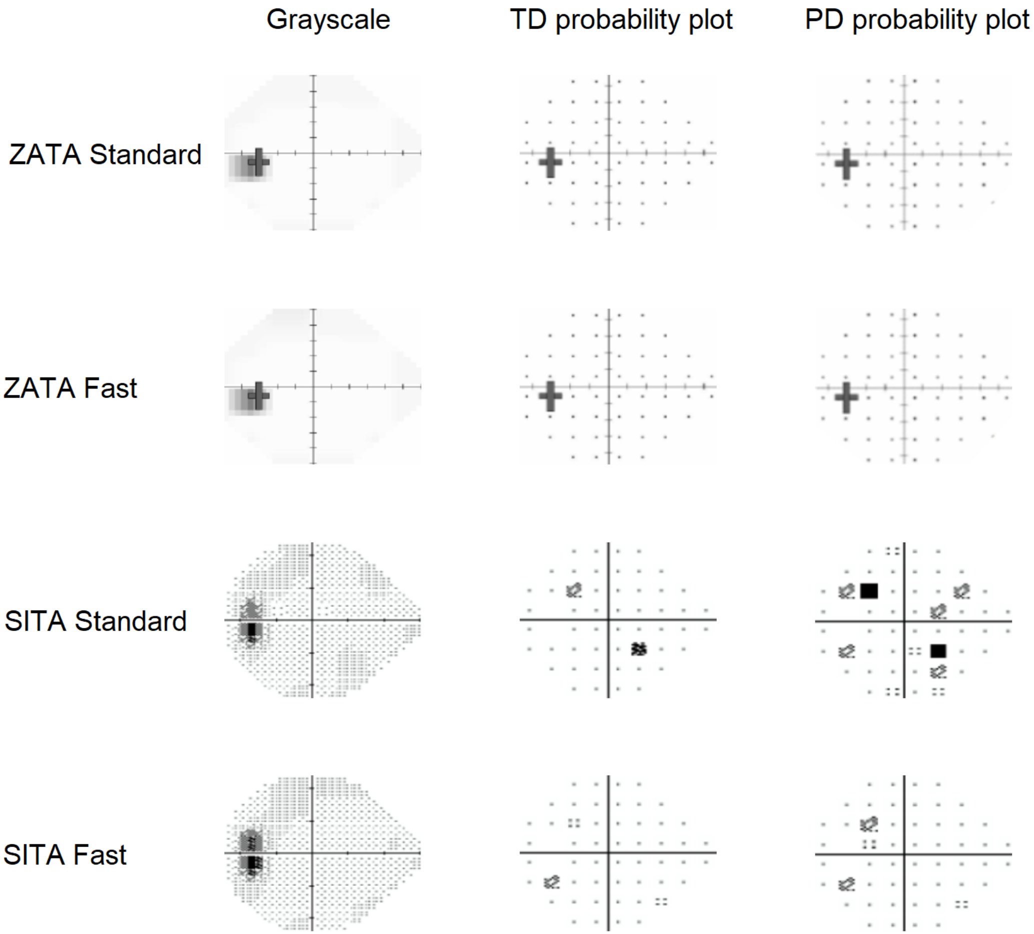

Participant:14

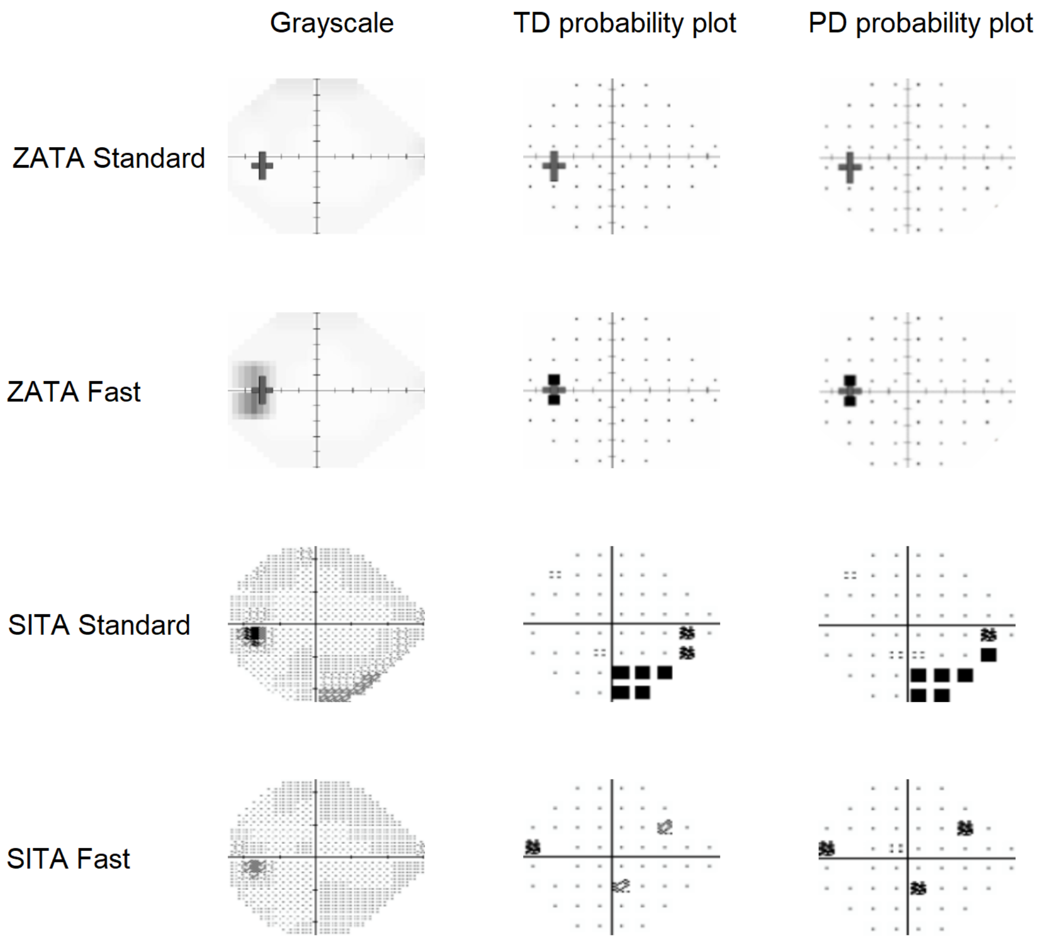

Participant:15

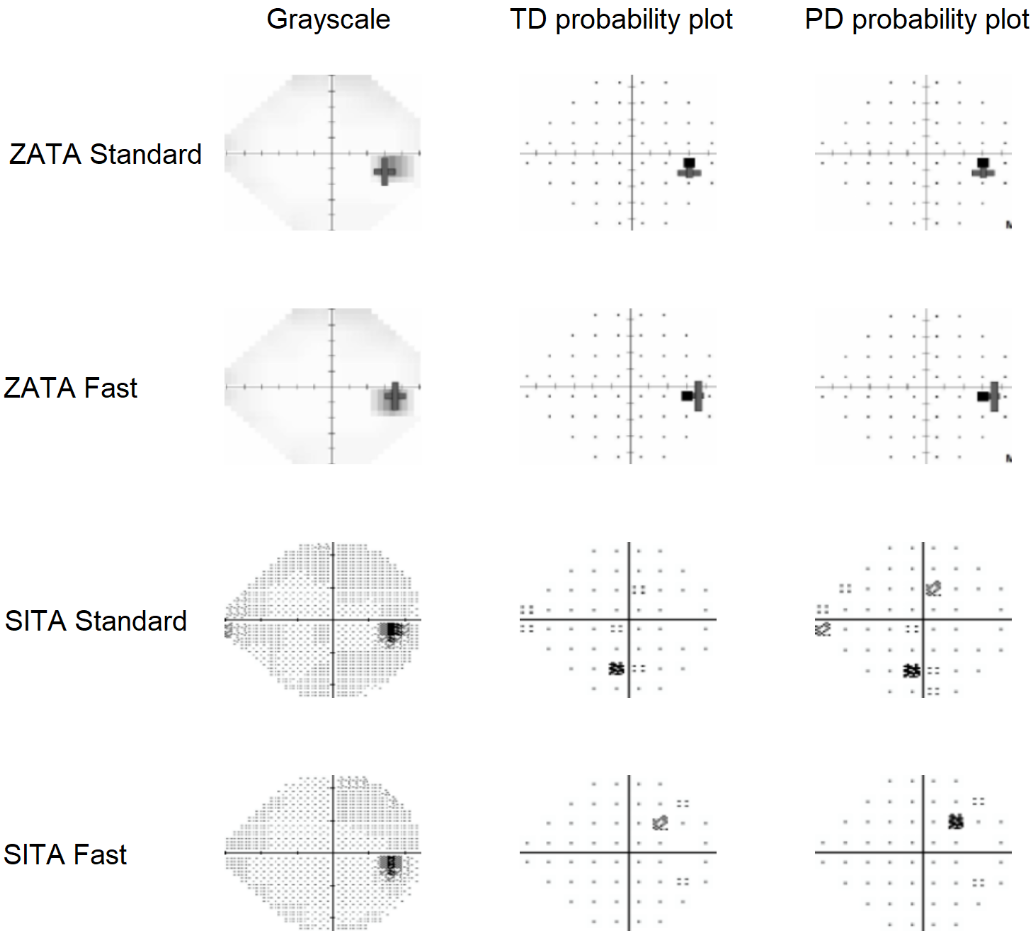

Participant:16

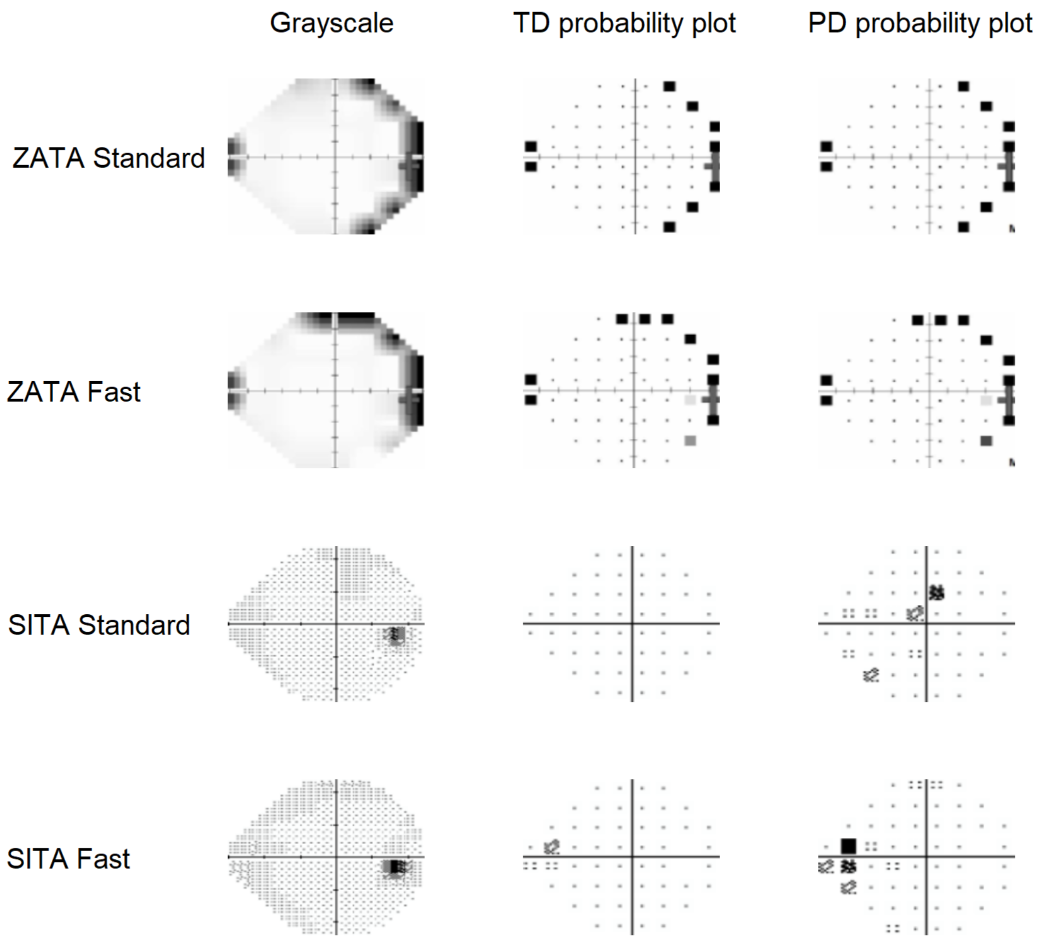

Participant:17

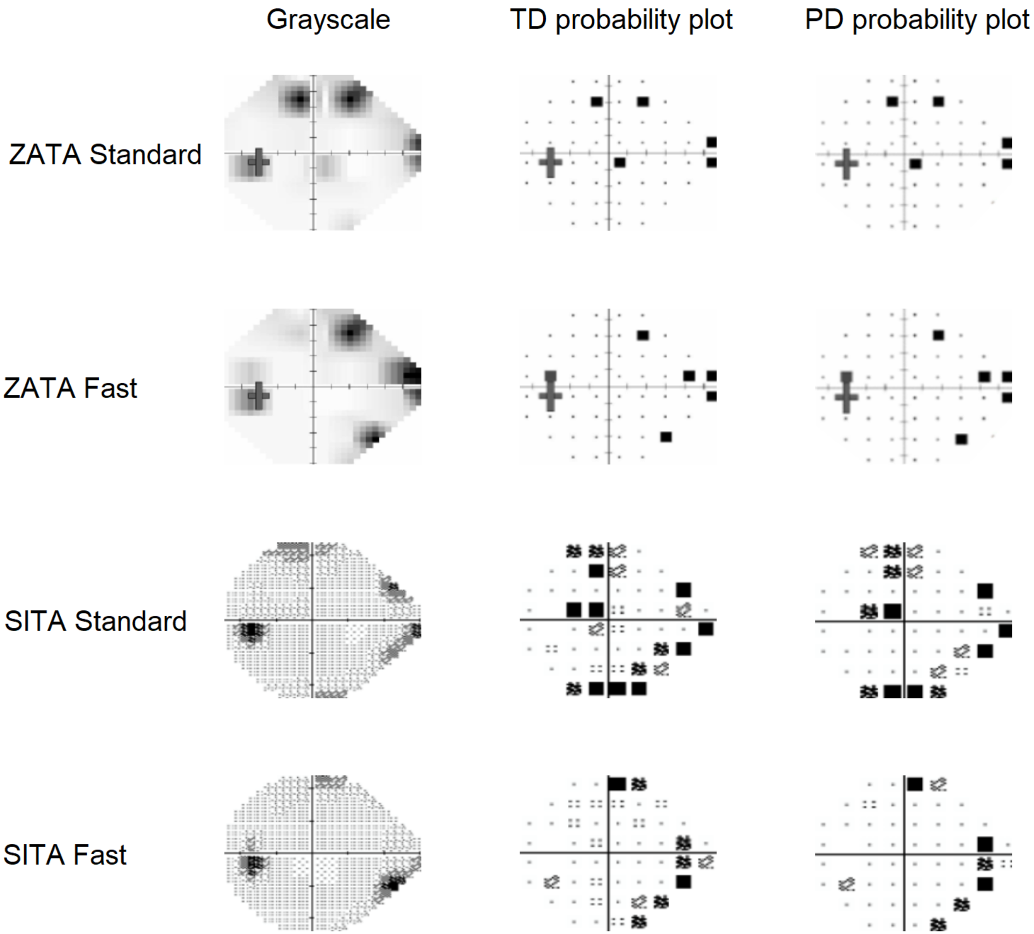

Participant:18

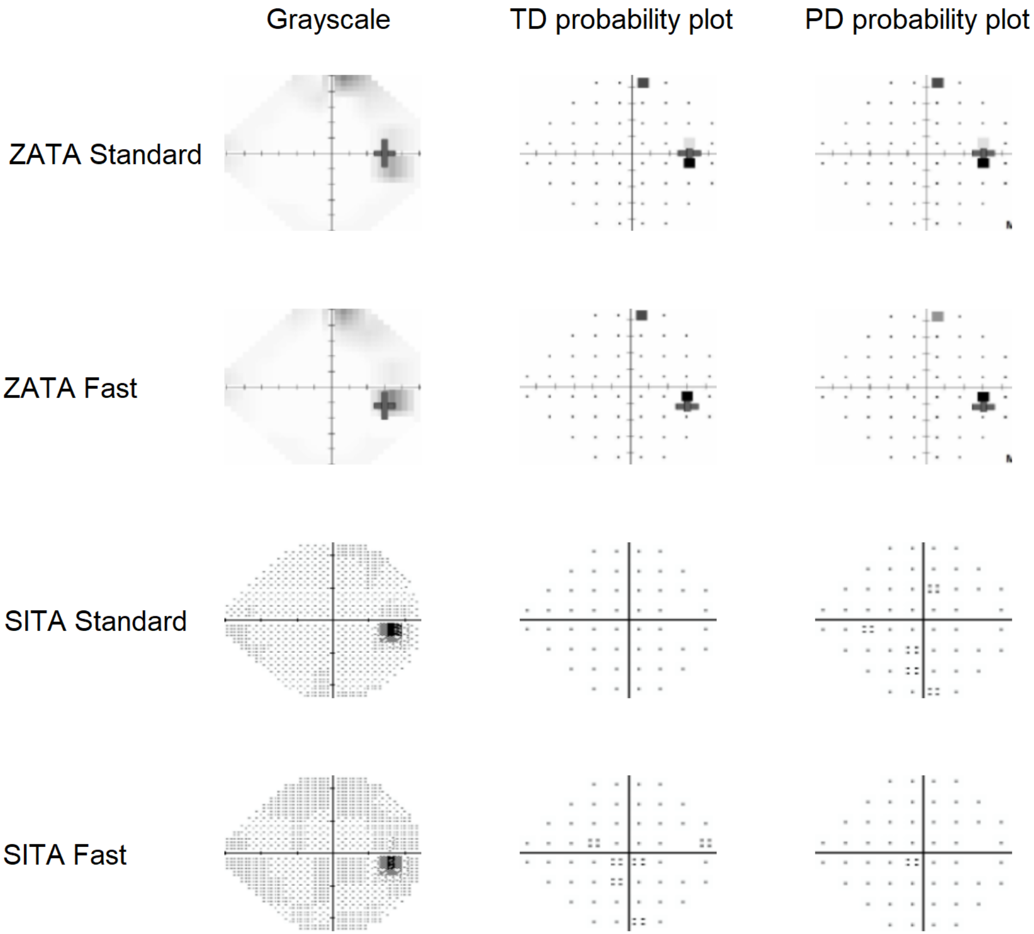

Participant:19

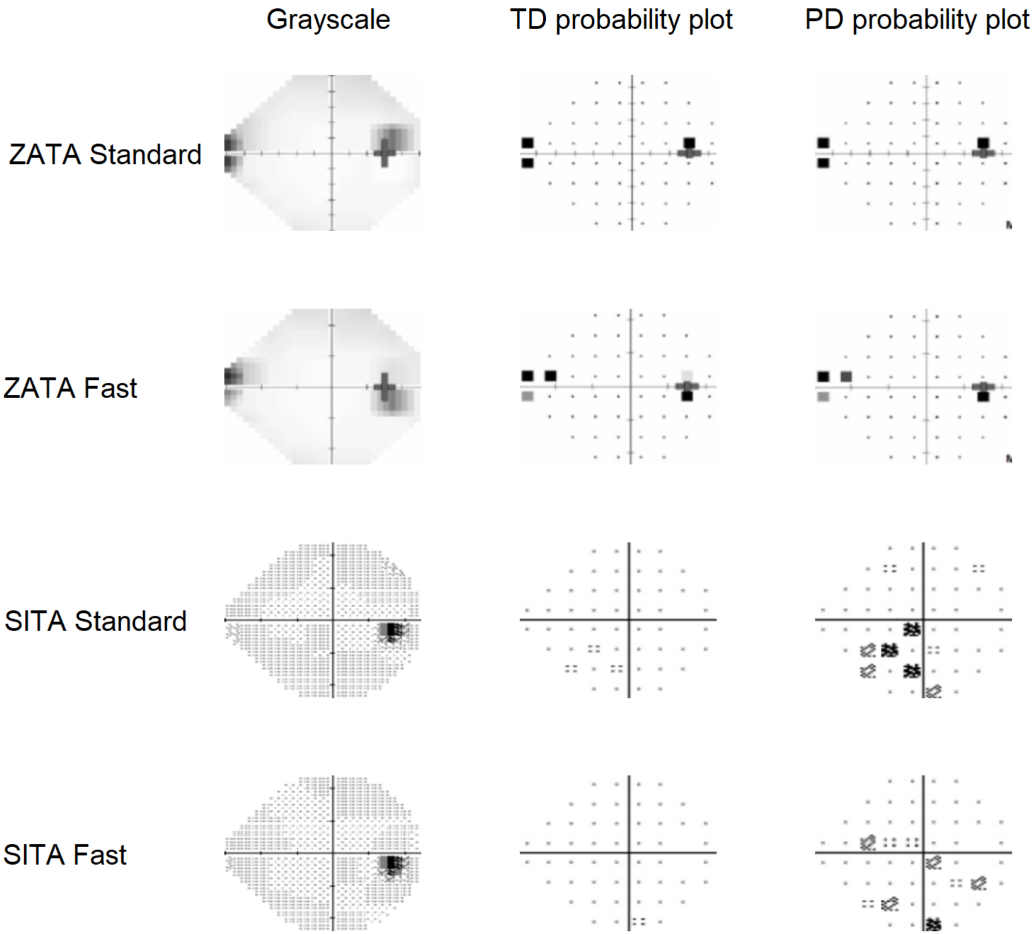

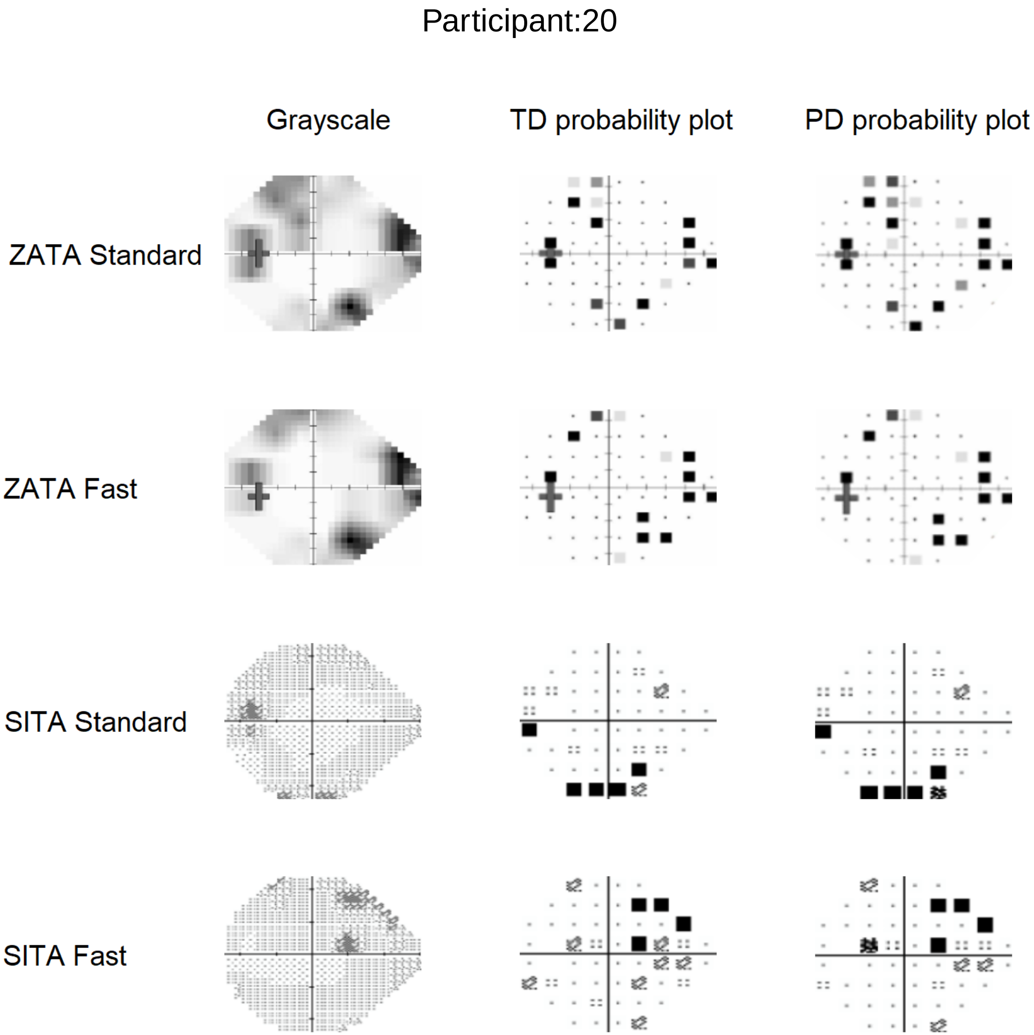

Participant:20
Grayscale
TD probability plot
PD probability plot
ZATA Standard
ZATA Fast
SITA Standard
SITA Fast

Participant:21

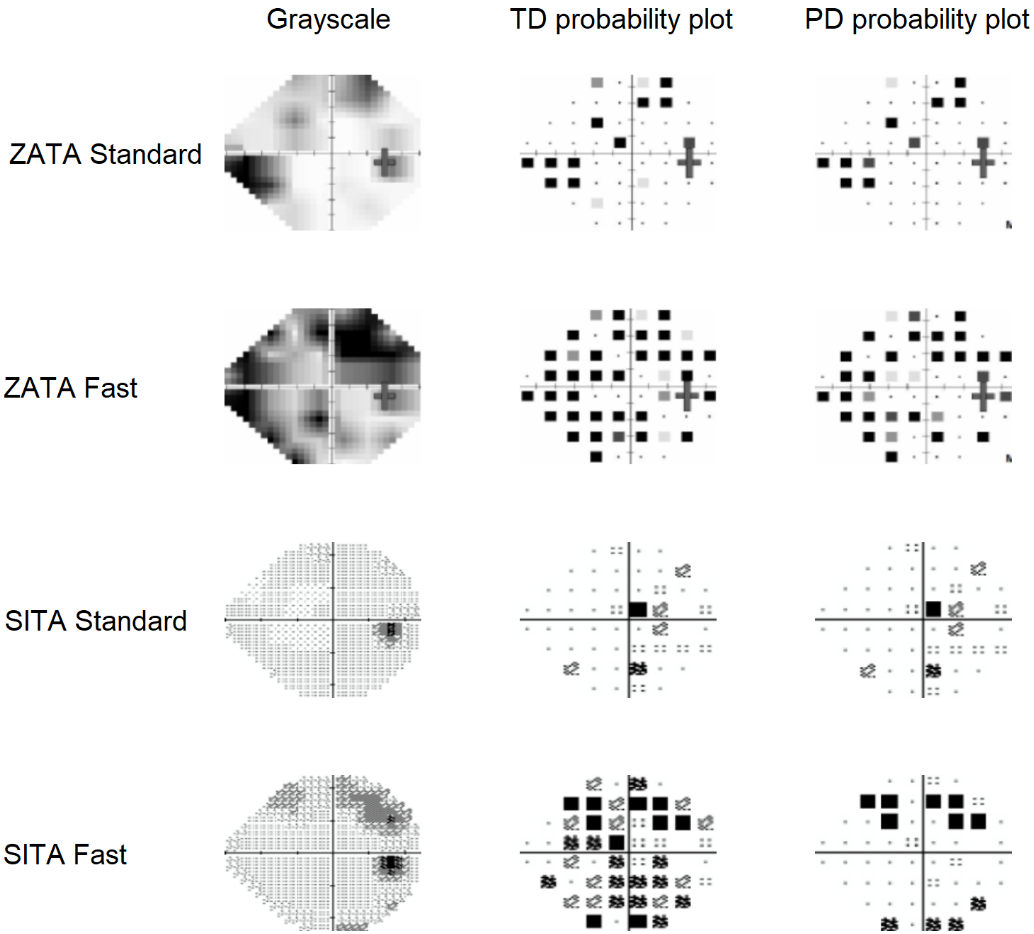

Participant:22

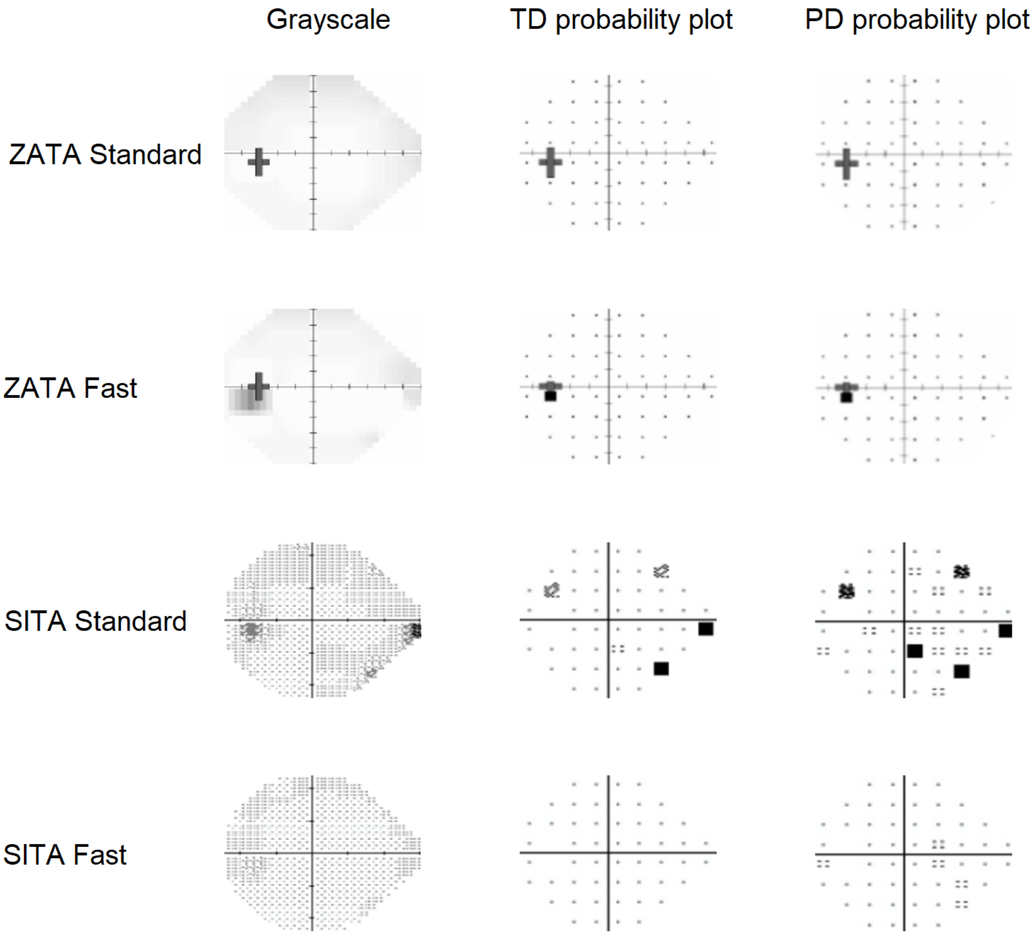

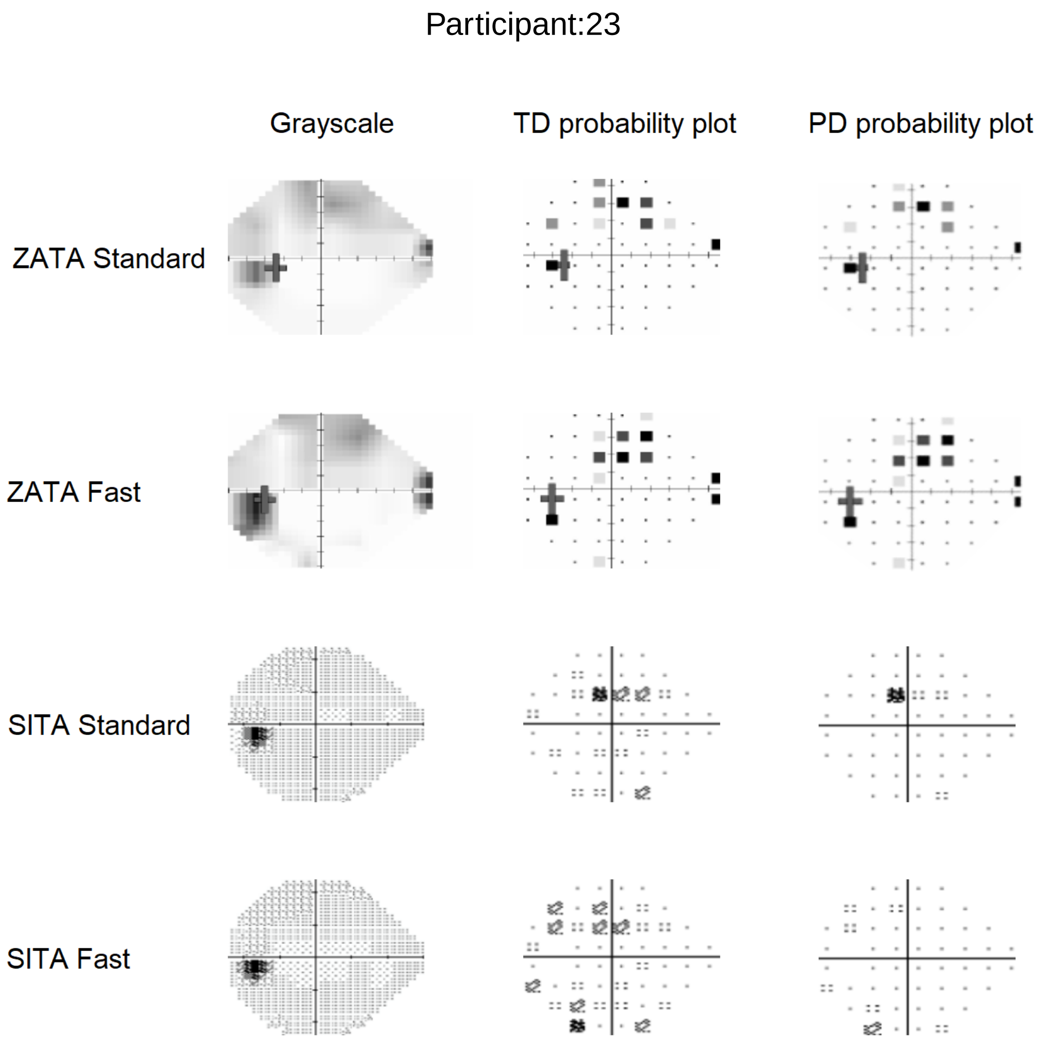

Participant:23
Grayscale
TD probability plot
PD probability plot
ZATA Standard
ZATA Fast
SITA Standard
SITA Fast

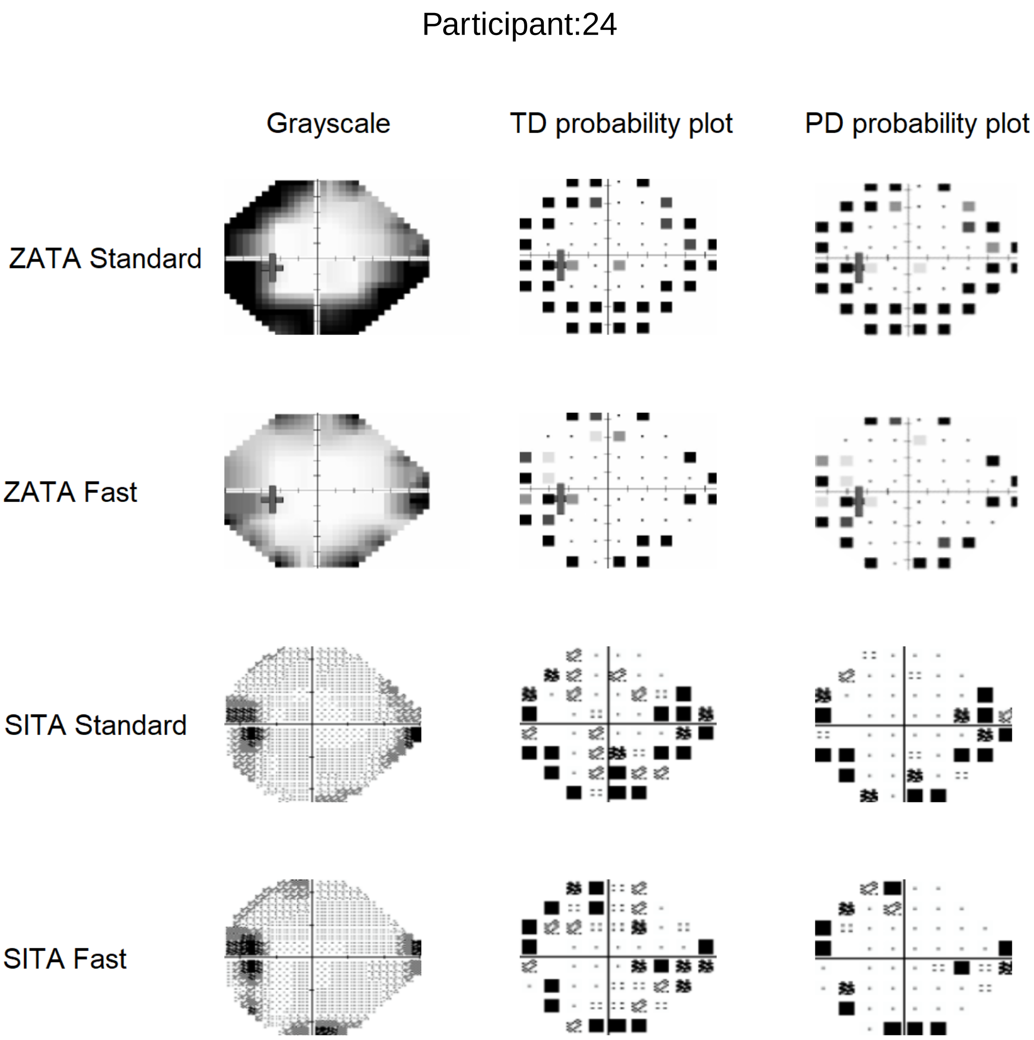
Participant:24
Grayscale
TD probability plot
PD probability plot
ZATA Standard
ZATA Fast
SITA Standard
SITA Fast

Participant:25

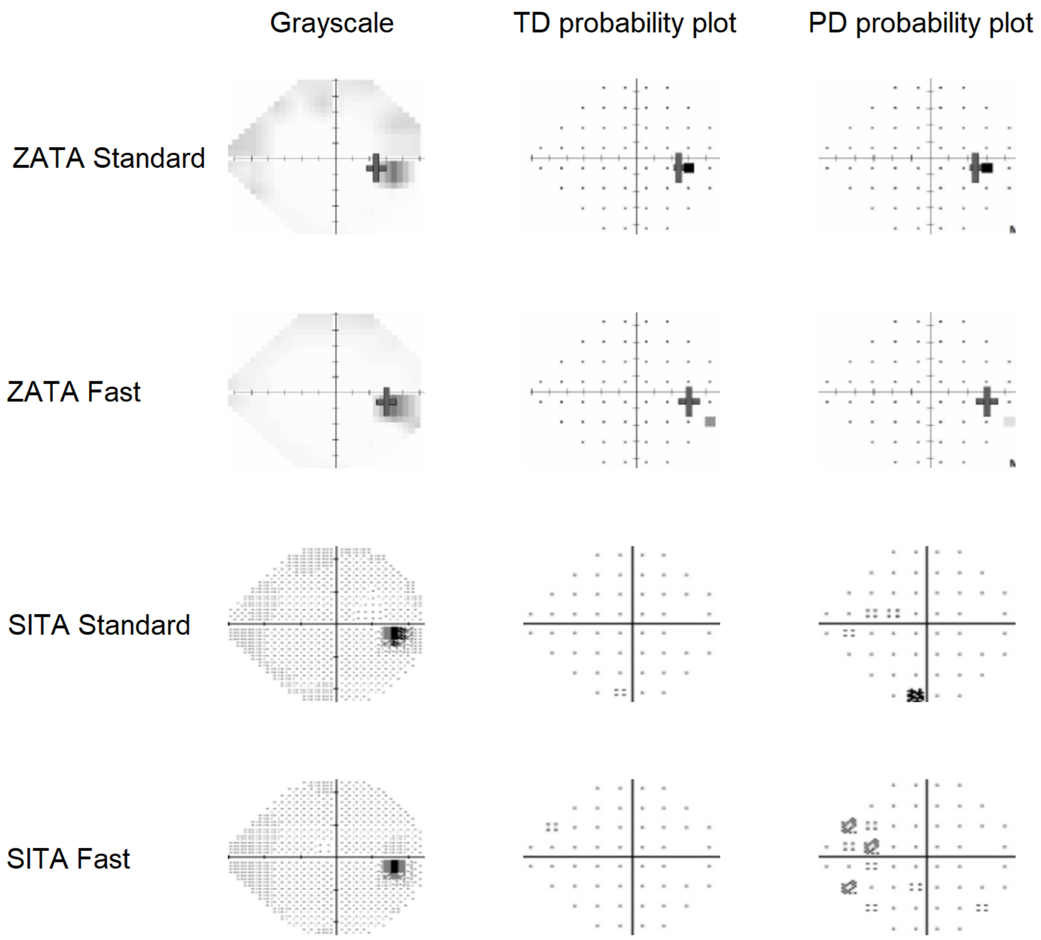

Participant:26

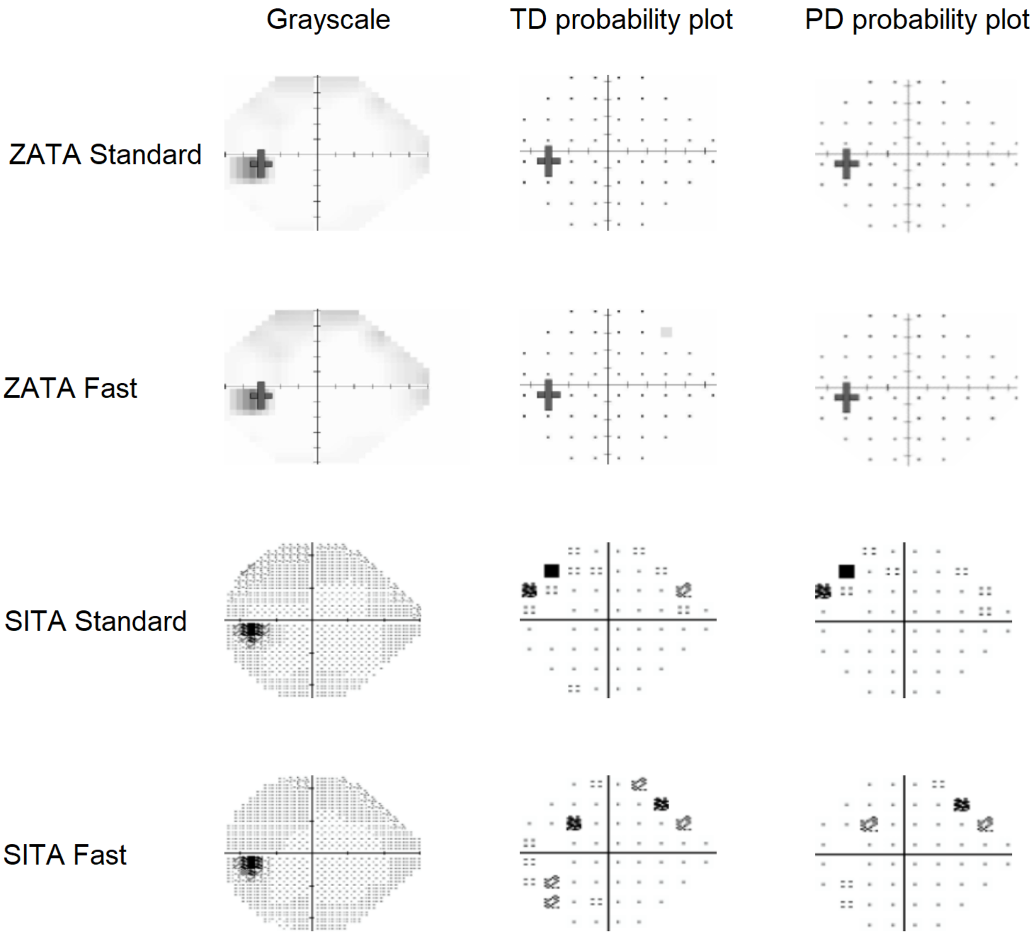

#### Participant:27

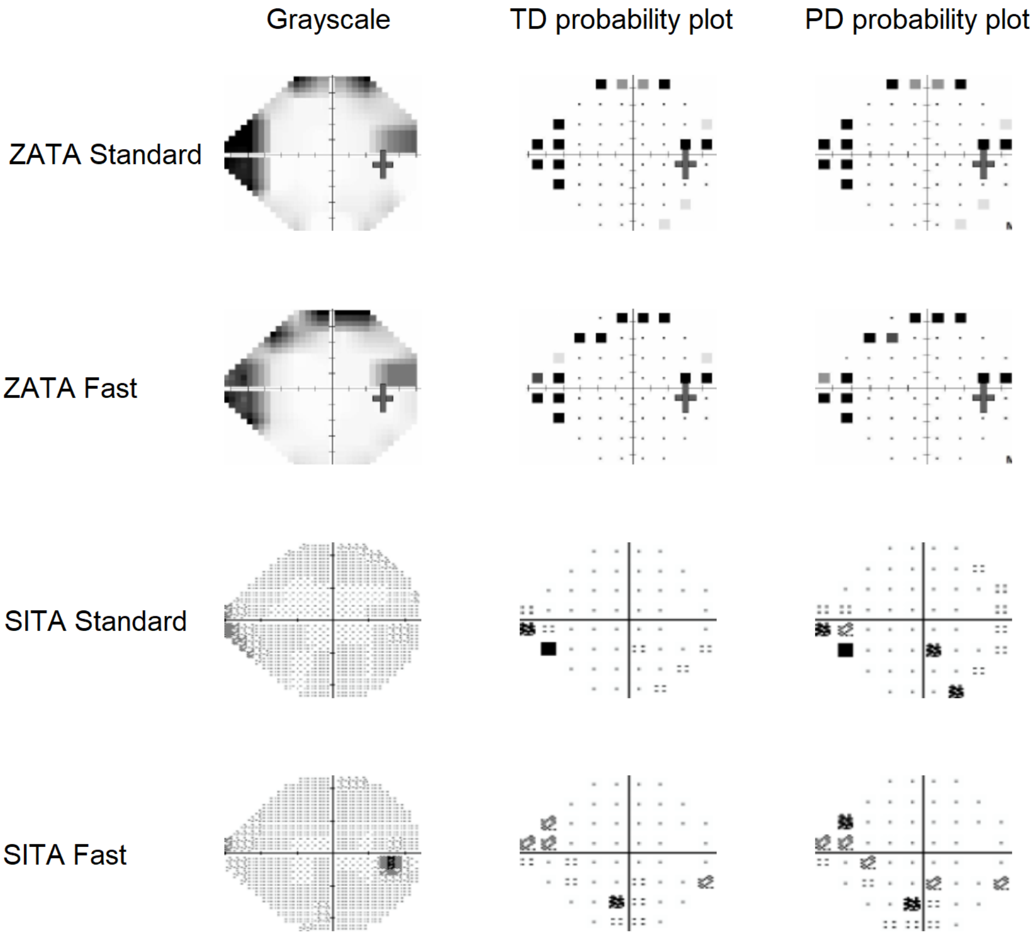

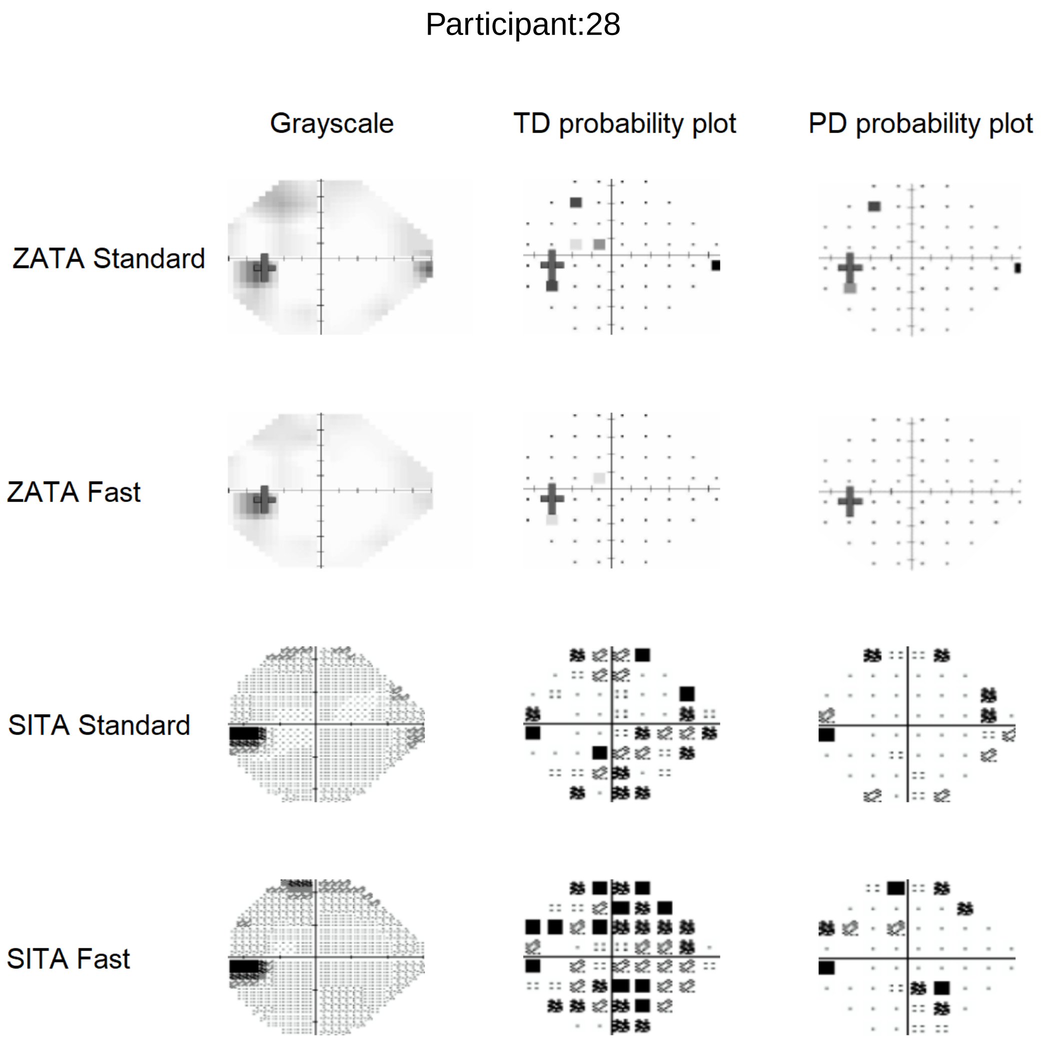
Participant:28
Grayscale
TD probability plot
PD probability plot
ZATA Standard
ZATA Fast
SITA Standard
SITA Fast

Participant:29

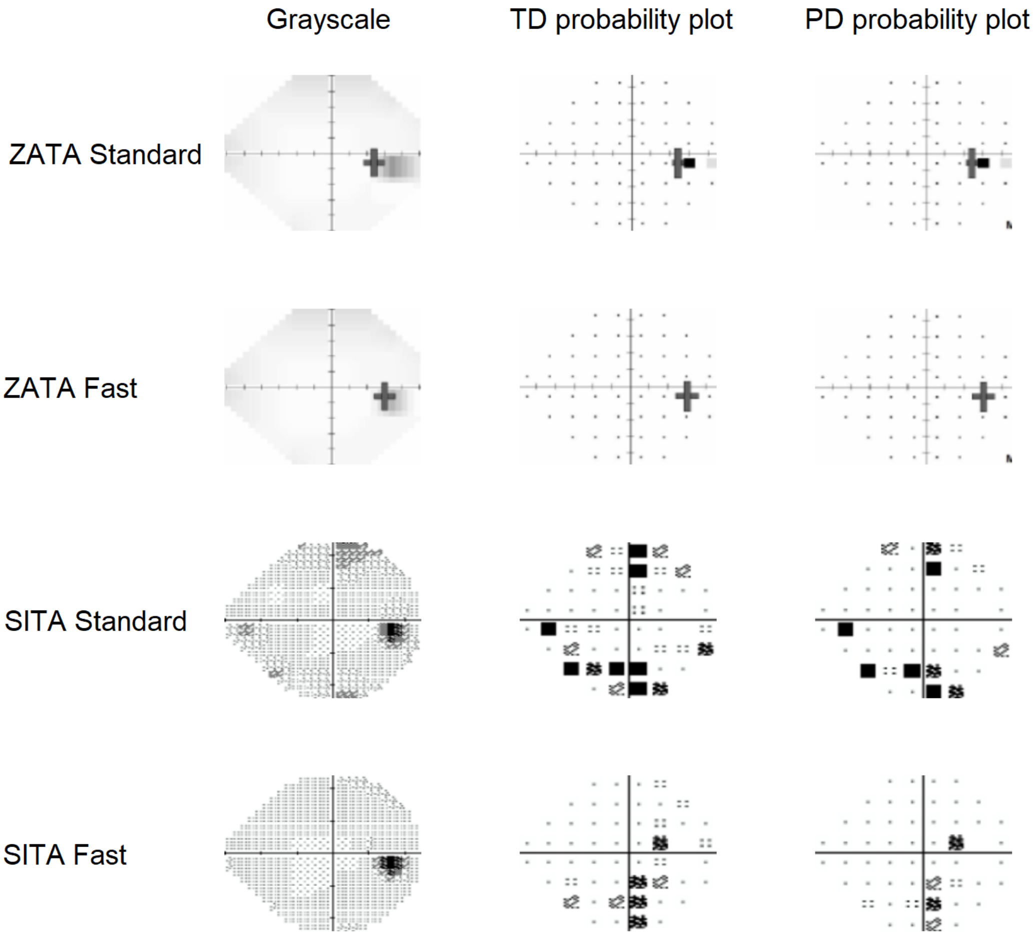

Participant:30

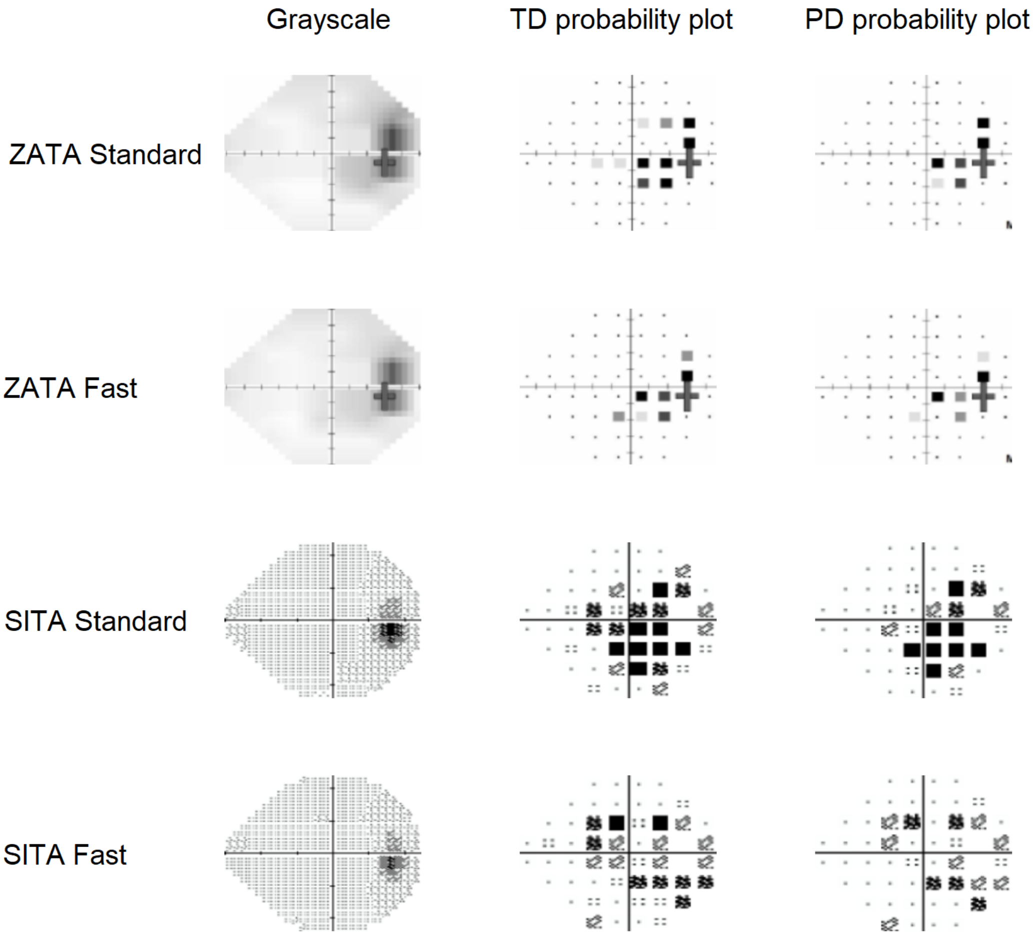

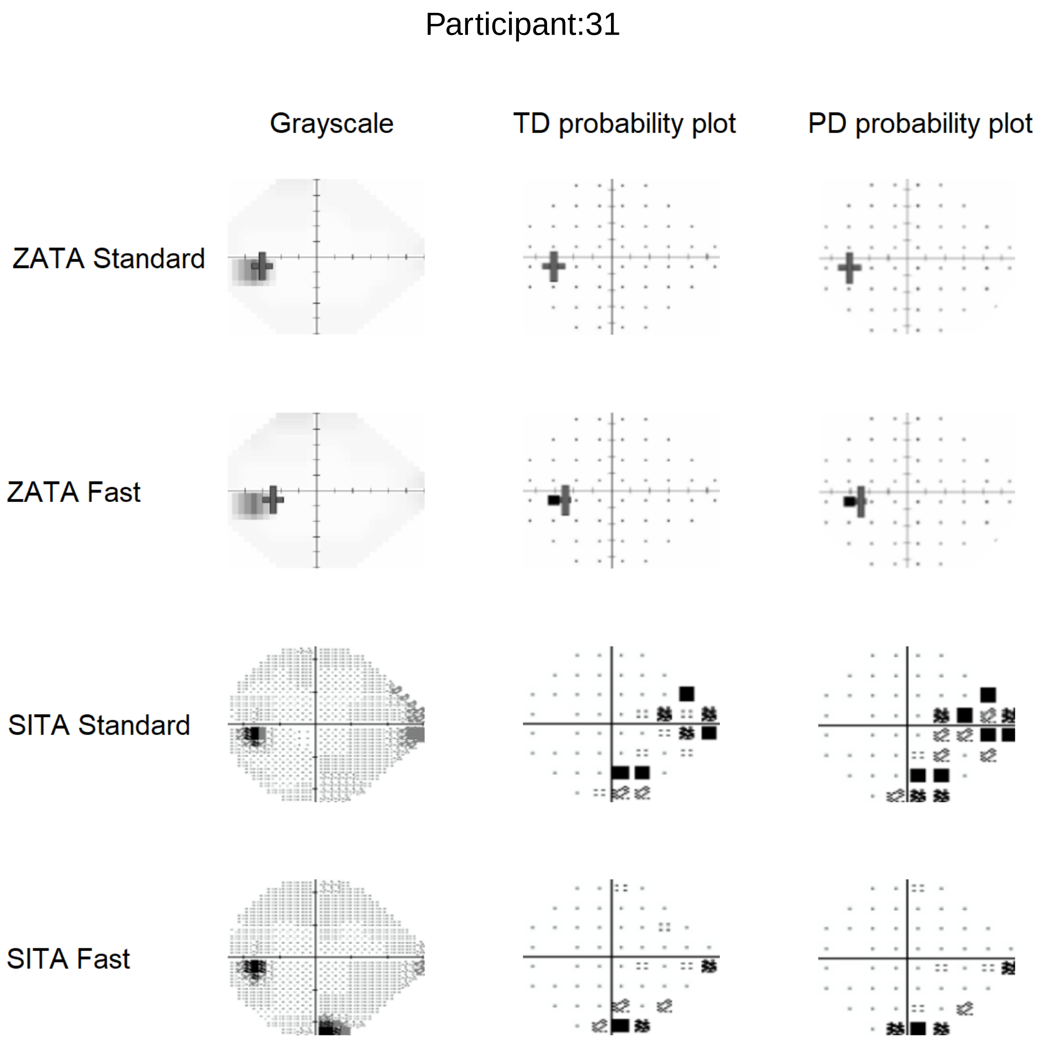
Participant:31
Grayscale
TD probability plot
PD probability plot
ZATA Standard
ZATA Fast
SITA Standard
SITA Fast

Participant:32

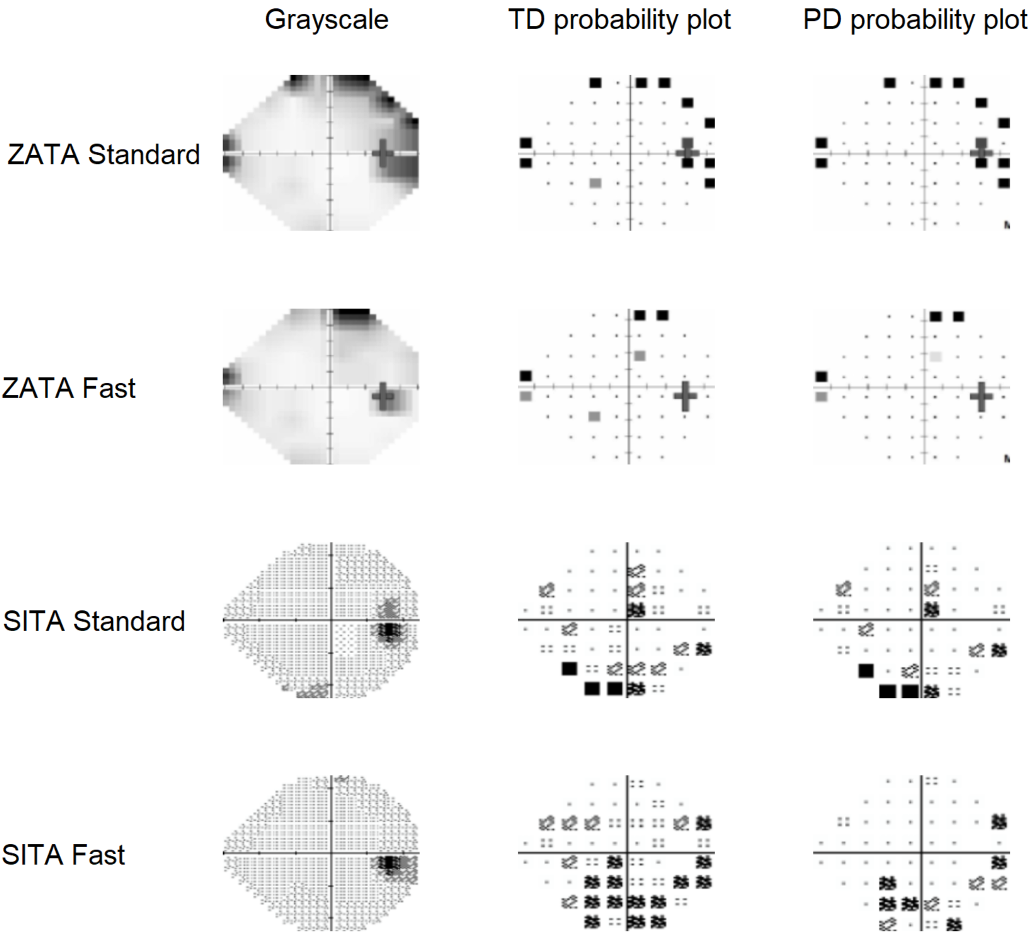

Participant:33

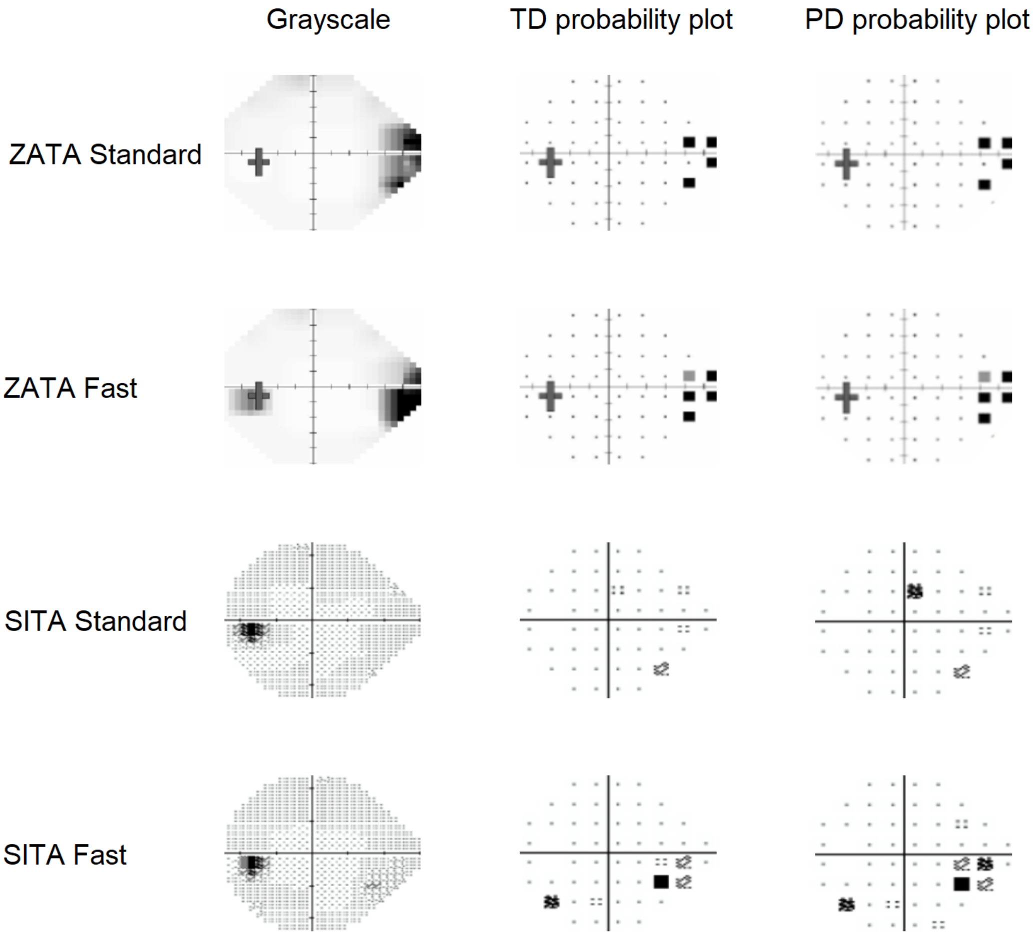

Participant:34

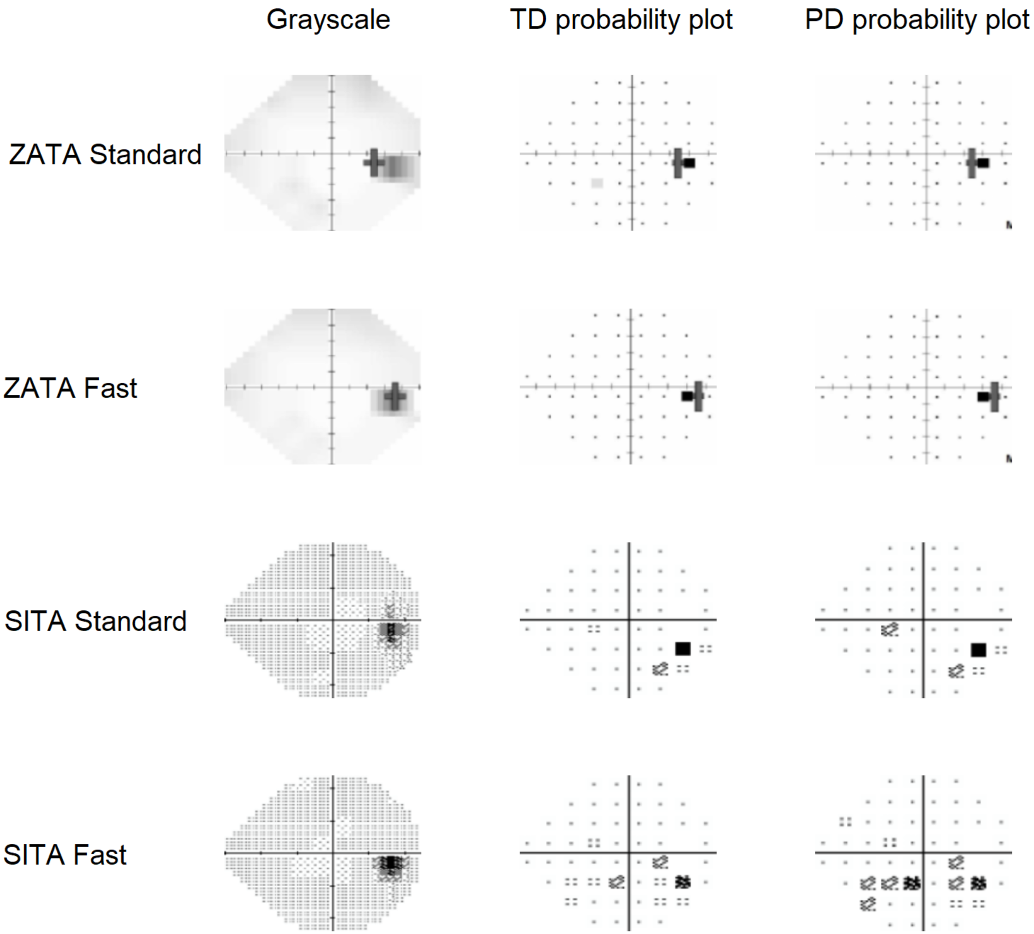

### Participant:35

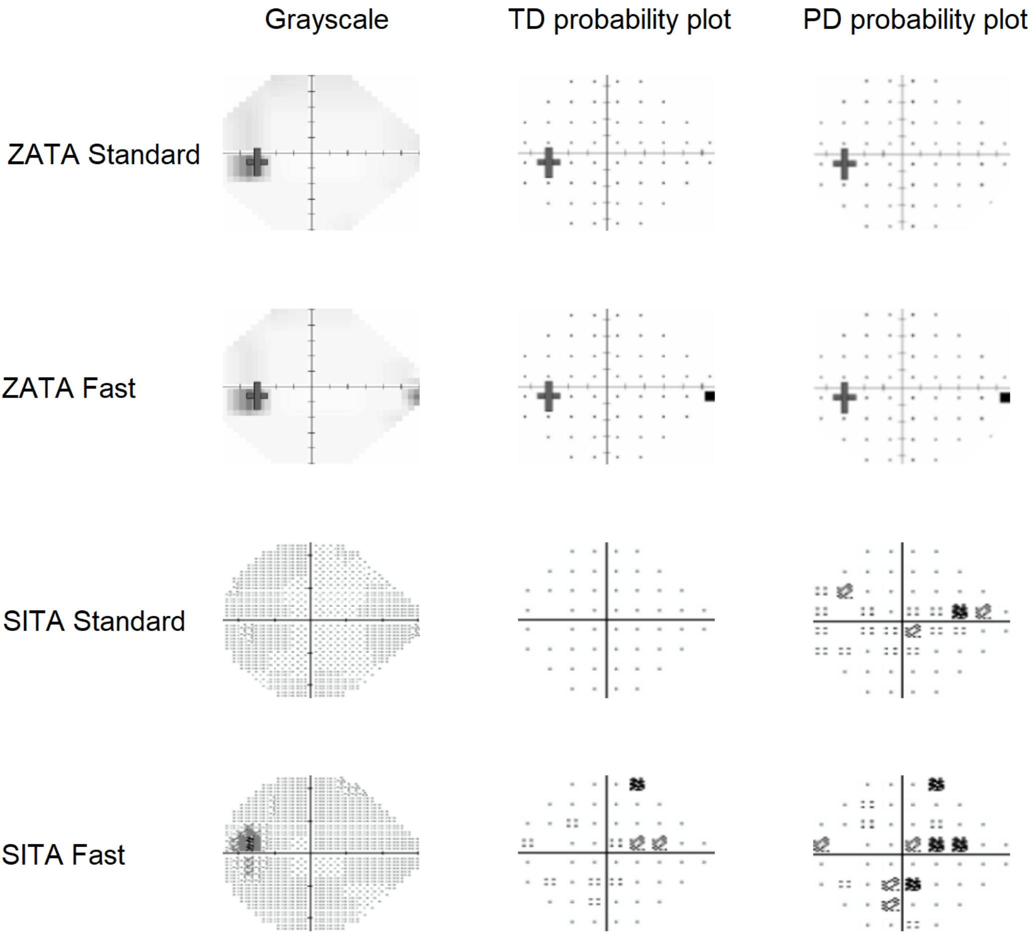

Participant:36

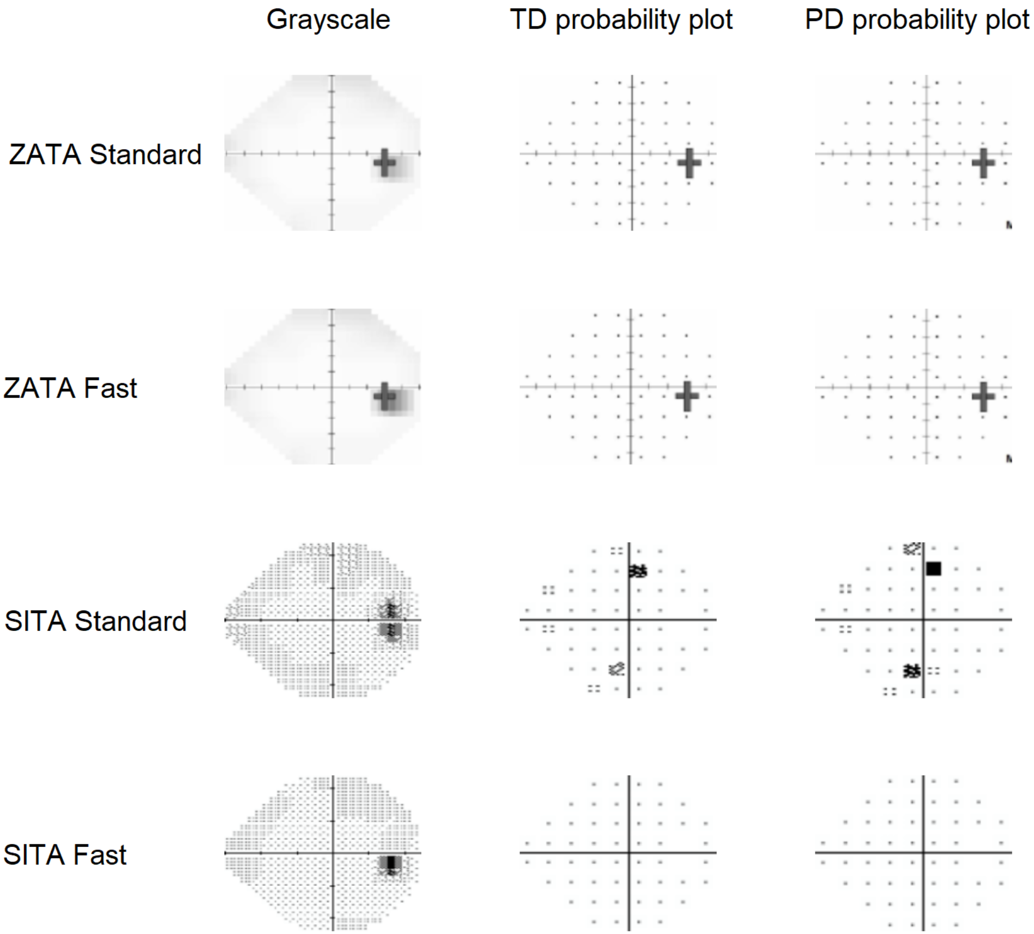

Participant:37

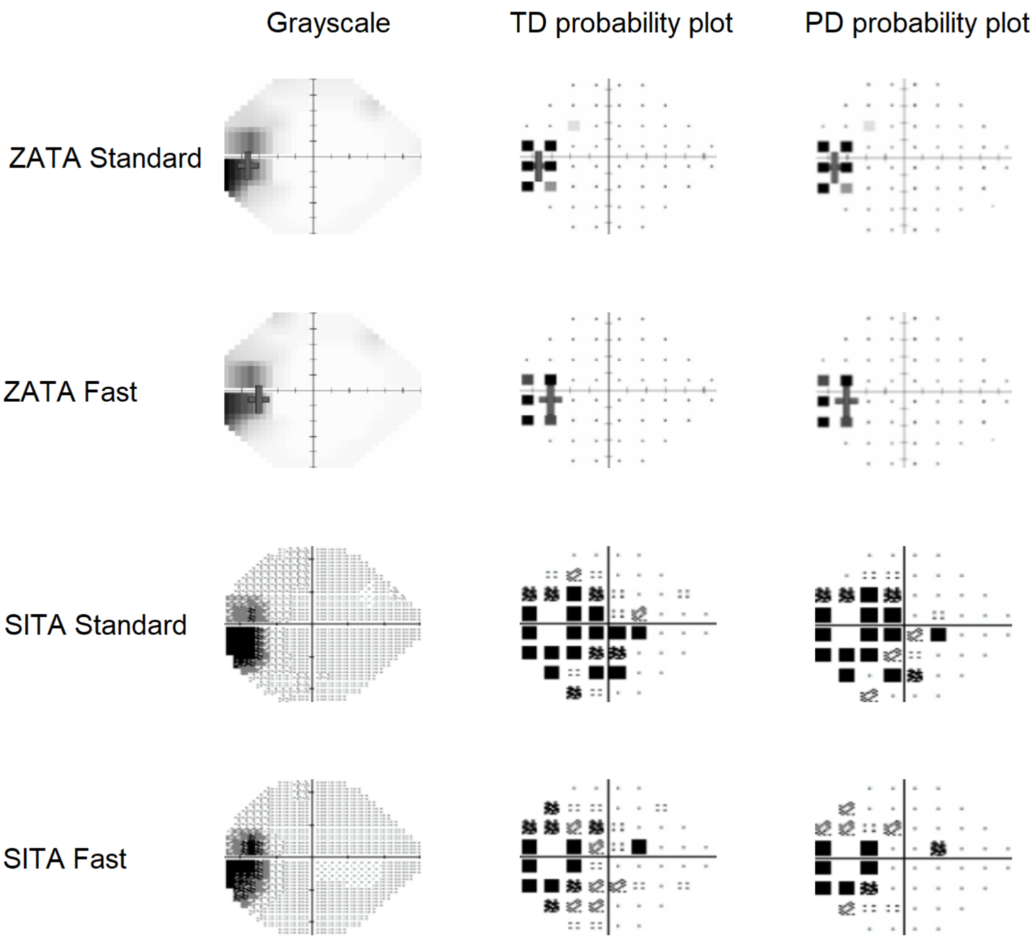

Participant:38

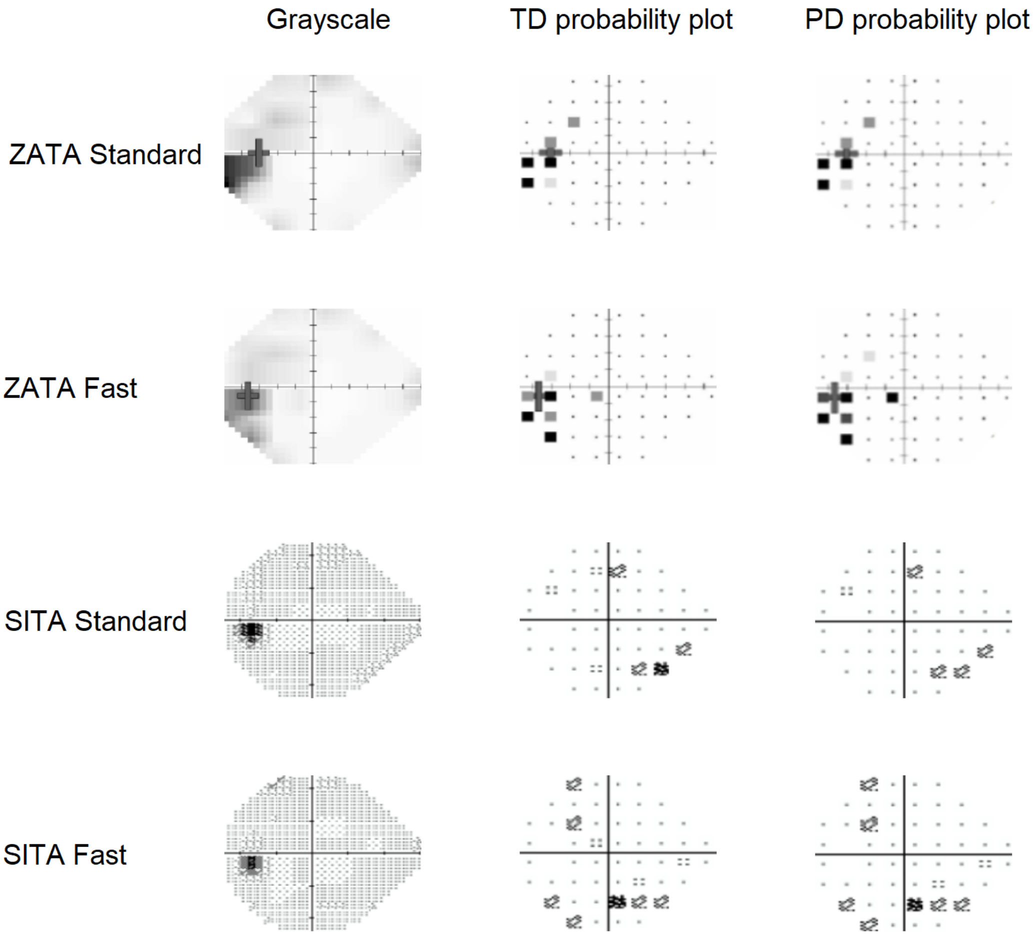

Participant:39

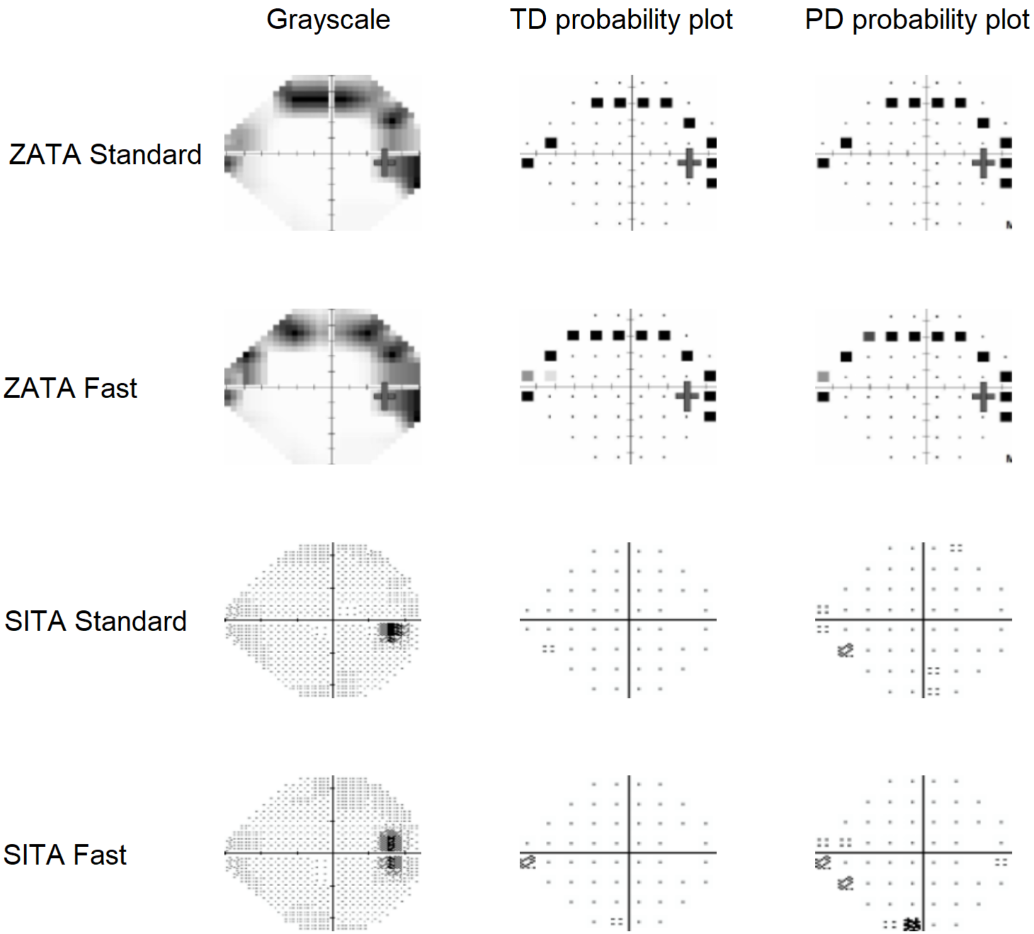

Participant:40

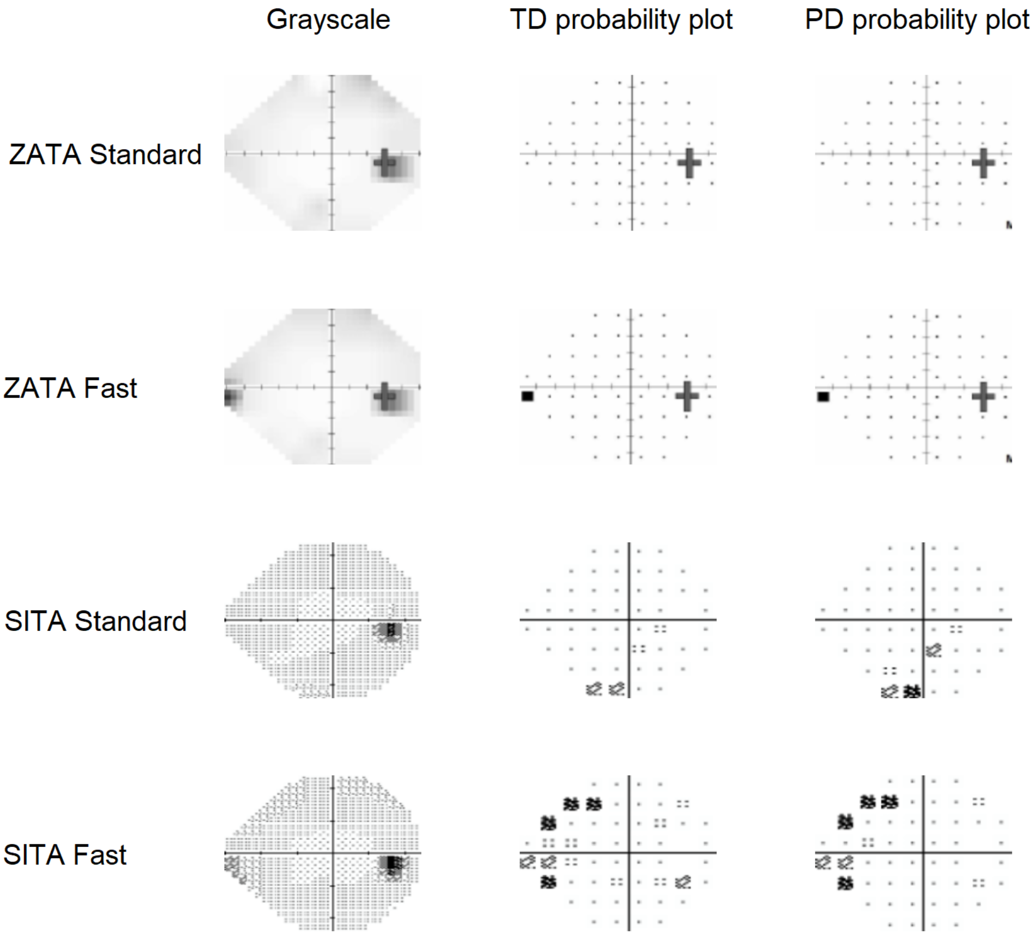

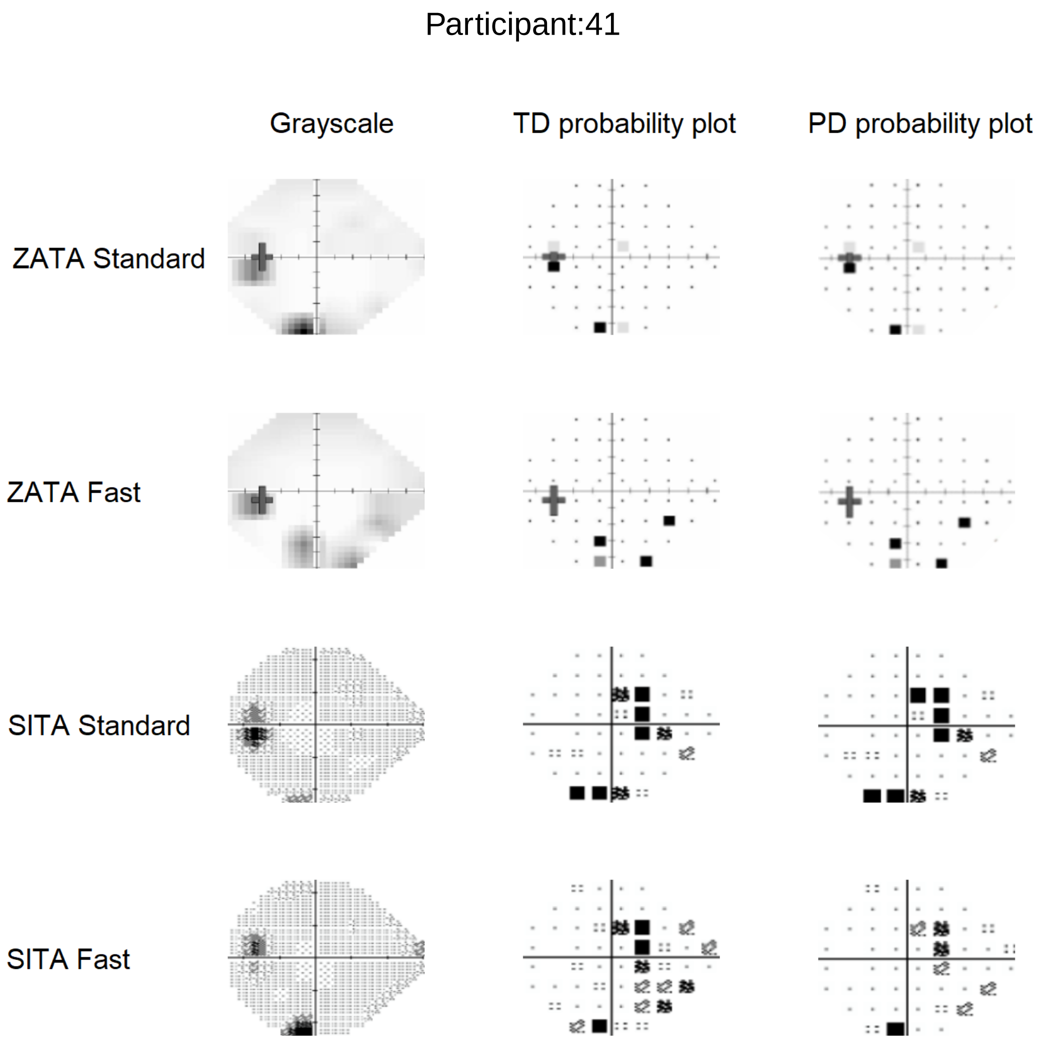
Participant:41
Grayscale
TD probability plot
PD probability plot
ZATA Standard
ZATA Fast
SITA Standard
SITA Fast

Participant:42

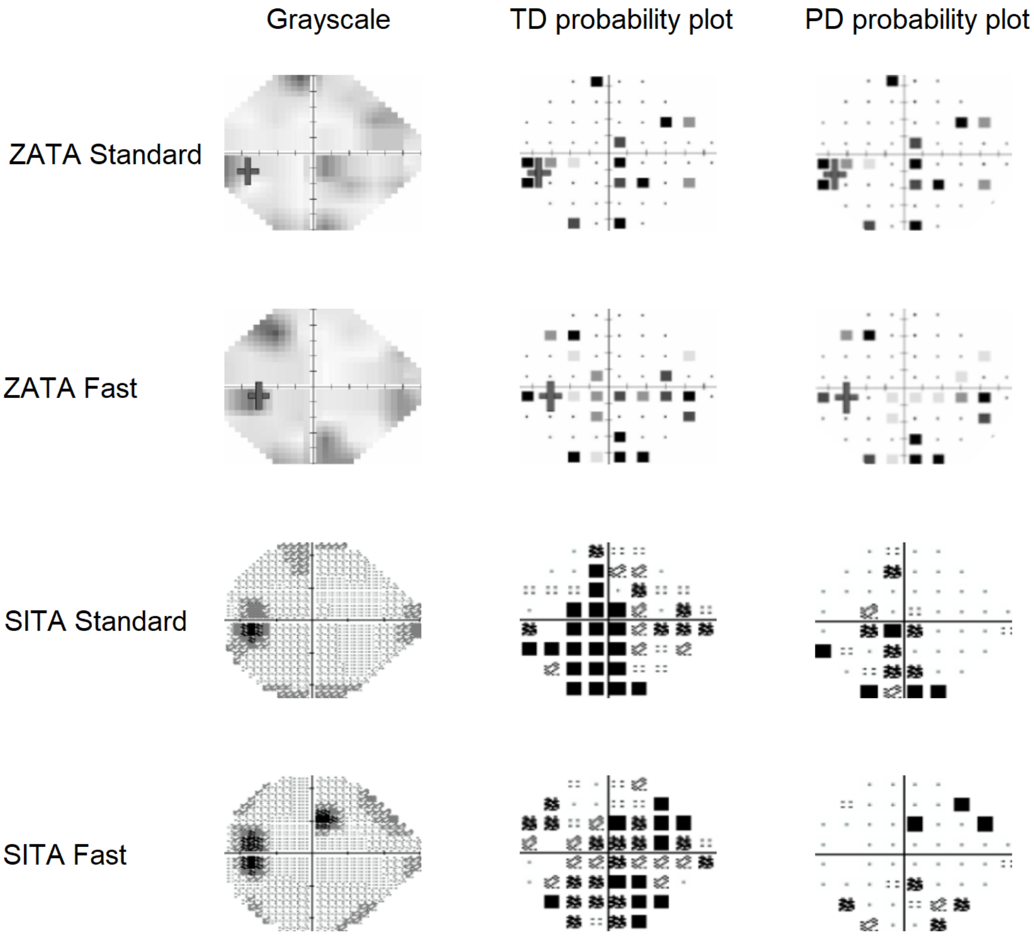

#### Participant:43

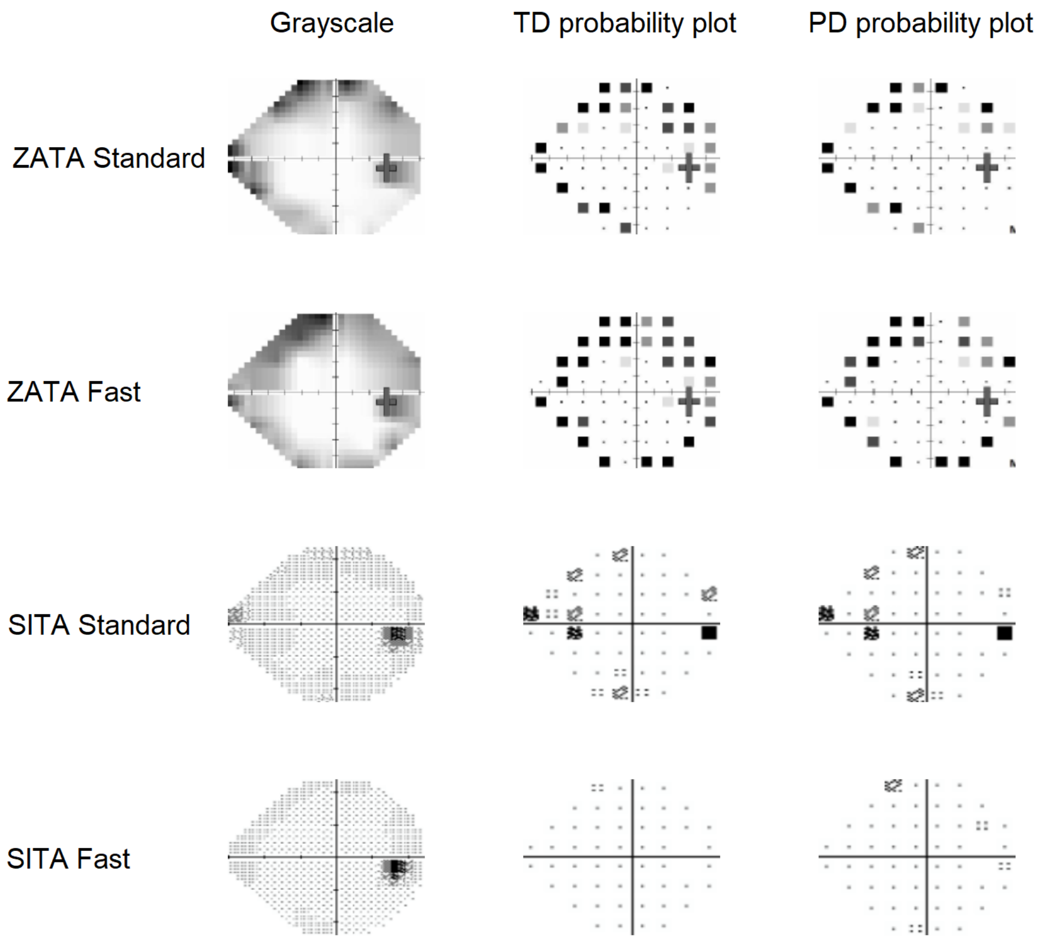

Participant:44

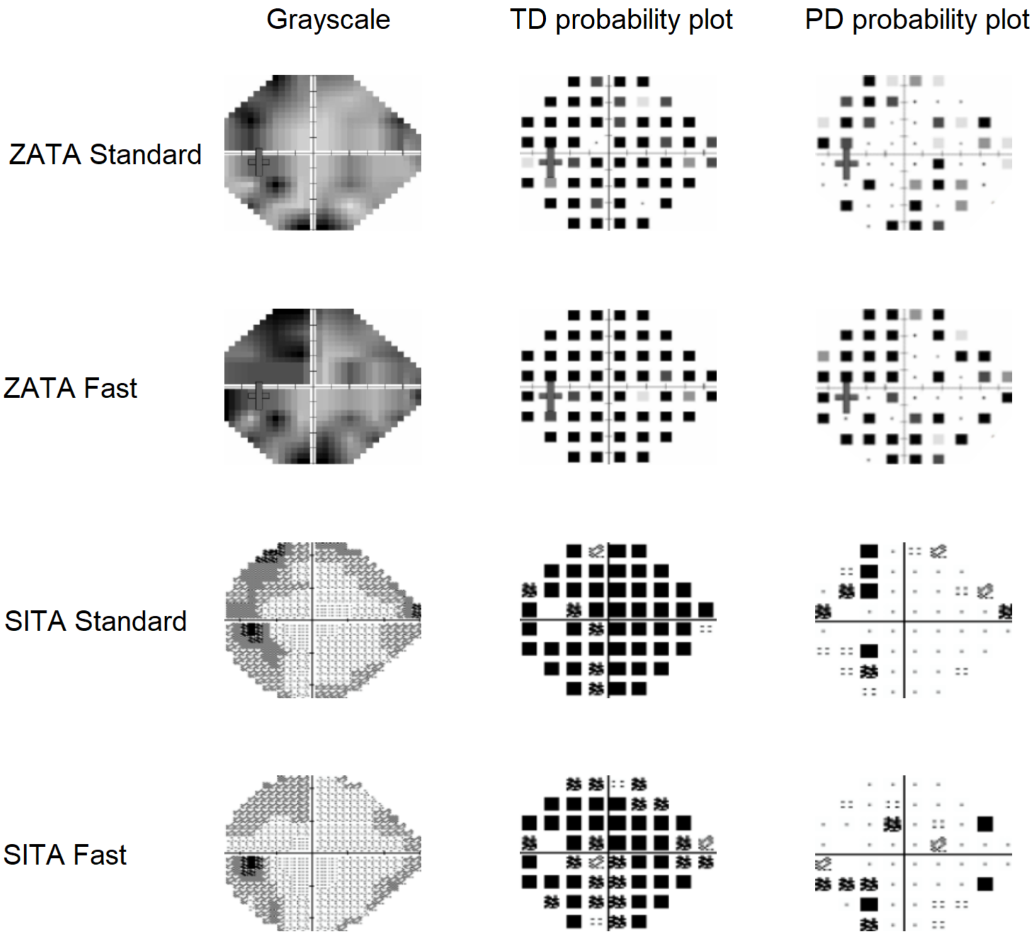

Participant:45

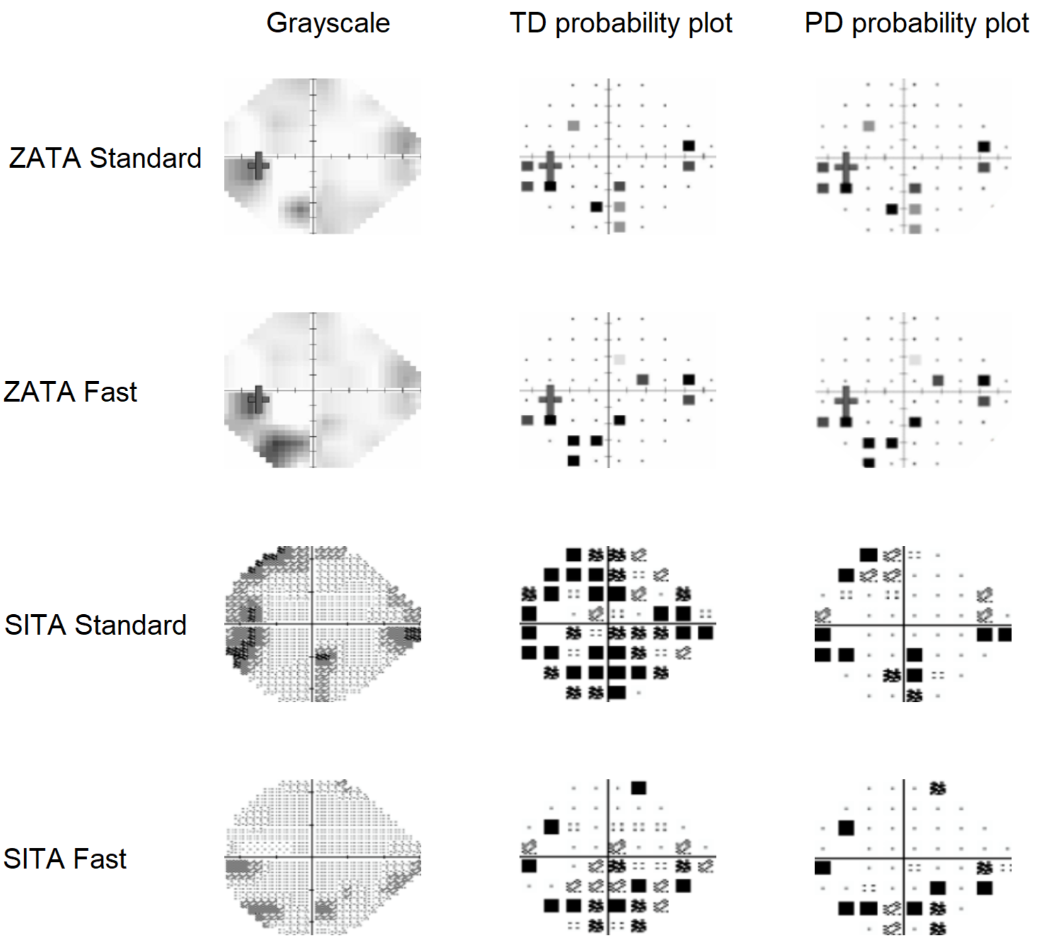

Participant:46

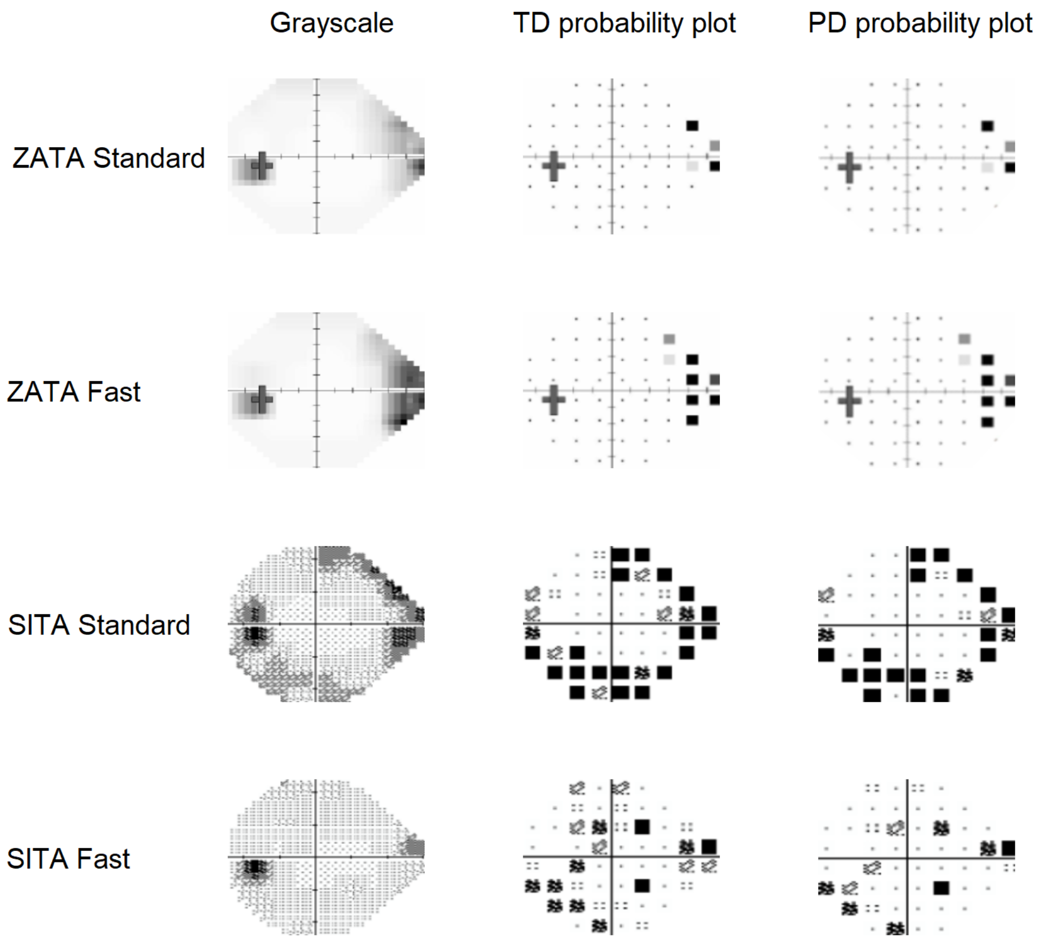

Participant:47

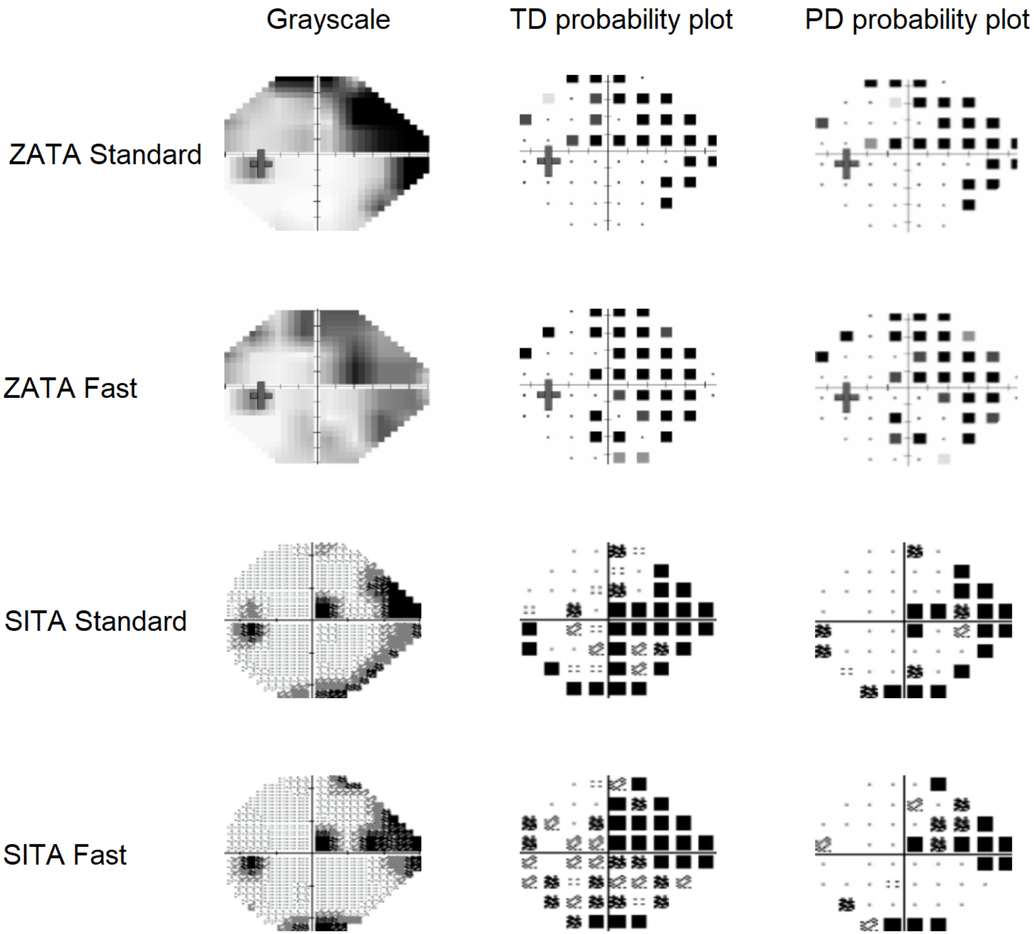

Participant:48

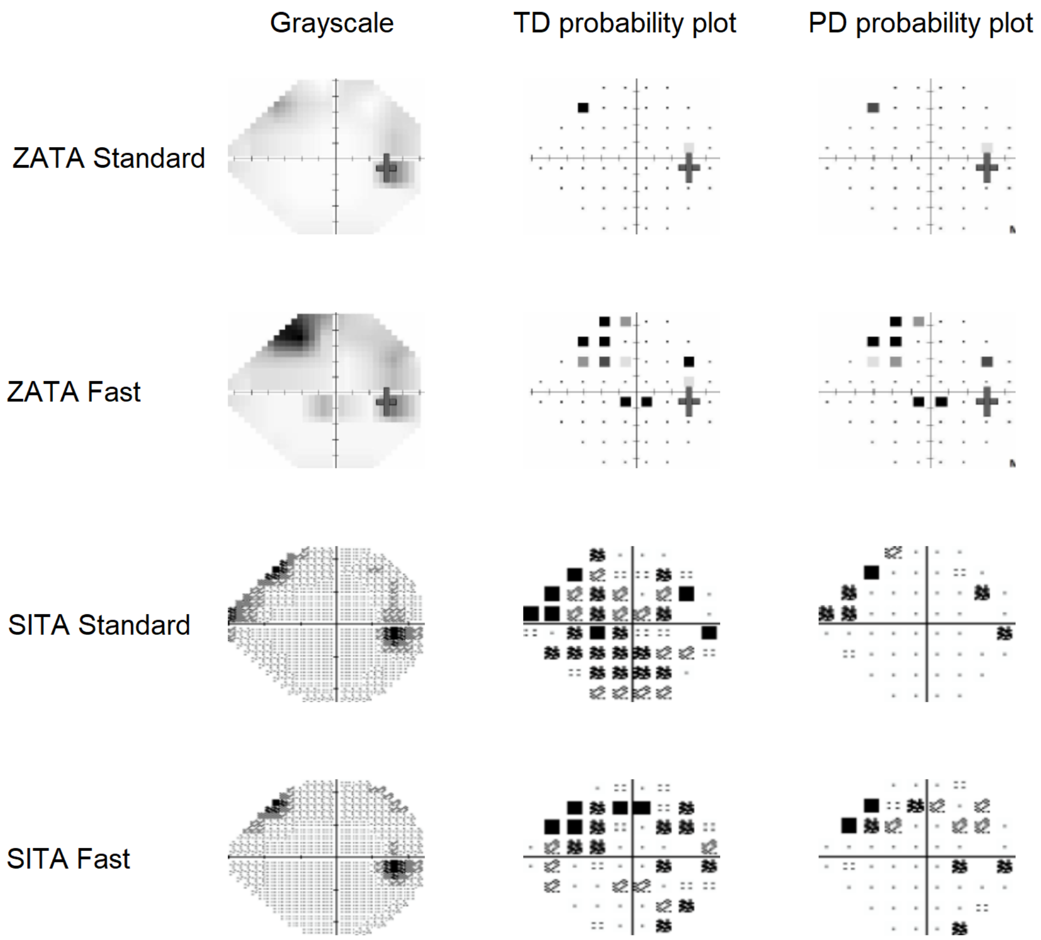

Participant:49

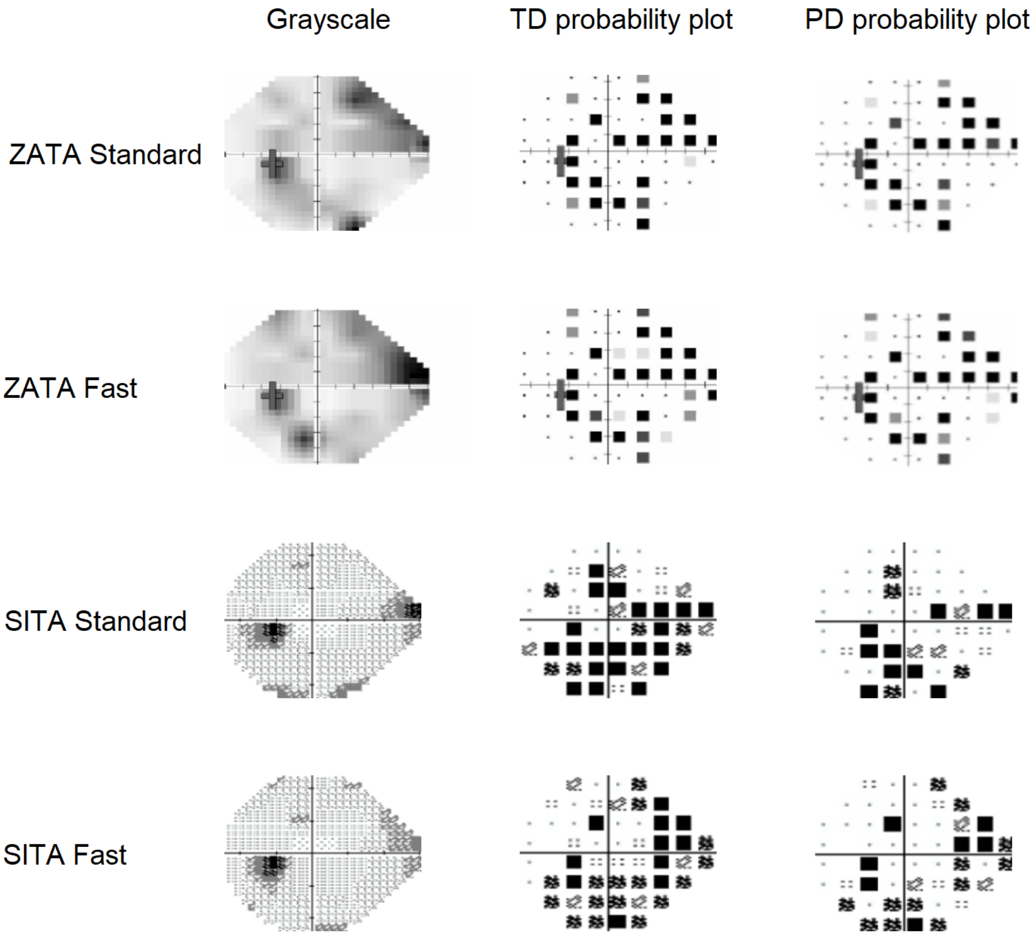

Participant:50

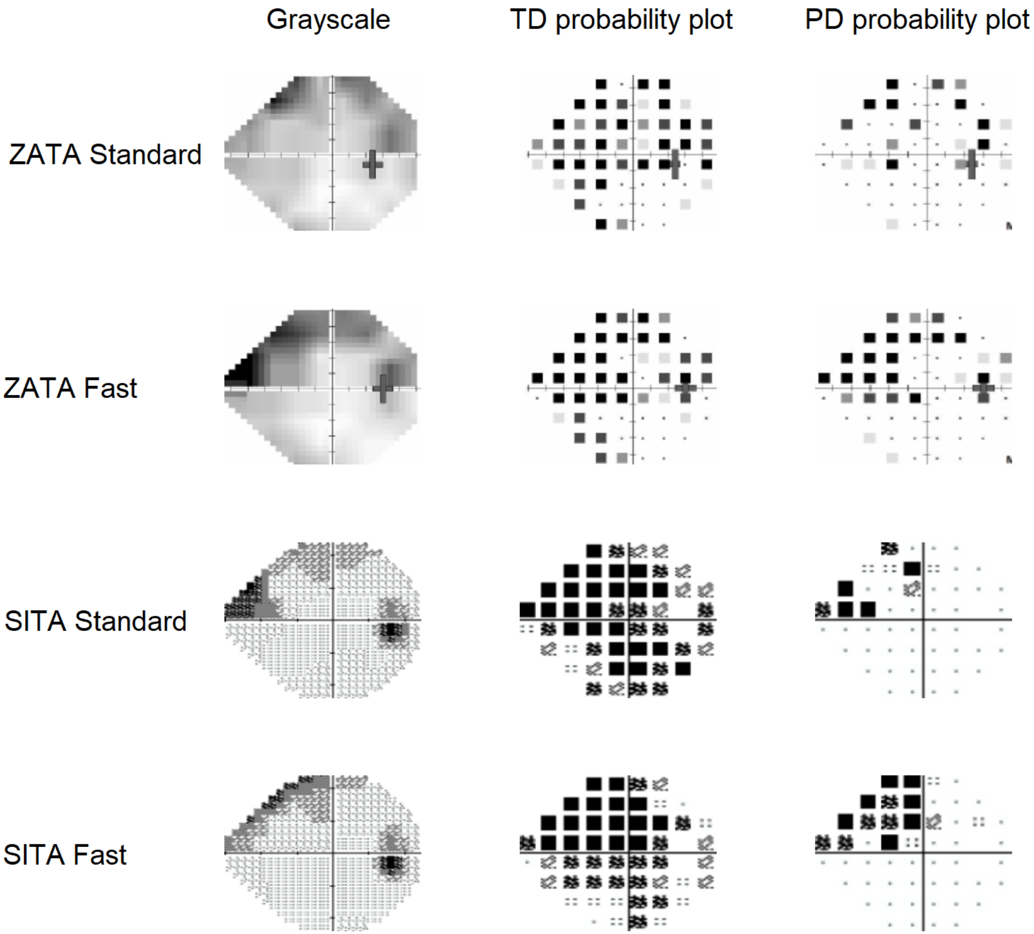

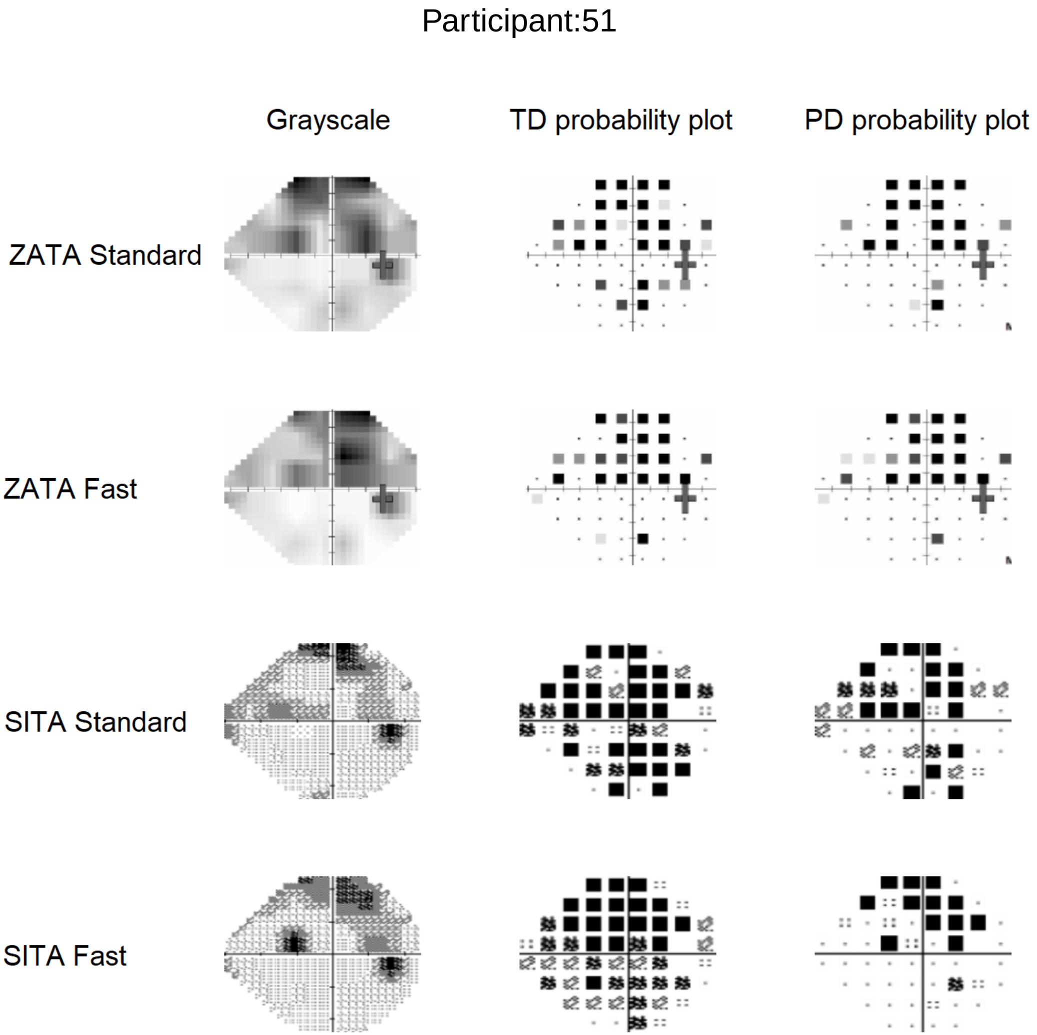

Participant:51
Grayscale
TD probability plot
PD probability plot
ZATA Standard
ZATA Fast
SITA Standard
SITA Fast

Participant:52

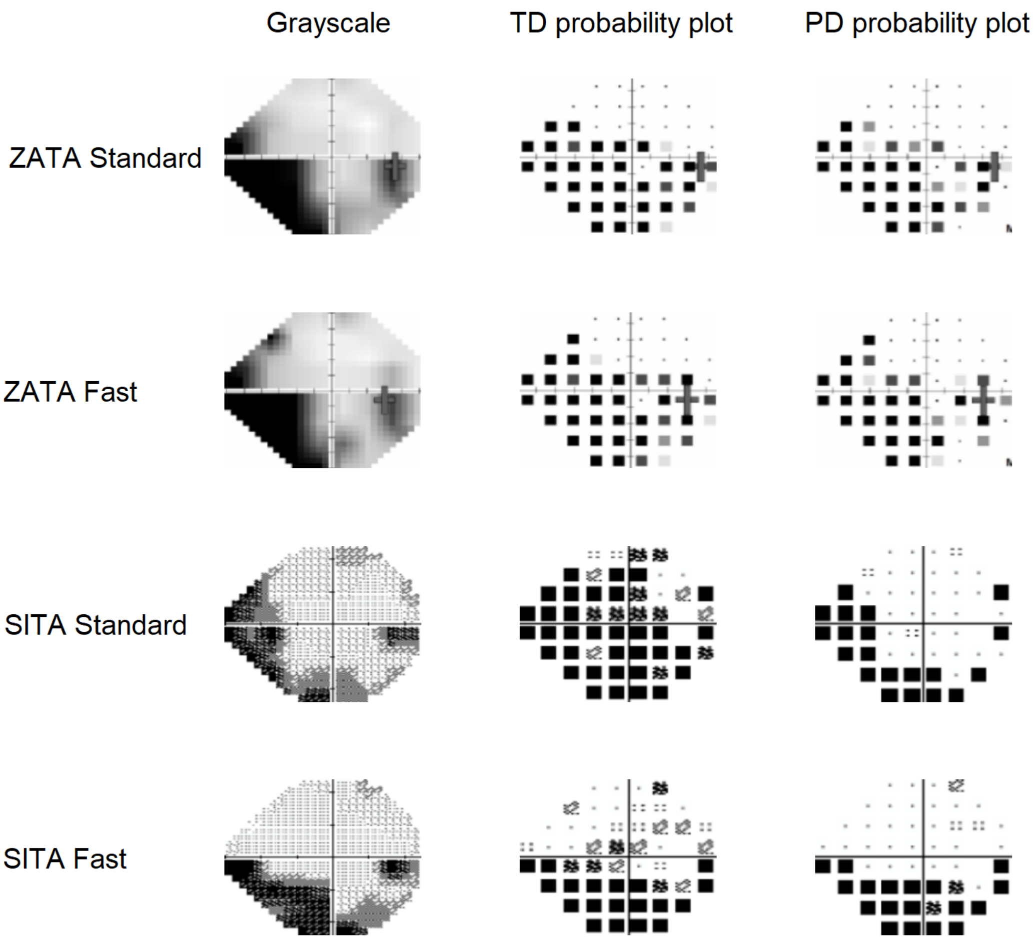

#### Participant:53

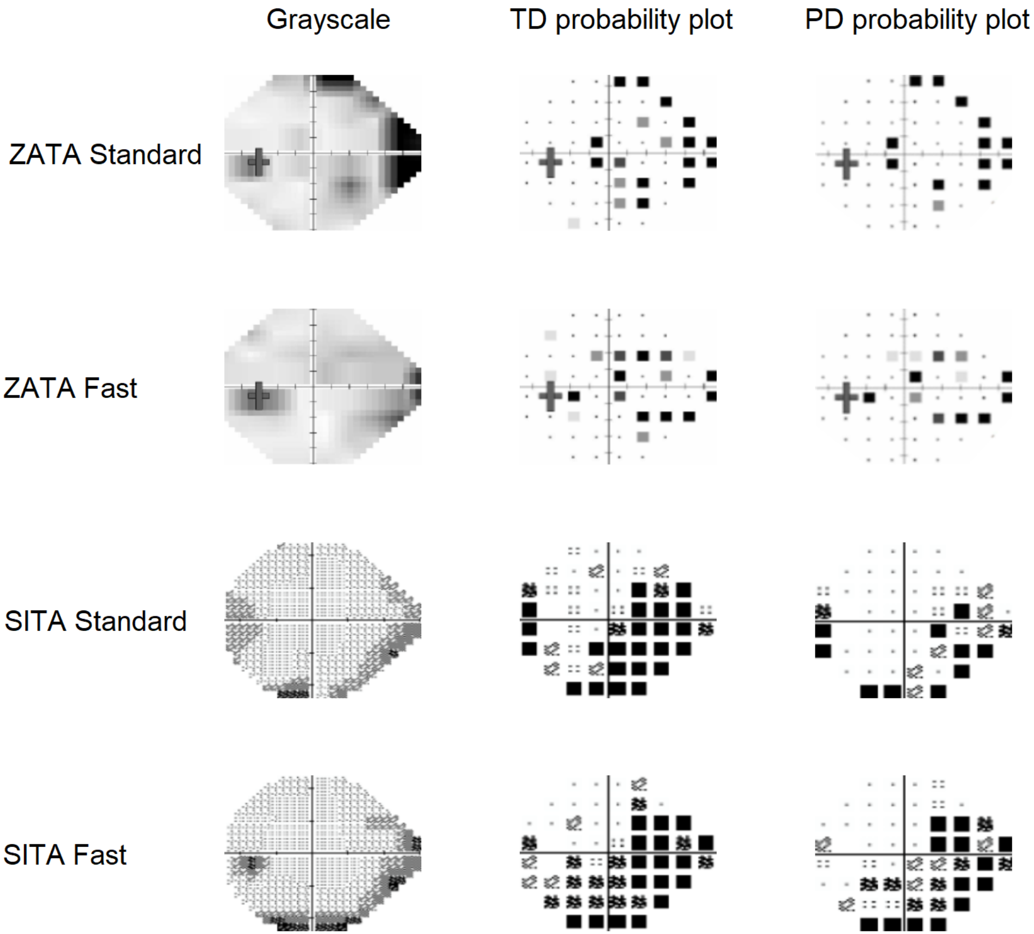

### Participant:54

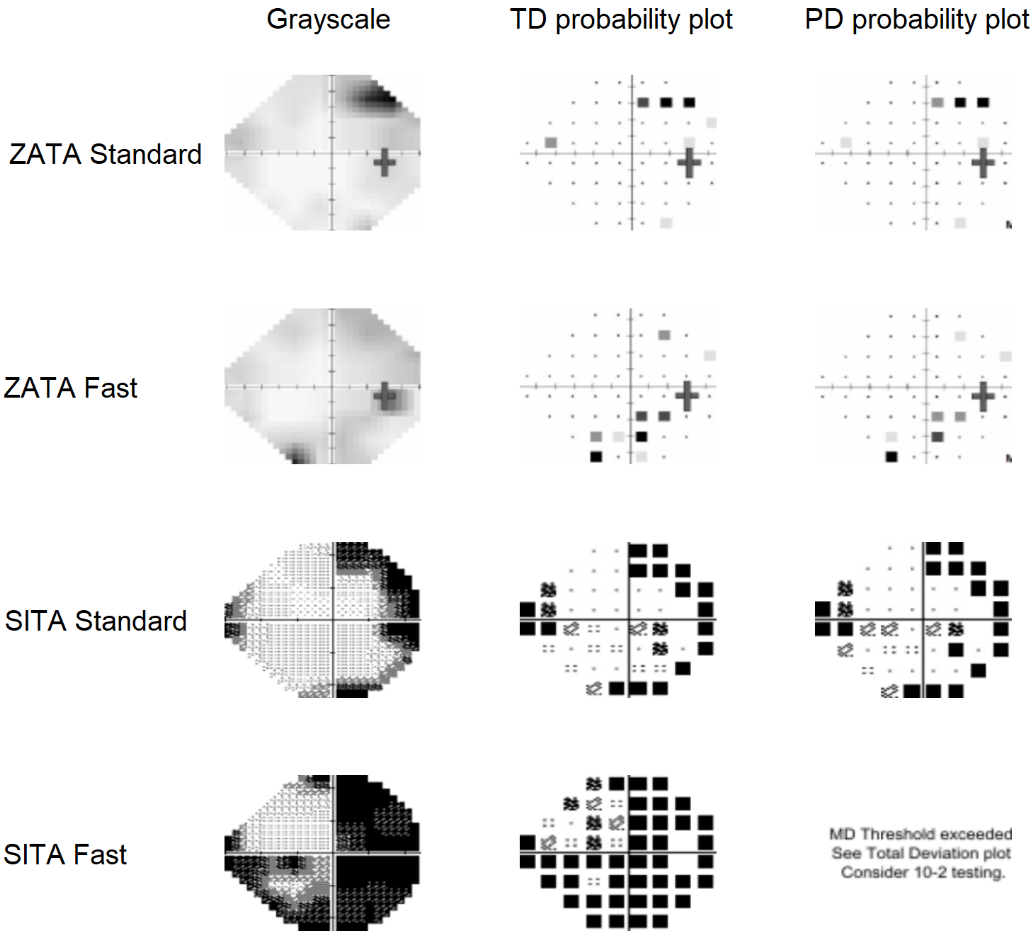

Participant:55

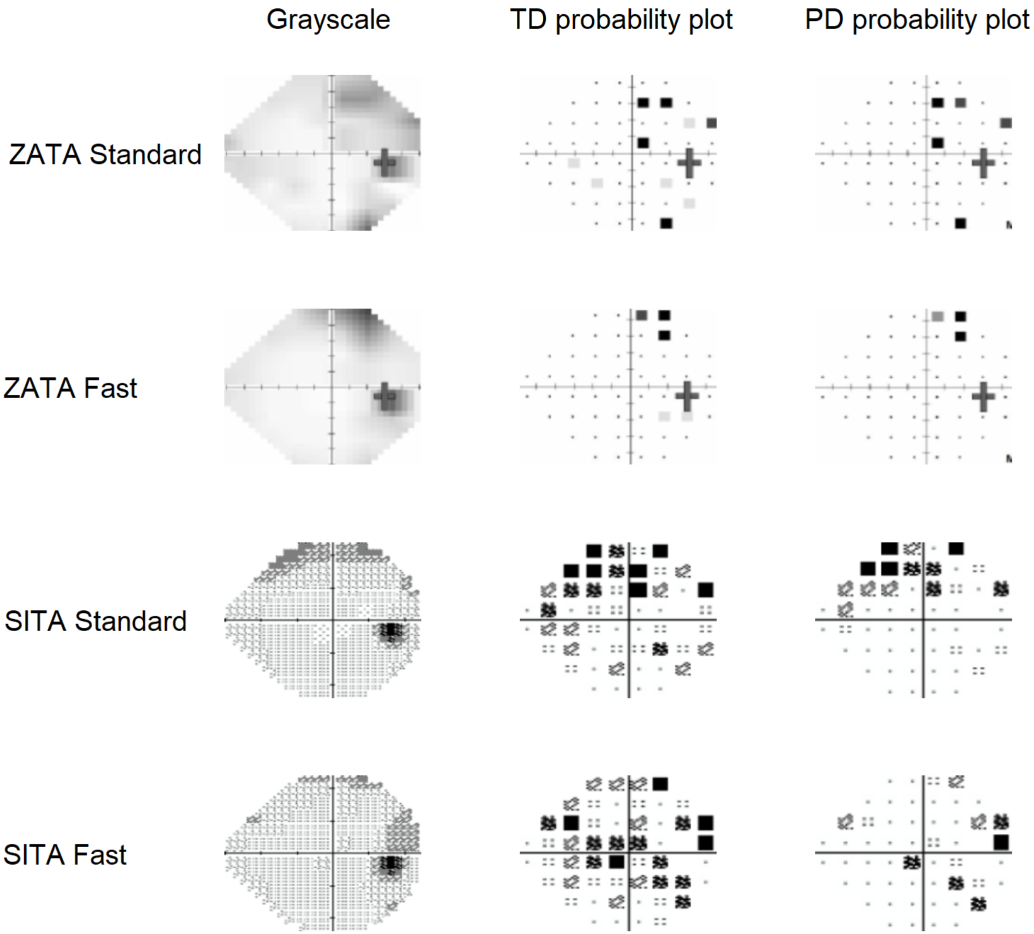

### Participant:56

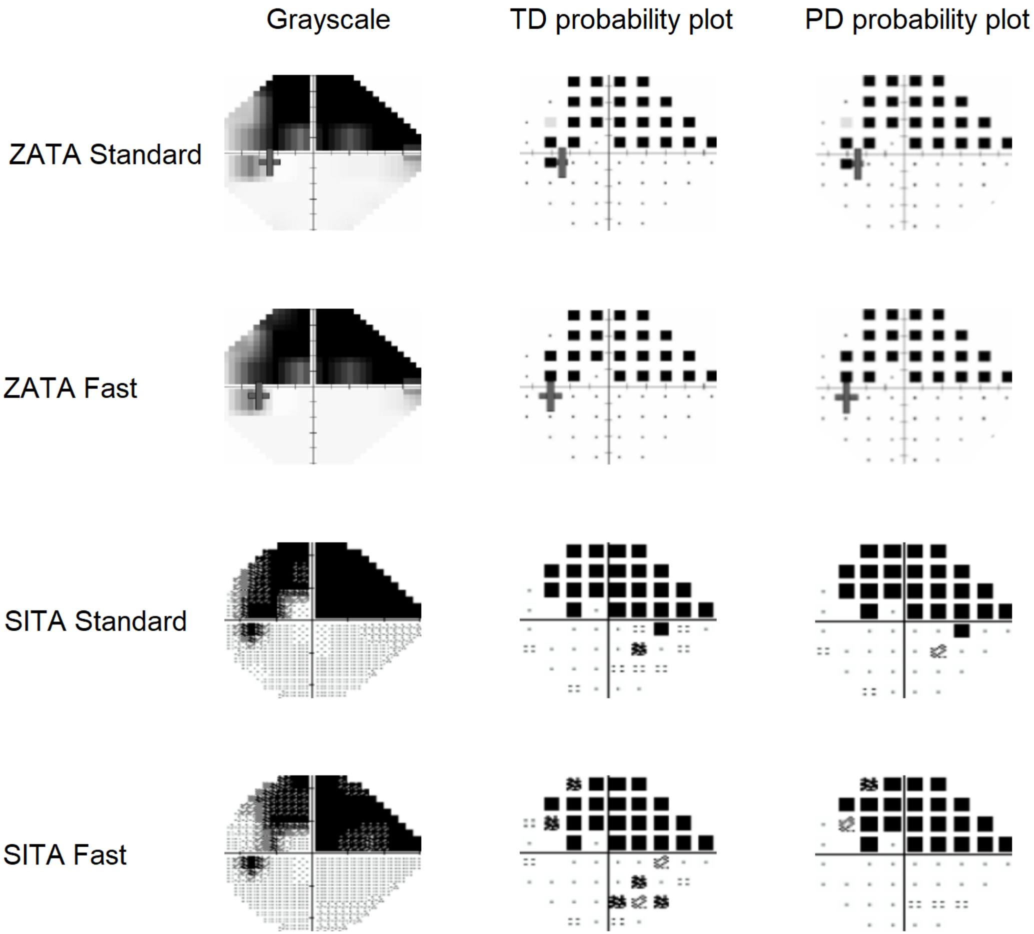

#### Participant:57

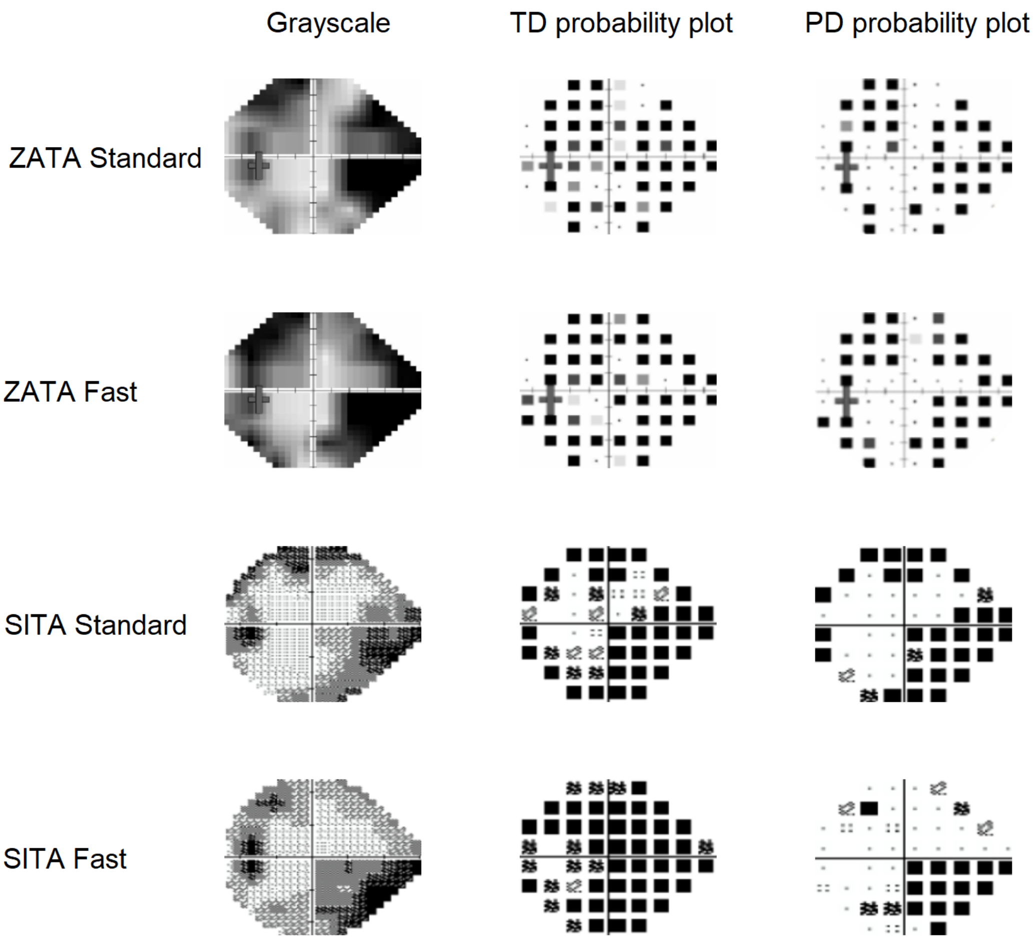

Participant:58

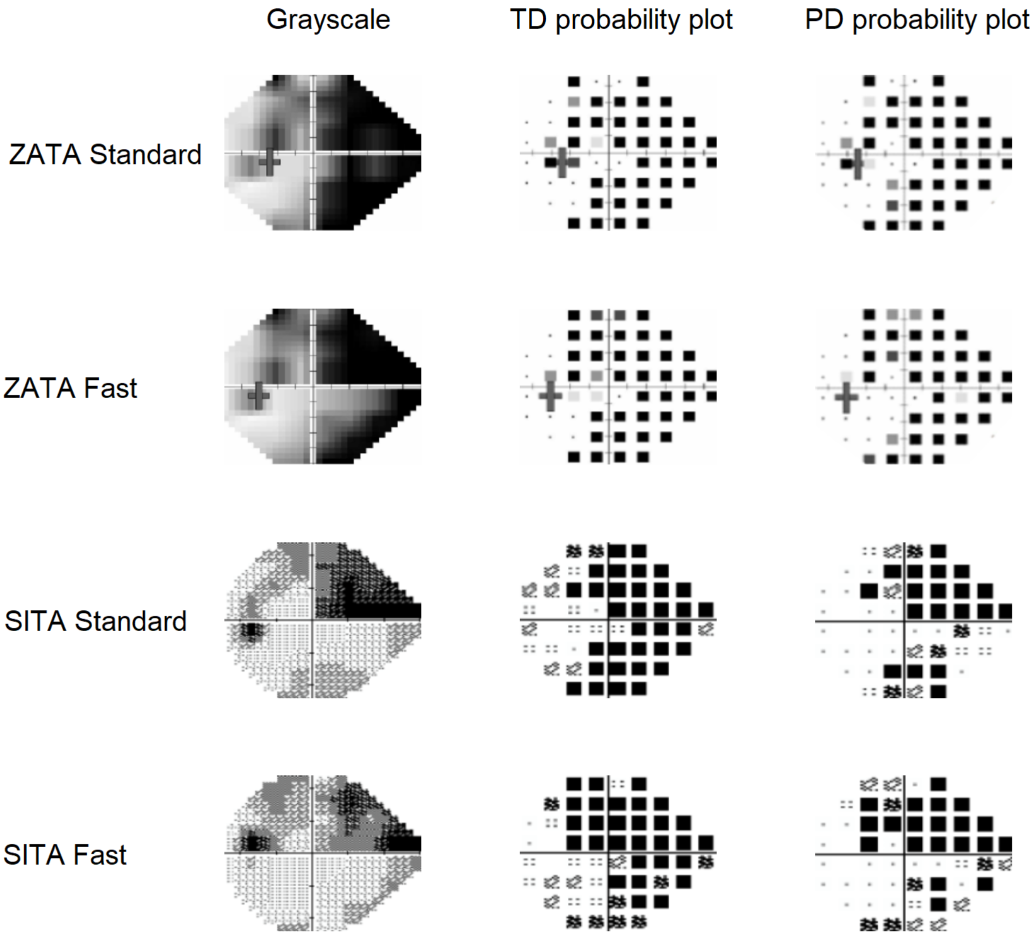

#### Participant:59

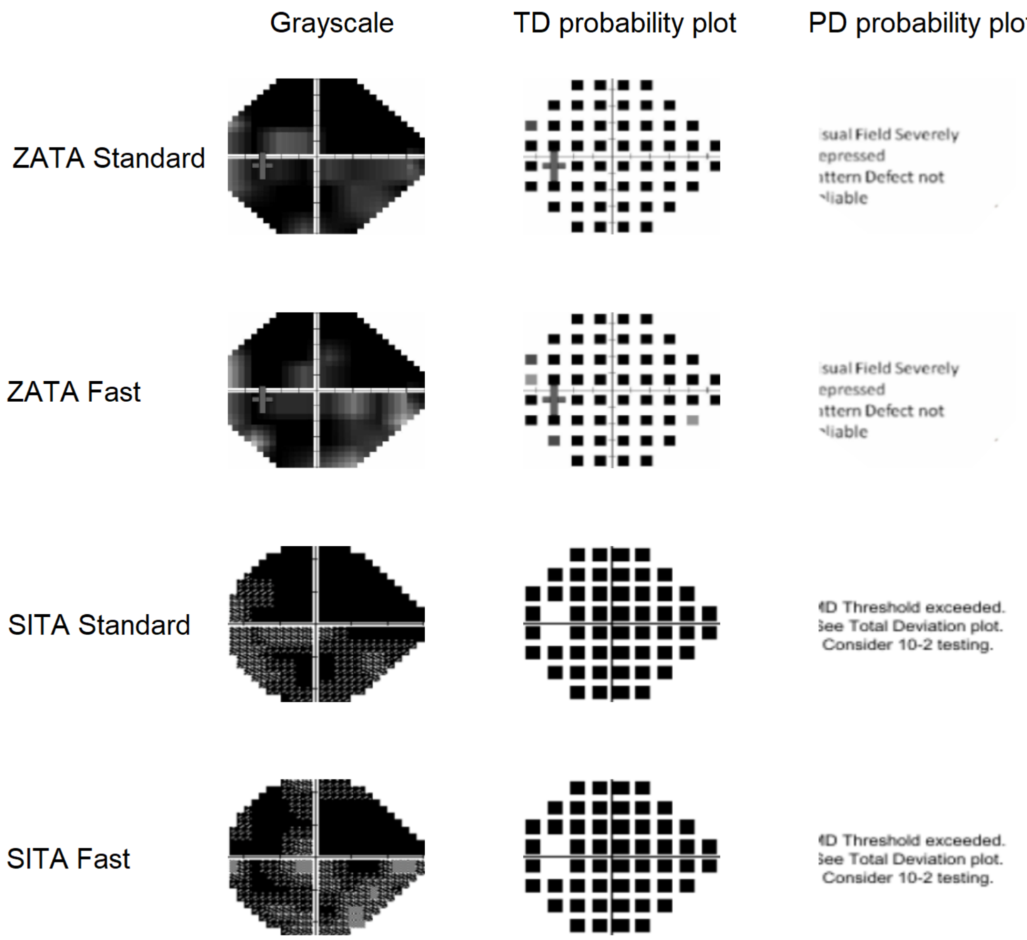

Participant:60

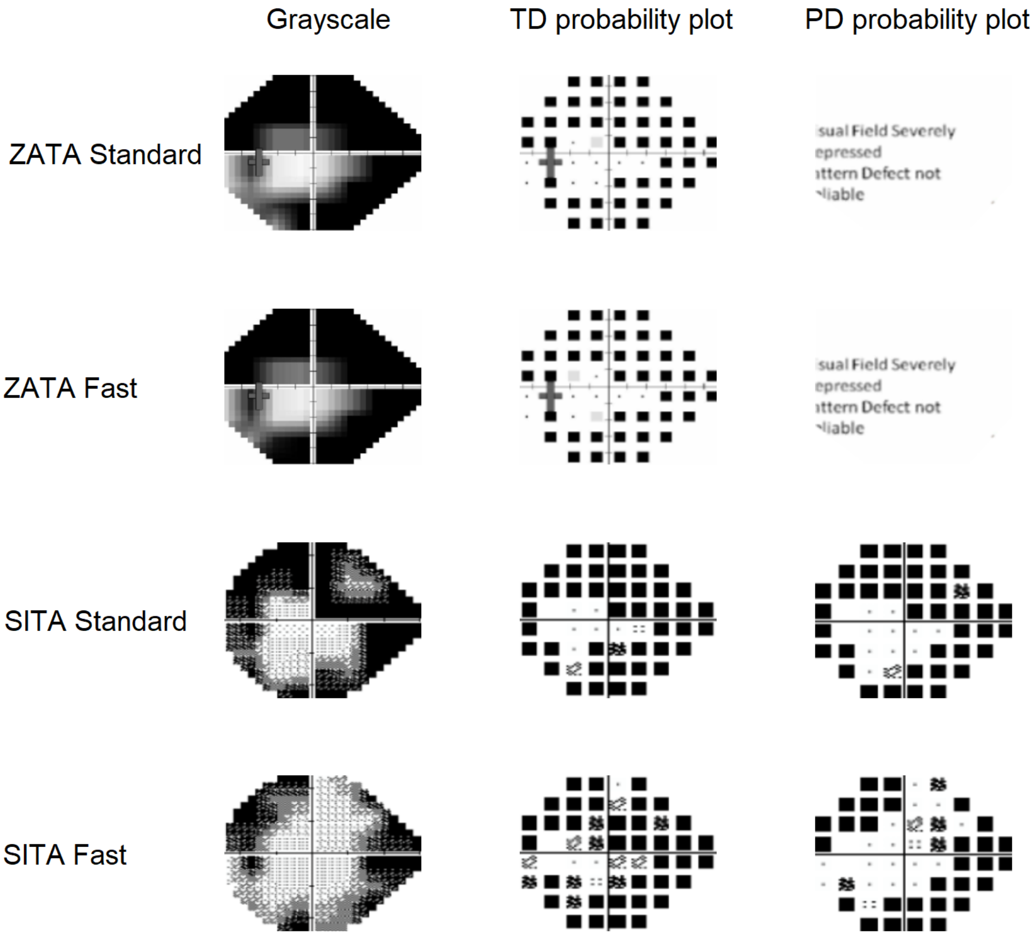

Participant:61

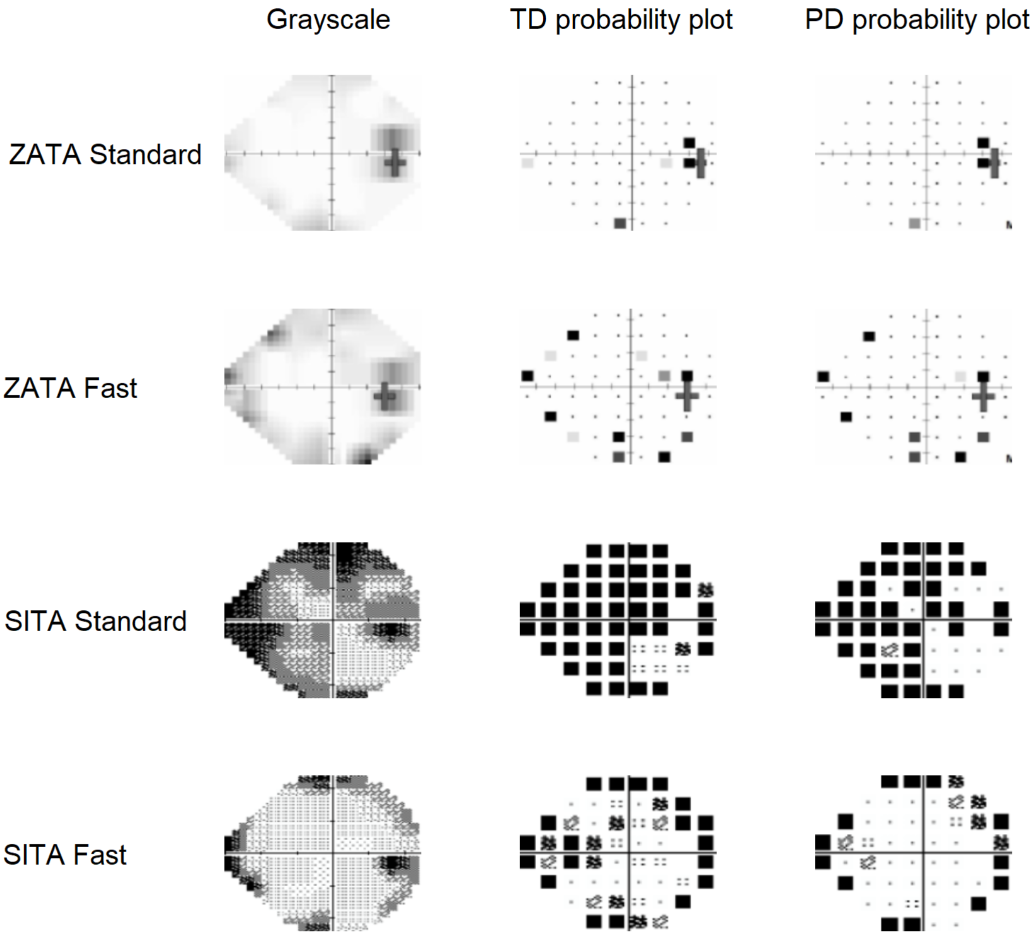

Participant:62

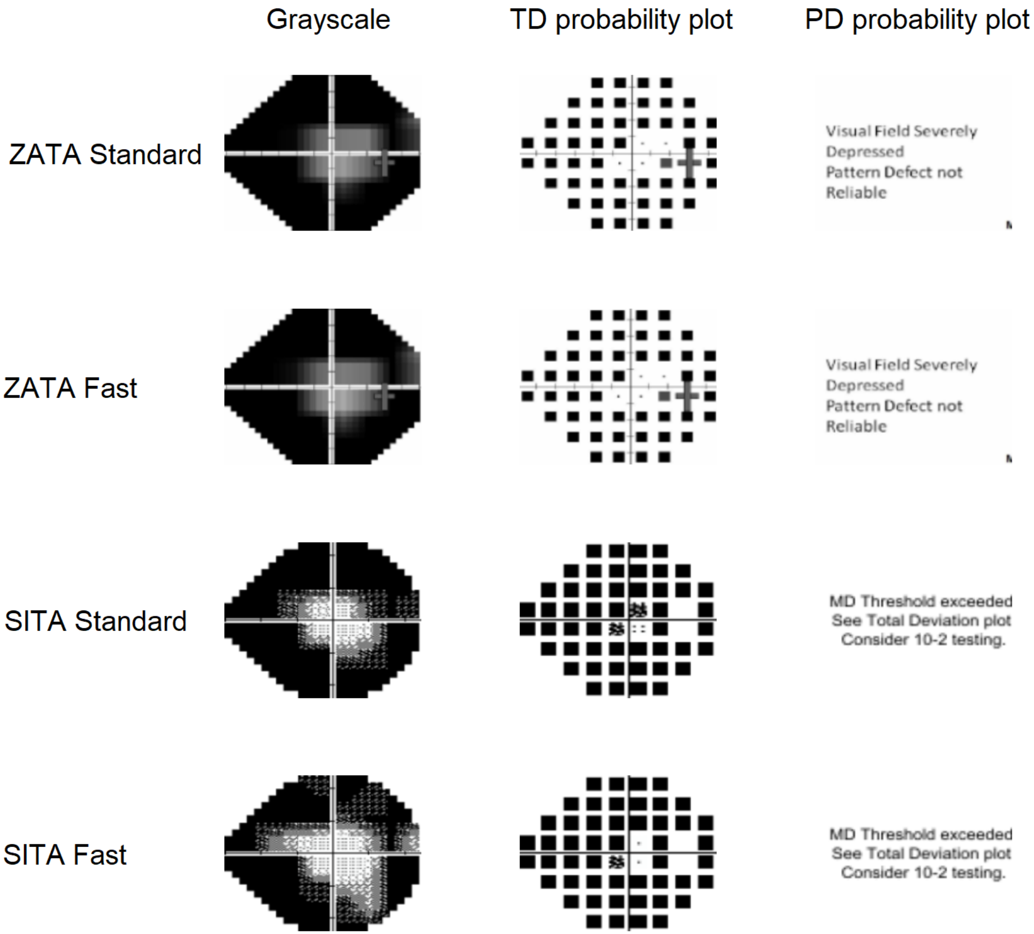

### Participant:63

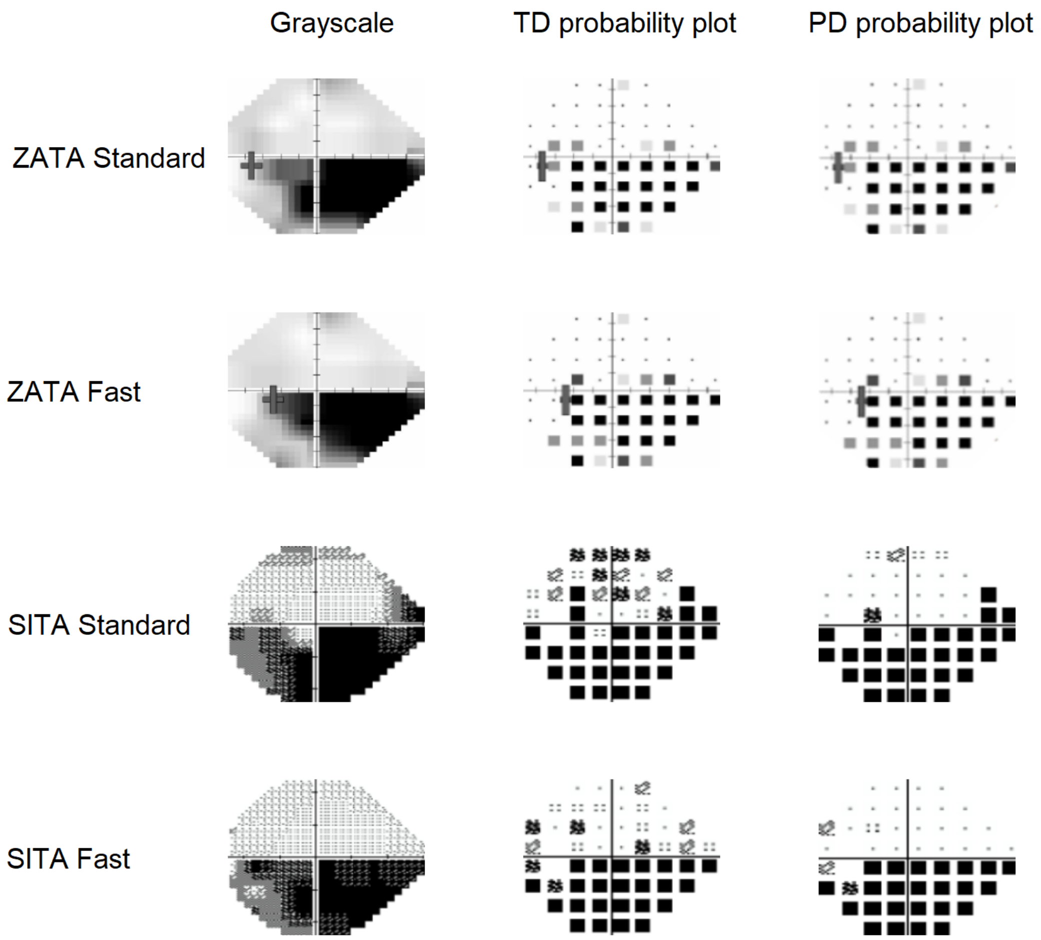

Participant:64

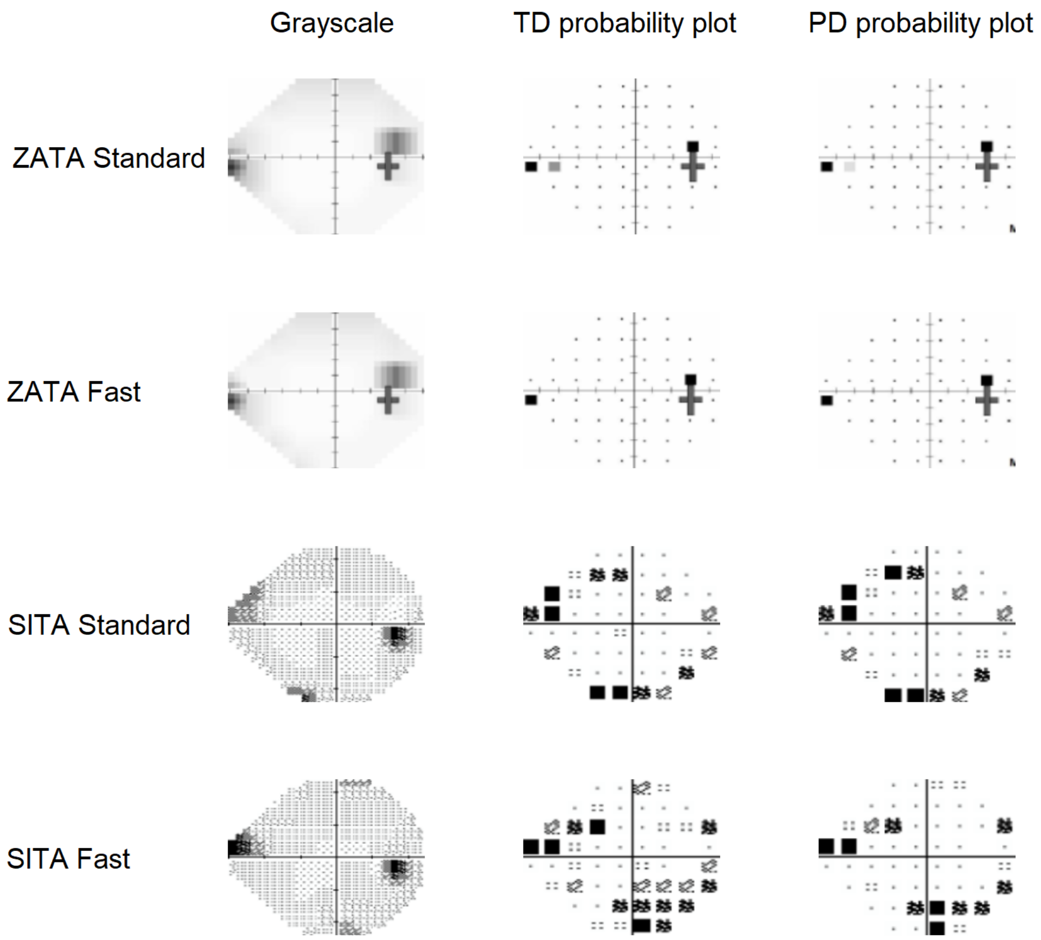

Participant:65

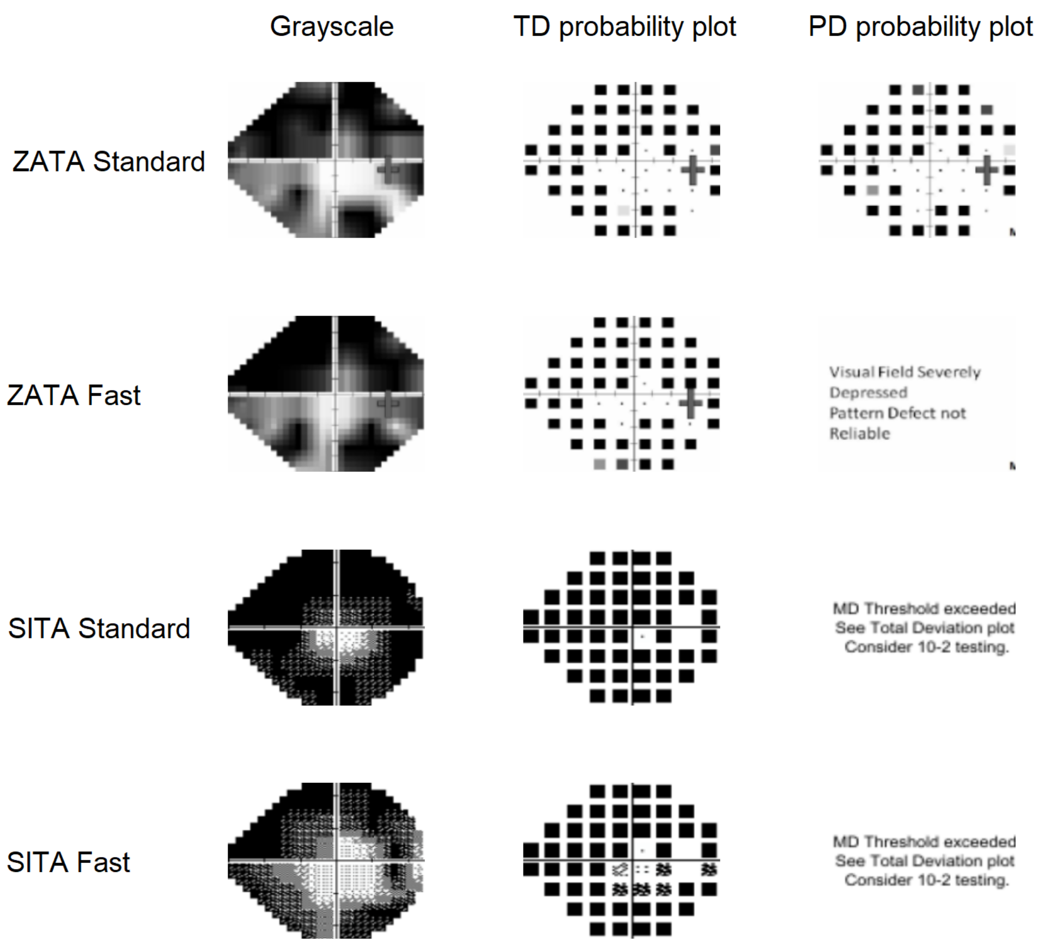

Participant:66

Grayscale | TD probability plot | PD probability plot

ZATA Standard

ZATA Fast

SITA Standard

SITA Fast

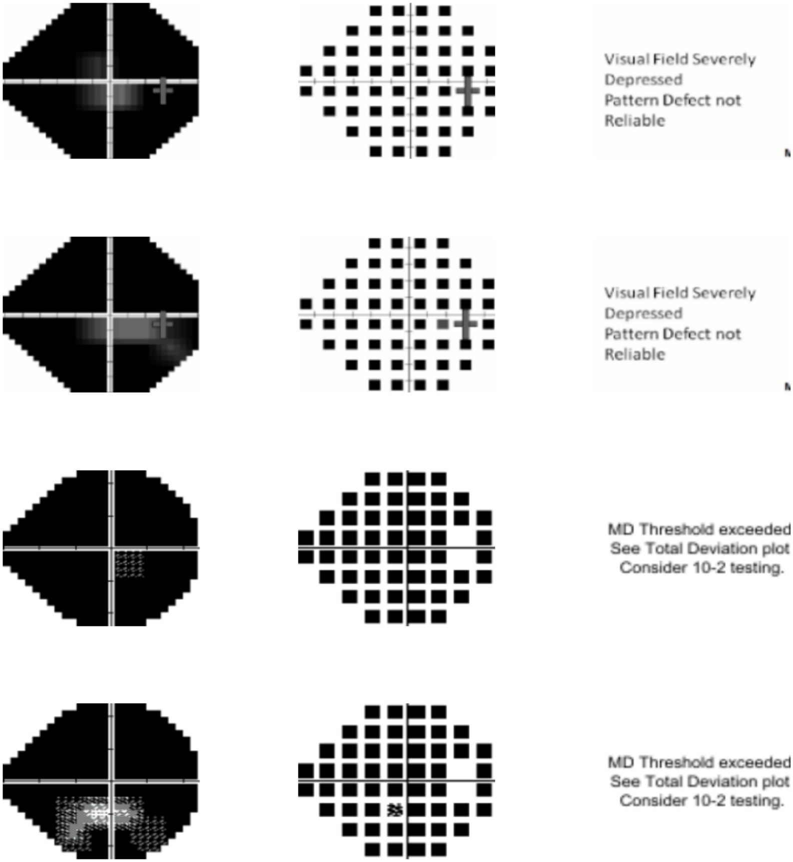

Participant:67

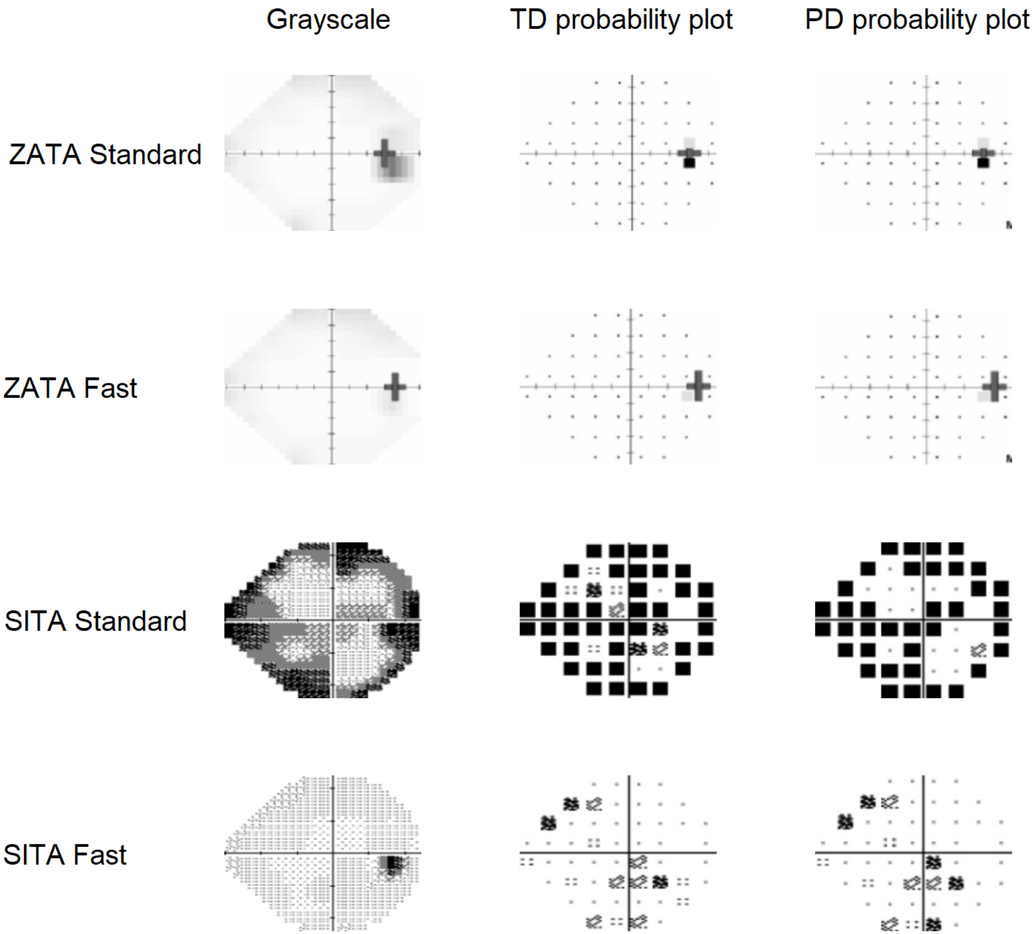

Participant:68

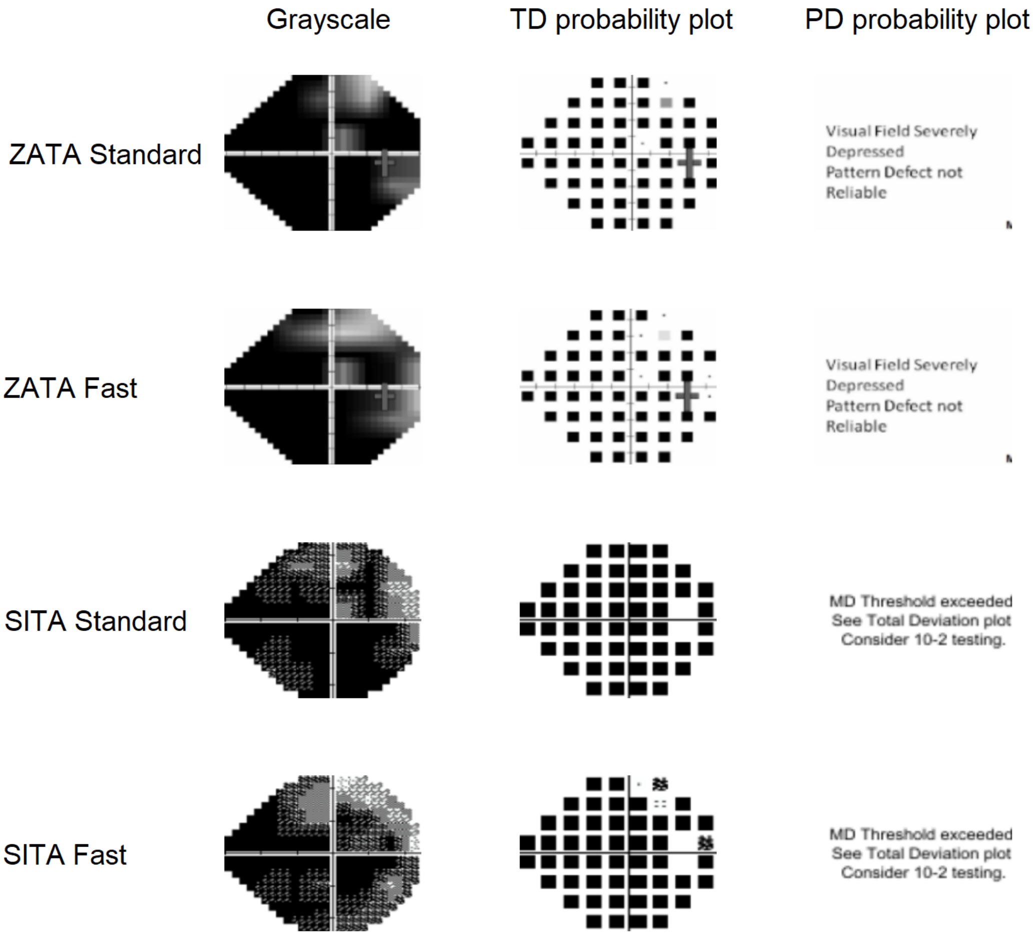

### Participant:69

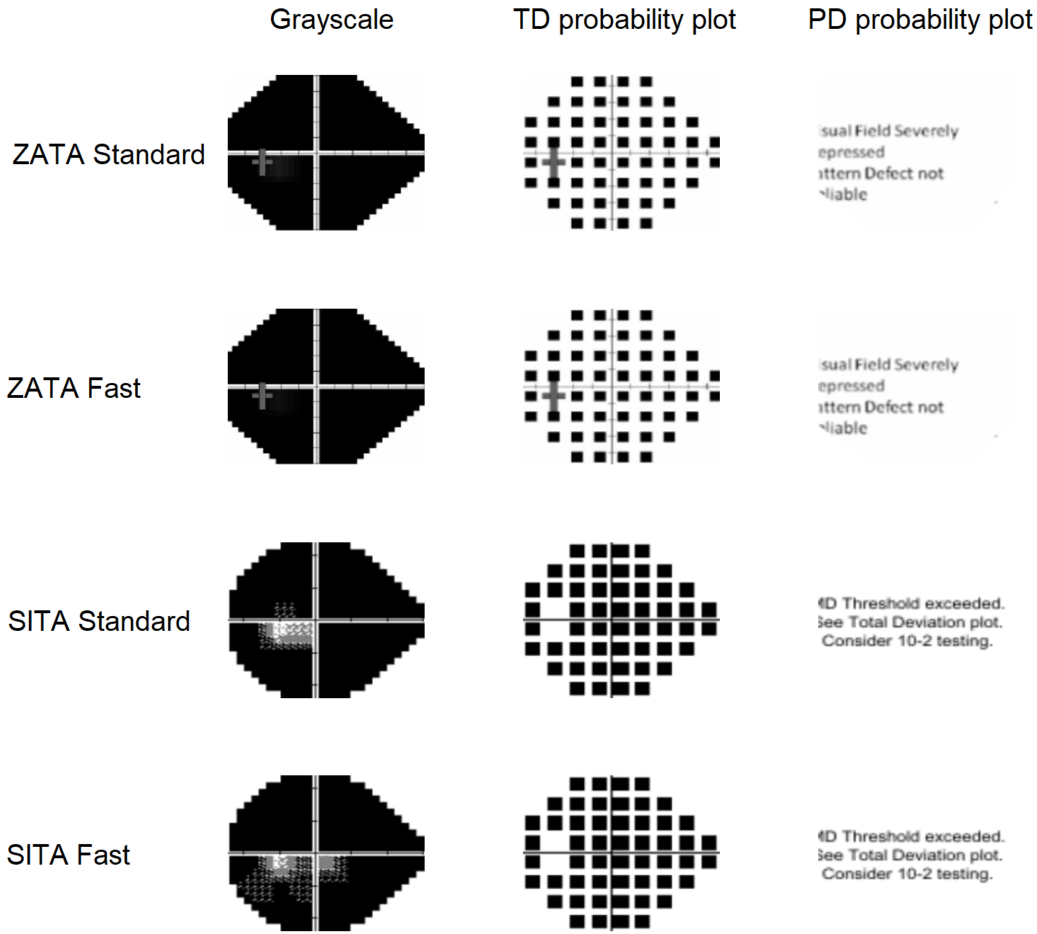

Participant:70

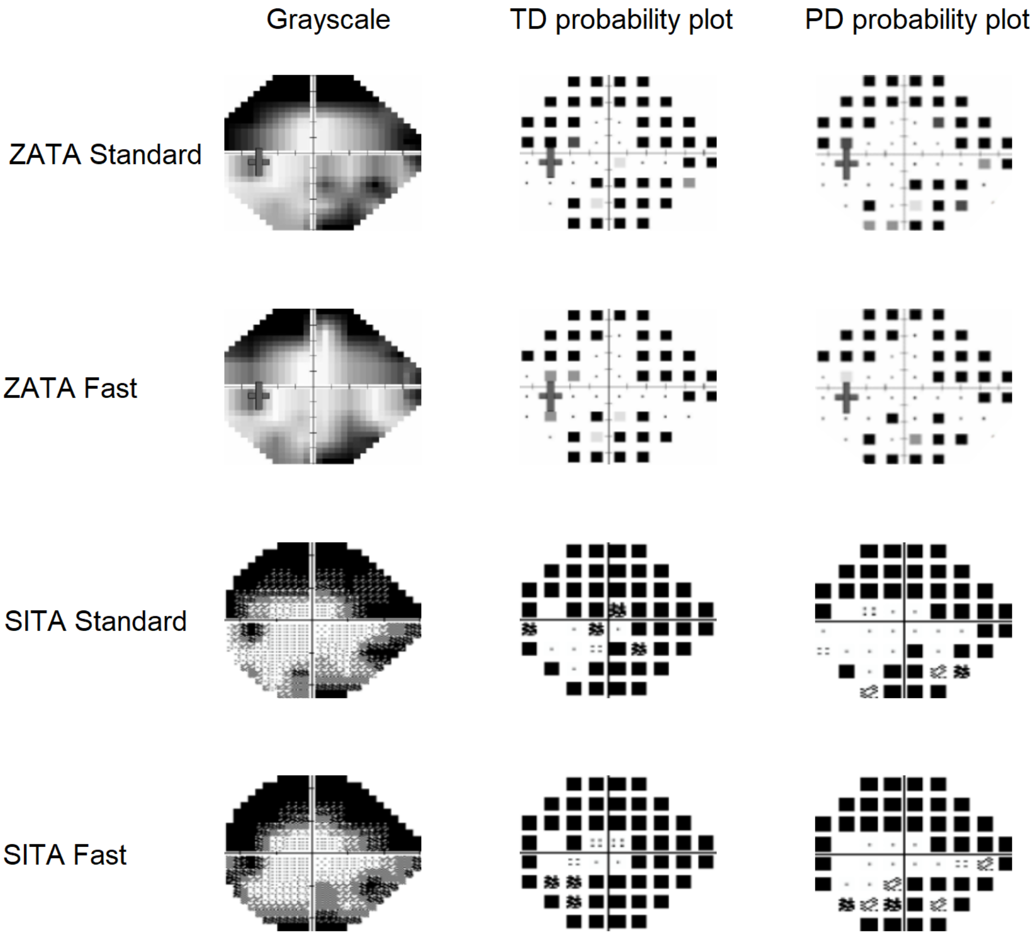

Participant:71

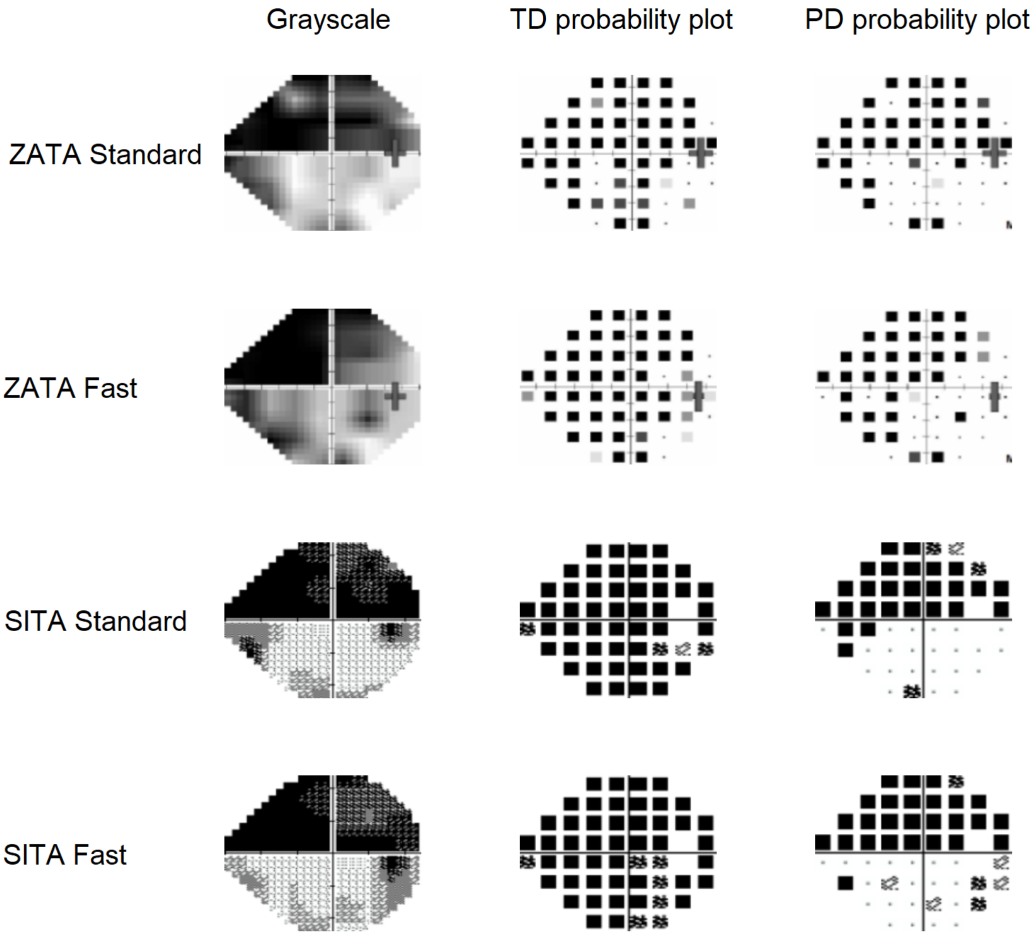

#### Participant:72

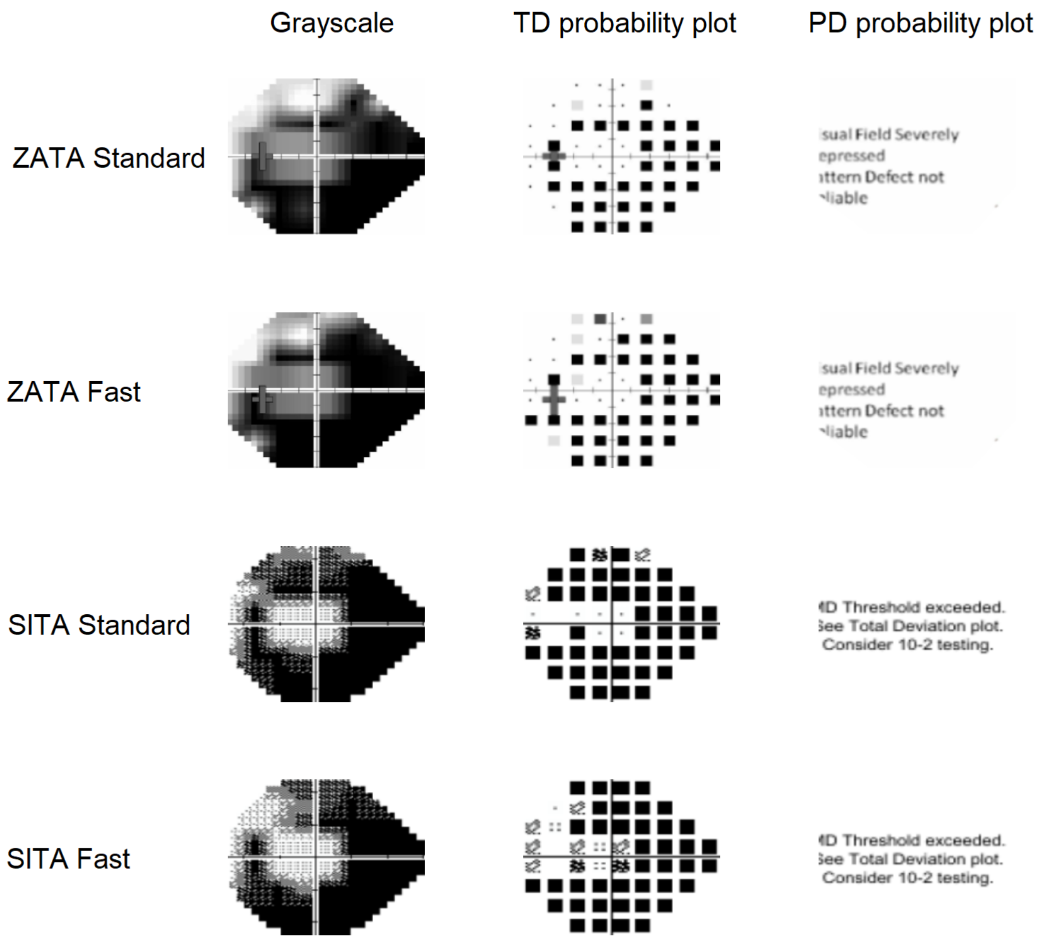

Participant:73

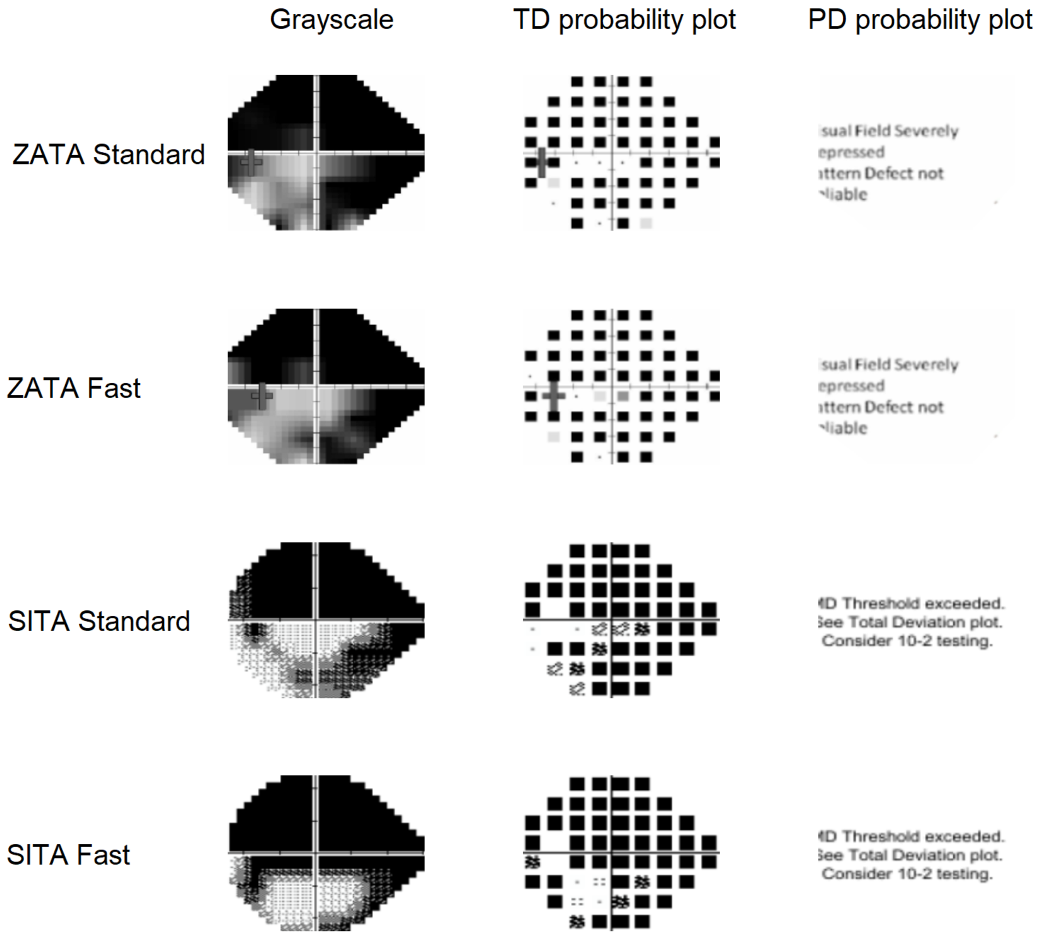

Participant:74

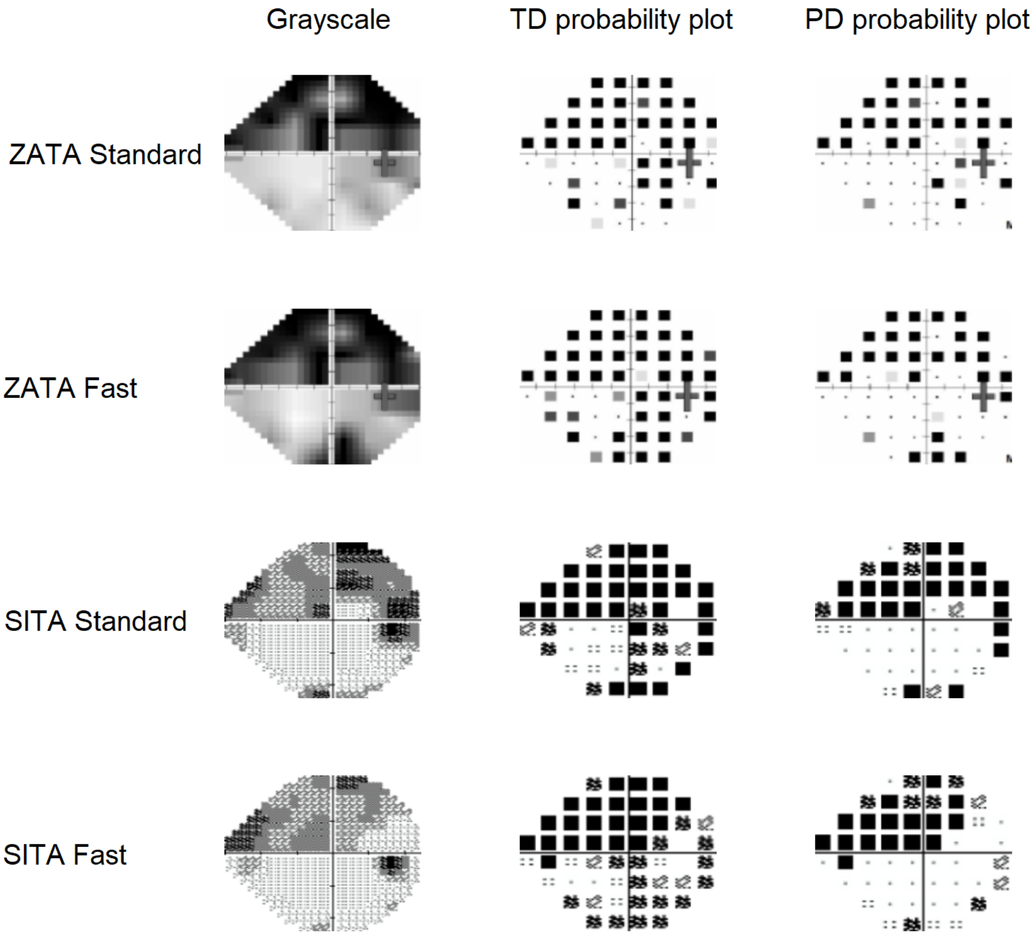

Participant:75

Grayscale | TD probability plot | PD probability plot

ZATA Standard

ZATA Fast

SITA Standard

SITA Fast

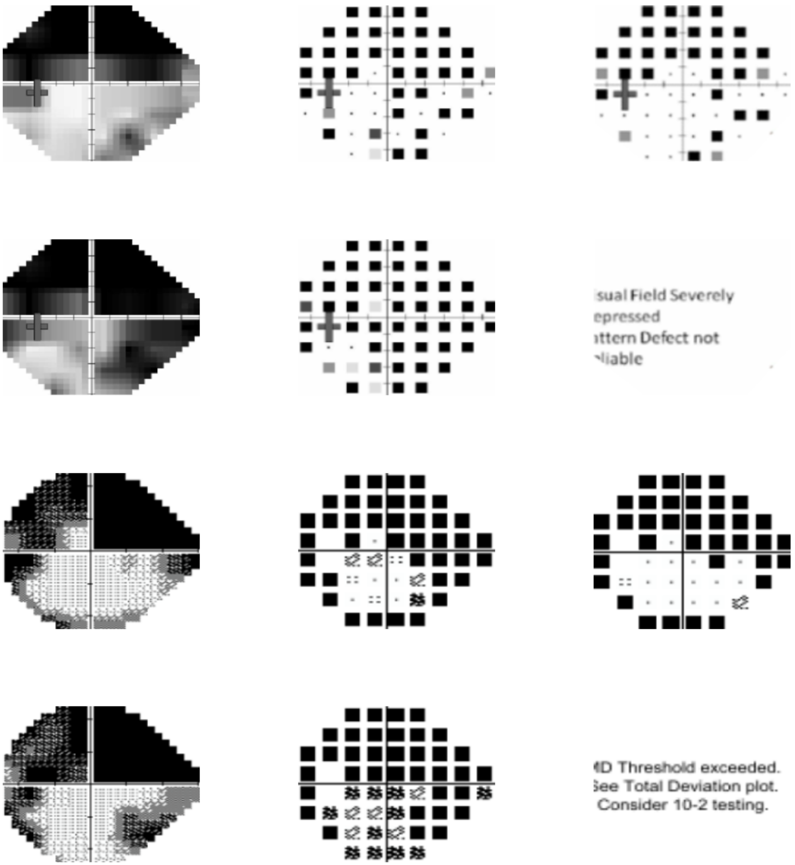

Participant:76

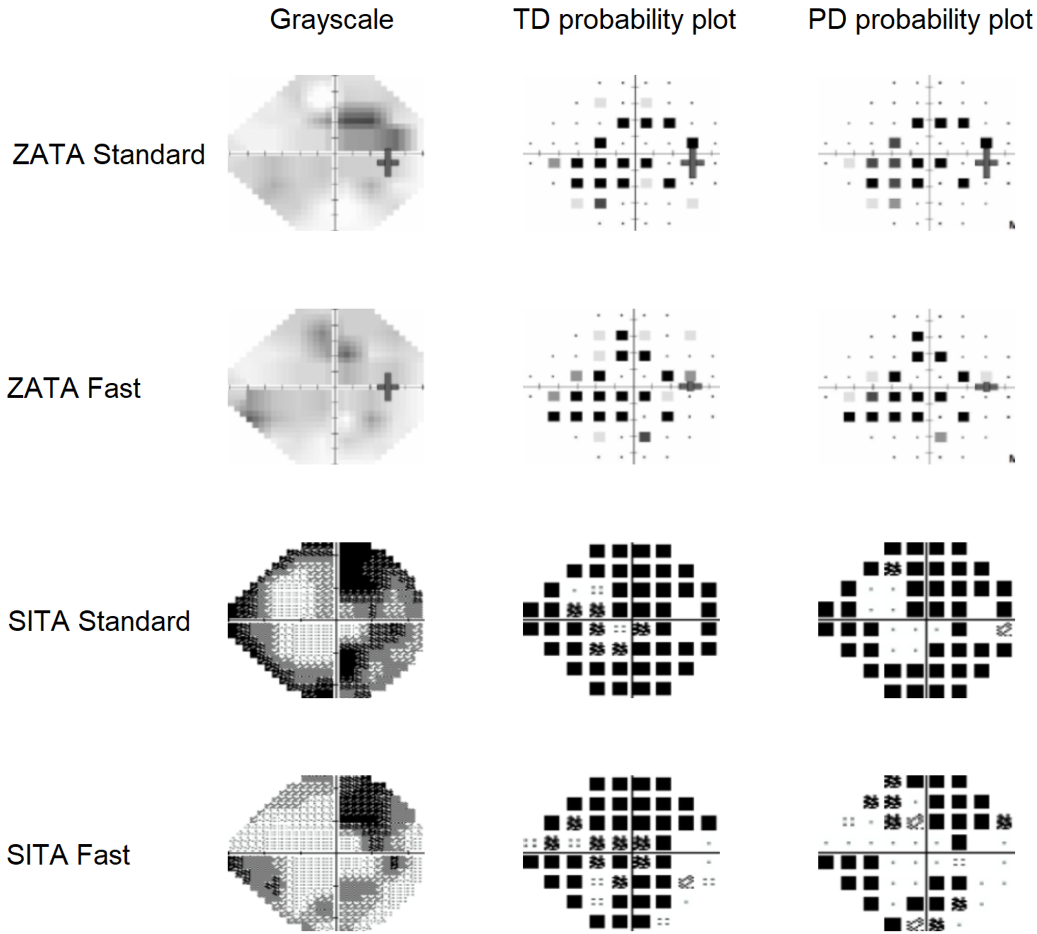

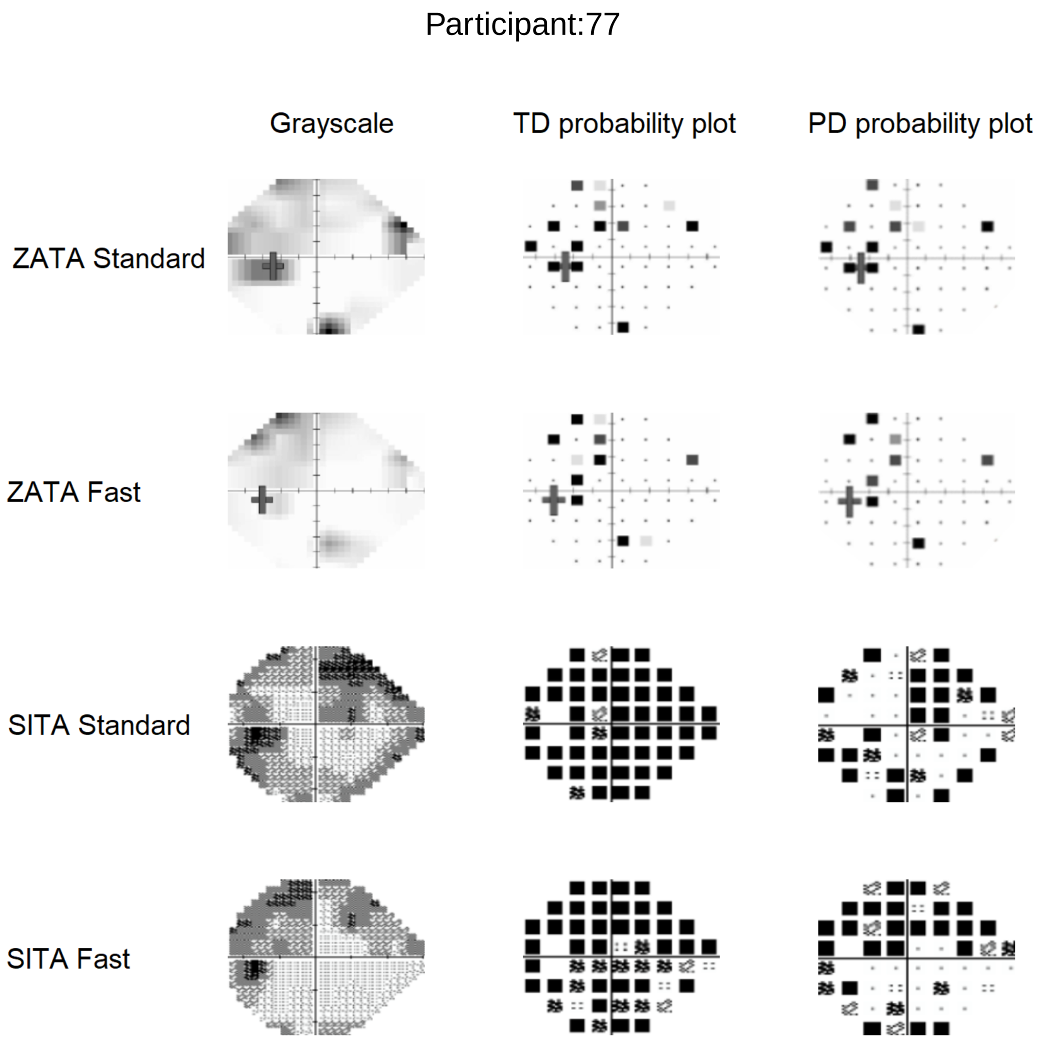
Participant:77
Grayscale
TD probability plot
PD probability plot
ZATA Standard
ZATA Fast
SITA Standard
SITA Fast
